# Methylome-Wide Association Study of Obsessive-Compulsive Disorder

**DOI:** 10.1101/2025.07.18.25331794

**Authors:** Kira D. Höffler, Anne-Kristin Stavrum, Matthew W. Halvorsen, Thorstein Olsen Eide, Kristen Hagen, André Høberg, Nordic OCD and Related Disorders Consortium (NORDiC), Gerd Kvale, James J. Crowley, Jan Haavik, Kerry J. Ressler, Bjarne Hansen, Torsten Klengel, Stephanie Le Hellard

## Abstract

**Background:** Obsessive-compulsive disorder (OCD) is a debilitating psychiatric condition influenced by both genetic and environmental risk factors. Epigenetic modifications, such as DNA methylation, may offer insights into biologically meaningful differences associated with the disorder.

**Methods:** We conducted the largest methylome-wide association study (MWAS) of OCD to date, analyzing saliva DNA samples from 414 individuals with OCD and 384 controls using the Illumina EPICv2 array. Differentially methylated positions (DMPs) and regions (DMRs) were identified, with additional analyses including sex-stratified comparisons, methylation quantitative trait loci (mQTL) mapping, and assessments of cell-type composition differences.

**Results:** We identified 35 DMPs and 17 DMRs associated with OCD, mapping to genes involved in neurotransmission, neurodevelopment, synaptic function, and gene regulation. Sex-stratified analyses revealed additional sex-specific methylation signals, highlighting biological differences between males and females. Most associated loci were influenced by genetic variation (mQTLs). Differences in estimated cell composition and the identification of immune-related genes suggest a potential role for immune system processes. Correlation analyses between brain and saliva methylation indicated that several findings may reflect brain-relevant biology.

**Conclusions:** Our findings emphasize the importance of integrating epigenetic, genetic, and sex-specific data to advance our understanding of OCD. DNA methylation may ultimately contribute to progress toward clinically relevant precision medicine approaches.

## Introduction

Obsessive-compulsive disorder (OCD) is a debilitating mental disorder that affects about 1 in 40 adults over their lifetime^1^. It is characterized by intrusive, distressing obsessions and repetitive compulsive behavior that can significantly impair daily functioning. OCD is a complex, multifactorial disorder influenced by both genetic and environmental factors^2^. Heritability is estimated at around 50%^3^ and a recent large-scale genome-wide association study (GWAS) identified 30 independent genetic loci associated with OCD^4^. Environmental factors, such as perinatal complications and stressful life events, have also been associated with OCD risk^5^.

Despite growing knowledge of OCD risk factors, the underlying biological mechanisms-particularly those involving gene-environment interactions - remain poorly understood. Epigenetic processes, such as DNA methylation, regulate gene expression and are influenced by both genetic and environmental factors. DNA methylation patterns may therefore help identify biological differences between individuals with OCD and healthy controls, potentially pointing to risk-associated genes and pathways.

To date, most epigenetic studies in OCD have focused on candidate genes, identifying potential involvement of *OXTR*^6–9^, *FKBP5*^10^, and *BDNF*^11,12^. Genome-wide DNA methylation studies have produced inconsistent findings, partly due to small sample sizes and methodological issues, which may compromise the interpretability of results^13–20^.

OCD exhibits sex differences in age of onset, symptomatology, and comorbidities. Males are more likely to experience an earlier onset, typically in childhood, and more often present with sexual, religious, or tic-related symptoms^21,22^. In contrast, females tend to develop OCD later, commonly during adolescence or postpartum periods, and more frequently exhibit contamination obsessions and cleaning compulsions. These sex-related patterns contribute to OCD heterogeneity and may also be reflected at the molecular level - a possibility inadequately addressed by traditional sex-adjusted approaches.

In this study, we present the largest epigenome-wide DNA methylation analysis of OCD to date, using the Illumina EPICv2 array on saliva-derived DNA from approximately 800 individuals, including those with OCD and healthy controls. We aimed to identify both sex-specific and cross-sex DNA methylation signals and conducted a methylation quantitative trait loci (mQTL) analysis to identify genetic variants associated with differential methylation. Additionally, we assessed differences in estimated cell-type proportions between cases and controls. By leveraging a larger sample size and integrating genetic and epigenetic data, our goal was to provide novel insights into the molecular mechanisms associated with OCD and support potential biomarker discovery.

## Methods

### Ethics

The study was approved by the Regional Committee of Medical Research Ethics (*REK Vest;* #2019/1097, #2018/52, #2013/543). All participants provided written informed consent.

### Participants

Individuals with OCD were recruited from eight specialized clinics across Norway (Bergen, Førde, Kristiansand, Molde, Oslo, Stord, Trondheim, and Tønsberg) from 2018 to 2023. Eligible individuals were 18 or older with a confirmed OCD diagnosis according to DSM-5^23^ criteria and fluency in Norwegian. Exclusion criteria included current psychosis, bipolar disorder, substance abuse or dependence, intellectual disability, severe eating disorders, and ongoing suicidal ideation. Psychoactive medication use was self-reported. Comorbidities were assessed using the Mini-International Neuropsychiatric Interview^24^ and baseline OCD severity using the Yale– Brown Obsessive–Compulsive Scale (YBOCS)^25^.

Additionally, age- (+/- 3 years) and sex-matched healthy individuals from a previous study^26^ were included. These participants were recruited from the general adult Norwegian population. The majority were randomly selected individuals aged 18-40 years from the Medical Birth Registry of Norway^27^, while the remainder were university students recruited at the University of Bergen between 2005 and 2007. The only inclusion criterion was age over 18 years, and no exclusion criteria were applied. OCD was not specifically screened for in this group, which may have reduced statistical power. However, the prevalence of OCD in this population is assumed to be no higher and possibly lower than in the general adult population.

Differences in age and sex between cases and controls were tested using the Wilcoxon rank-sum test and the chi-square test, respectively.

### Sample Collection

Saliva samples from individuals with OCD were collected under supervision using Oragene-DNA (OG500) kits (*DNA Genotek*, Ottawa, Canada) before psychotherapy. Saliva samples from controls were collected using the same Oragene kits.

### DNA Methylation

DNA was extracted from saliva samples at *HUNT* (Norwegian University of Science and Technology, Trondheim, Norway; all controls, 86 cases) and at *Life & Brain GmbH* (Bonn, Germany; 328 cases). DNA methylation of bisulfite-converted DNA was measured using the Infinium MethylationEPIC v2.0 array (*Illumina*, San Diego, USA) at *Life & Brain GmbH*. Cases and their matched controls were processed on the same slide. Sample slides and positions were randomized and balanced for OCD case-control status, sex, age, and collection site.

### Data Processing

We used R version 4.4.0 for preprocessing, quality control, and data analysis. Plots were created using ggplot2^28^. All scripts are available on GitHub (https://github.com/KiraHoeffler/OCD_MWAS). Due to ethical restrictions and data privacy regulations, individual-level data cannot be shared.

### Quality Control

We used an adapted version of the *CPACOR*^29^ pipeline to quality-control the DNA methylation data. Quality control procedures are detailed in the Supplementary Materials.

### Methylome-Wide Association Analyses (MWAS)

To identify differentially methylated positions (DMPs) associated with the OCD diagnosis, MWAS were conducted separately in males and females using the limma package^30^, applying the following linear model:

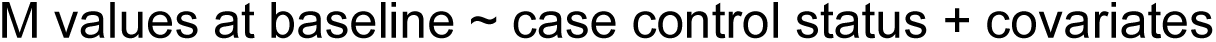

M values were used to represent DNA methylation levels due to their better statistical properties than beta values^31^. Covariates included age at sample collection, the first two principal components (PCs) of estimated cell type proportions (epithelial cells, combined immune cells, and fibroblasts), and the M value of cg05575921 in the *AHRR* gene to account for smoking^32,33^.

To adjust for technical variation, we tested different combinations of control probe PCs (2, 5, 10, and 15). Similarly, to account for population structure, we evaluated models including 2, 5, or 10 ancestry PCs, calculated as described elsewhere^34^. Covariate combinations were assessed based on quantile-quantile (QQ) plots^35^ and genomic inflation factors (λ). For females, a model including 15 control probe PCs and two ancestry PCs was selected; for males, five control probe PCs and two ancestry PCs were optimal. We used the vif() function from the car package^36^ to confirm that there were no multicollinearity issues among the covariates.

Bacon correction was applied to the p values^37^. The QQ plots for the selected models are in **Figure S1A-B**, with genomic inflation factors (λ) of 1.08 in females, 1.02 in males, indicating minimal inflation. P values were false discovery rate (FDR)-adjusted.

### Cross-Sex Meta-Analysis

To identify DNA methylation signals associated with OCD diagnosis consistent across sex, we conducted a meta-analysis of female- and male-specific MWAS results using CpGs common to both datasets (excluding probes on the Y chromosome)^38^. The analysis was performed using a random effects model and implemented via the metagen() function from the meta package^39^, because different effect sizes were expected for males and females. For each CpG, we extracted heterogeneity statistics including τ^2^ (tau-squared), I^2^, Cochran’s Q, and its associated p-value. A QQ plot of the resulting p values is shown in **Figure S1C**, with λ of 0.92, indicating no inflation. P values were adjusted for multiple testing using FDR correction.

### Sex Interaction

To assess whether the results were sex-specific, we fitted a linear model including a sex-by-diagnosis interaction term across both male and female samples, using either the average M values across DMRs or the M-values of individual DMPs as the outcome. The model was adjusted for age, smoking, two cell type PCs, 15 control probe PCs (excluding PC4 due to multicollinearity with sex), and two ancestry PCs. P values were FDR-adjusted.

### Differentially Methylated Regions (DMRs)

To identify DMRs, we used ENmix-combp^40,41^ with a maximum probe distance of 750 bp as previously recommended^42^ and a strict seed p value threshold of 0.001. We required each DMR to contain at least three nominally significant DMPs, with more than half of the DMPs within the region meeting nominal significance. DMRs with a Sidak-corrected p value < 0.05 were considered statistically significant.

### Methylation Quantitative Trait Loci (mQTL) Analysis

Genotyping data were processed following the quality control procedures described in the latest PGC OCD GWAS study^4^. mQTL analysis details are provided in the Supplementary Materials.

### Gene Ontology (GO) Analysis

We used the goMeth() function from the missMethyl package (v1.41.0)^43^ to perform GO enrichment analysis, focusing on biological processes, separately for the female-specific, male-specific, and cross-sex meta-analysis summary statistics. The analysis included the top 0.5% of CpGs, using all CpGs tested in the respective MWAS as the background. P values were adjusted using FDR correction.

### Cell Type Proportion Analyses

We compared the estimated cell type proportions for B cells, CD4T+ cells, epithelial cells, fibroblasts, monocytes, natural killer cells, and neutrophils between individuals with OCD and healthy controls. Eosinophils and CD8+ T cells were excluded due to their very low proportions, which were often undetectable in samples. These analyses were performed using the following linear model:

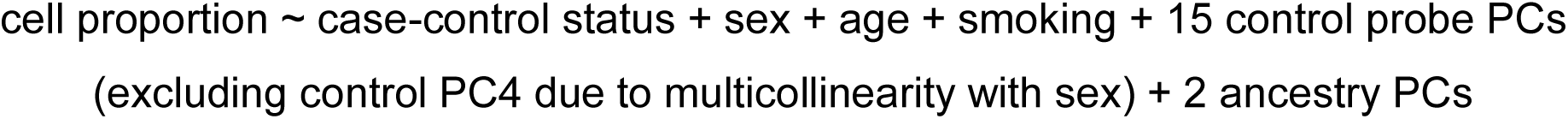

To adjust for multiple tests while accounting for cell type correlation, we applied the method proposed by Li and Ji (2005)^44^, which estimated 1.82269 independent tests. We repeated the analyses in a subset of individuals with OCD who were not taking any psychoactive medication.

To test for sex-specific effects, we fit linear models including a diagnosis-by-sex interaction term for each cell type, adjusting for the same covariates as in the main model.

## Results

### DNA methylation associated with OCD

We analyzed DNA methylation in 414 individuals with OCD and 384 healthy controls after quality control (**Table 1**). We conducted sex-stratified analyses, followed by a meta-analysis to identify effects consistent across sexes. This approach was chosen over a traditional sex-adjusted model to (1) detect both shared and sex-specific methylation signals, (2) include sex chromosomes, and (3) improve discovery power based on previous evidence^38^. Cases and controls were age- and sex-matched, with no significant group differences (**Table 1**); sensitivity analyses excluding medicated individuals and those with comorbidities are described later.

**Table 1:**
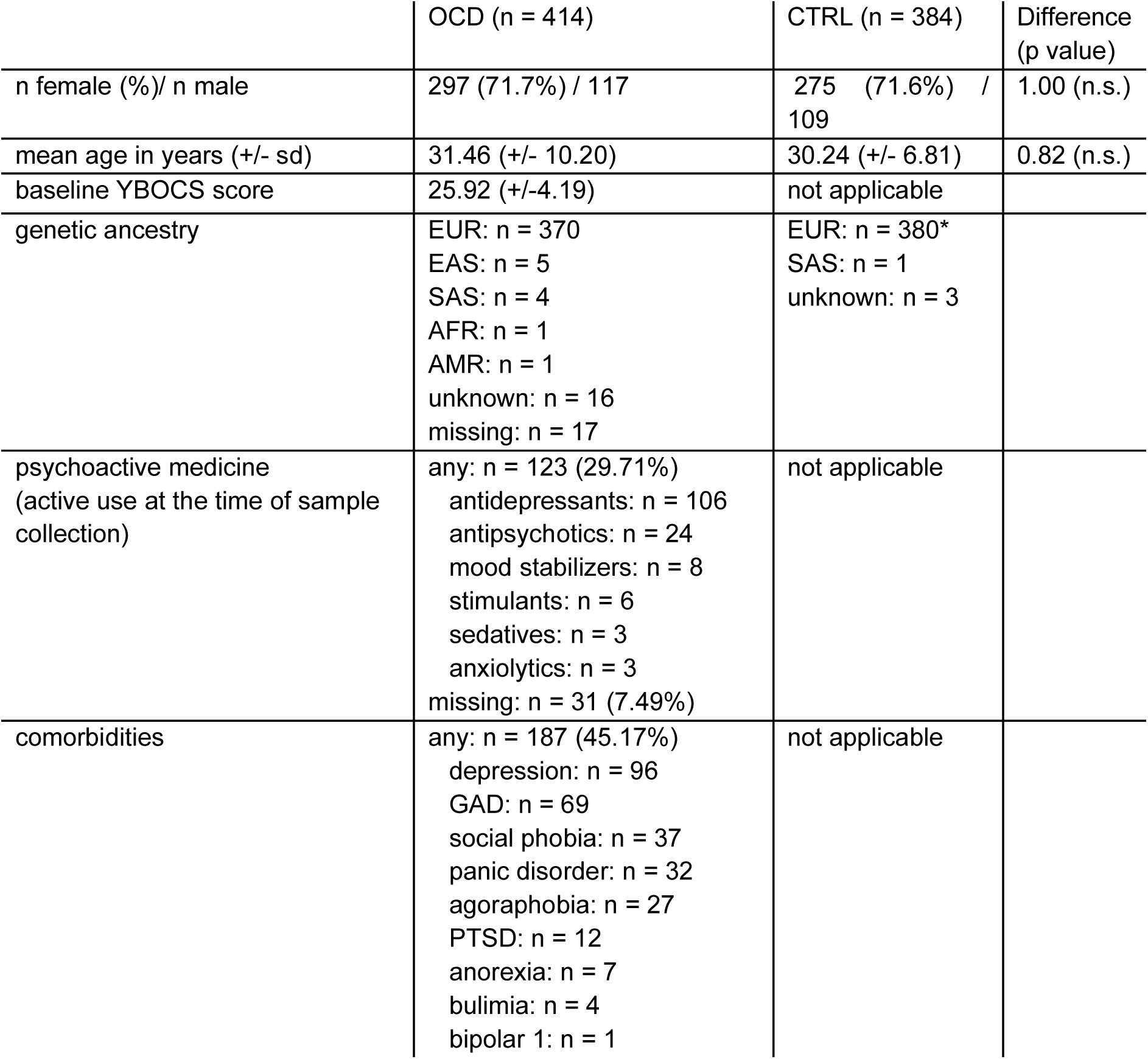
Demographic Characteristics. . Genetic ancestry was inferred from genotype data using the 1000 Genomes as reference^45^ as previously described^46^. “Unknown genetic ancestry” indicates that no specific ancestry group matched perfectly, likely due to mixed genetic ancestry. * Genetically predicted ancestry was not available for 248 control individuals; however, their self-reported ancestry was European, and all clustered within the European group in the ancestry PCA (see Methods). AFR: African, AMR: Native American, CTRL: Healthy Control, EAS: East Asian, EUR: European, GAD: Generalized Anxiety Disorder, n.s.: not significant, OCD: Obsessive-Compulsive Disorder, PTSD: Post-Traumatic Stress Disorder, SAS: South Asian, YBOCS: Yale–Brown Obsessive–Compulsive Scale

|  | OCD (n = 414) | CTRL (n = 384) | Difference (p value) |
| --- | --- | --- | --- |
| n female (%) / n male | 297 (71.7%) / 117 | 275 (71.6%) / 109 | 1.00 (n.s.) |
| mean age in years (+/- sd) | 31.46 (+/- 10.20) | 30.24 (+/- 6.81) | 0.82 (n.s.) |
| baseline YBOCS score | 25.92 (+/- 4.19) | not applicable |  |
| genetic ancestry | EUR: n = 370<br>EAS: n = 5<br>SAS: n = 4<br>AFR: n = 1<br>AMR: n = 1<br>unknown: n = 16<br>missing: n = 17 | EUR: n = 380*<br>SAS: n = 1<br>unknown: n = 3 |  |
| psychoactive medicine (active use at the time of sample collection) | any: n = 123 (29.71%)<br>antidepressants: n = 106<br>antipsychotics: n = 24<br>mood stabilizers: n = 8<br>stimulants: n = 6<br>sedatives: n = 3<br>anxiolytics: n = 3<br>missing: n = 31 (7.49%) | not applicable |  |
| comorbidities | any: n = 187 (45.17%)<br>depression: n = 96<br>GAD: n = 69<br>social phobia: n = 37<br>panic disorder: n = 32<br>agoraphobia: n = 27<br>PTSD: n = 12<br>anorexia: n = 7<br>bulimia: n = 4<br>bipolar 1: n = 1 | not applicable |  |

The cross-sex meta-analysis identified 35 differentially methylated positions (DMPs; single CpG sites, **Table 2**, **Table S1A**) and 17 differentially methylated regions (DMRs; clusters of CpG sites; **Table 3**; **Table S2A**) significantly associated with OCD (highlighted in the Manhattan plot in **Figure 1**). Examples are shown in **Figure 2**, full results are in **Supplementary Files 2 and 3**.

**Figure 1:**
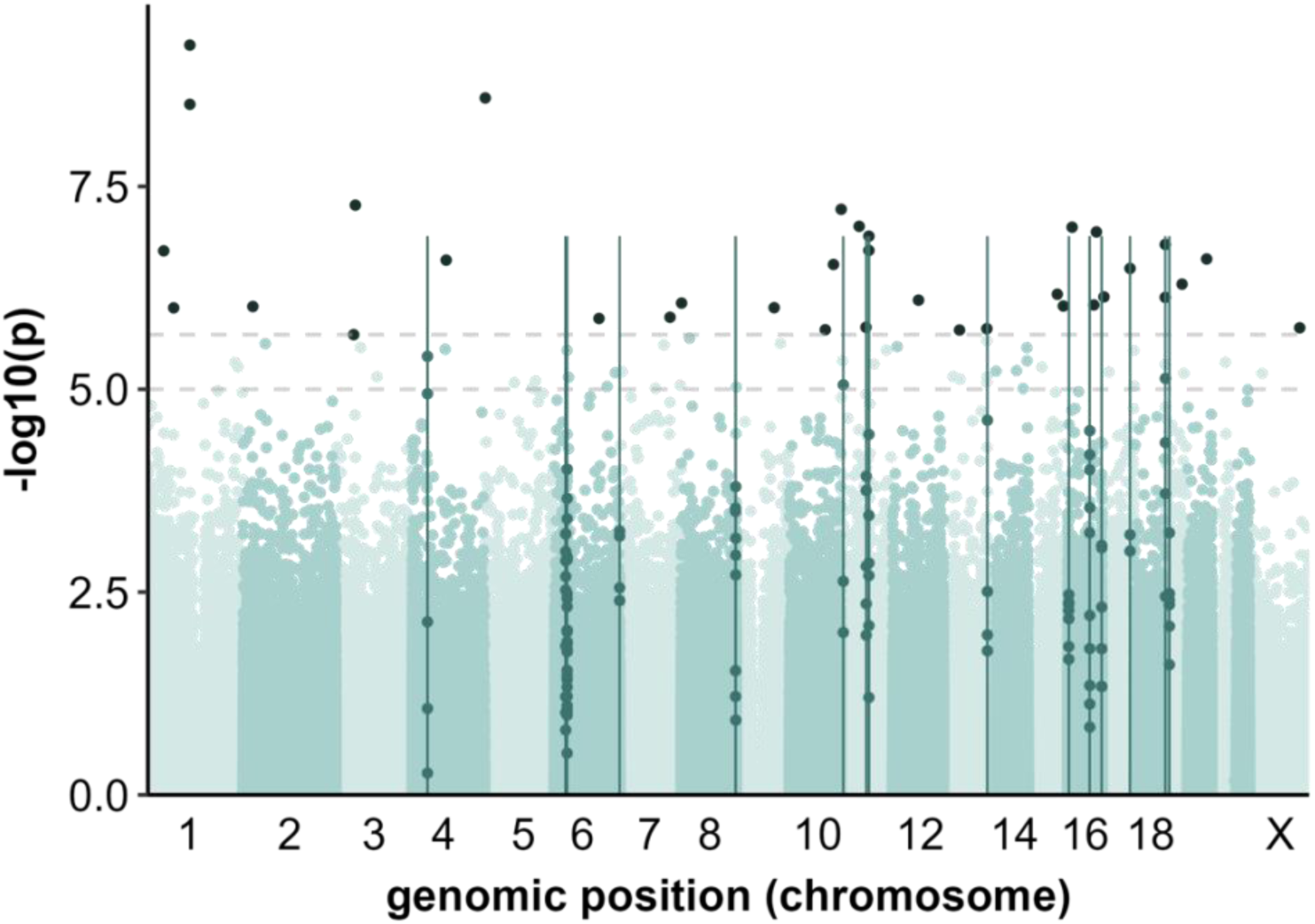
Manhattan plot of the cross-sex meta-analysis. Differentially methylated positions (dots above the upper dashed line; FDR threshold) and regions (vertical lines) are highlighted in dark green. FDR: false discovery rate.

**Figure 2:**
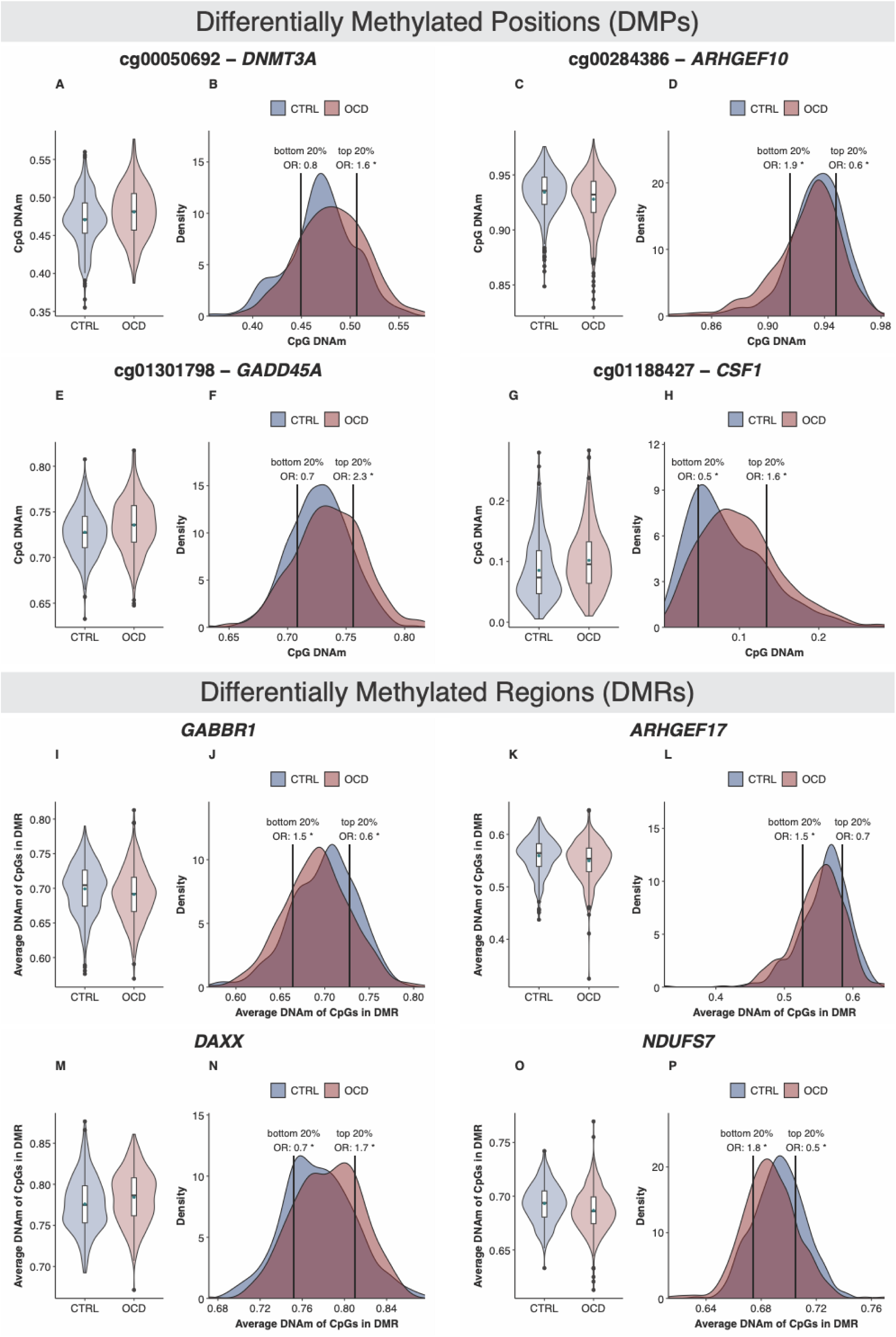
Examples of DNA methylation differences between individuals with OCD and healthy controls in DMPs (A-H) and DMRs (I-P). Respective left plots are box violin plots showing DNA methylation differences between cases and controls. Respective left plots are density plots of DNA methylation levels, with OCD odds ratios (ORs) calculated for the highest and lowest 20% of methylation values (indicated by vertical lines). Significant ORs are marked with an asterisk (*). CTRL: healthy control, DMP: differentially methylated position, DMR: differentially methylated region, DNAm: DNA methylation, OCD: obsessive-compulsive disorder

**Table 2:** Differentially methylated positions (DMPs) associated with OCD identified in the cross-sex meta-analysis and female-specific analysis. No significant DMPs were observed in the male-specific analysis. The DMPs were annotated using GRCh38/hg38. adjusted: adjusted, chr: chromosome, CTRL: healthy control, diff: difference, OCD: obsessive-compulsive disorder, prox: proximal, regul: regulatory, SE: standard error.

| cglD | location | gene | effect | SE | p value | adj p value | regul. Region | CpG island | median diff (OCD-CTRL) |
| --- | --- | --- | --- | --- | --- | --- | --- | --- | --- |
| cross-sex meta-analysis |  |  |  |  |  |  |  |  |  |
| cg01188427 | chr1:109934023 | *CSF1* | 0.510 | 0.082 | 5.72E-10 | 4.60E-04 | distal_enhancer |  | 0.0218 |
| cg03863908 | chr5:1123623 | *SLC12A7* | -0.186 | 0.031 | 2.57E-09 | 8.25E-04 | distal_enhancer |  | -0.0091 |
| cg19627034 | chr1:109934046 | *CSF1* | 0.355 | 0.060 | 3.07E-09 | 8.25E-04 | distal_enhancer |  | 0.0197 |
| cg03565274 | chr3:50592277 | *HEMK1* | 0.157 | 0.029 | 5.38E-08 | 9.74E-03 | GeneBody |  | 0.0186 |
| cg11415512 | chr11:1108571 | *MUC2* | -0.142 | 0.026 | 6.05E-08 | 9.74E-03 | promoter |  | -0.0084 |
| cg02456934 | chr16:75074611 | *ZNRF1* | 0.125 | 0.024 | 1.15E-07 | 1.16E-02 | GeneBody |  | 0.0127 |
| cg09816471 | chr16:11668004 | *SNN* | 0.156 | 0.029 | 1.01E-07 | 1.16E-02 | prox_enhancer | Island | 0.014 |
| cg17428739 | chr11:47986426 | *PTPRJ* | 0.121 | 0.023 | 9.81E-08 | 1.16E-02 | distal_enhancer |  | 0.0163 |
| cg02776498 | chr11:73343140 | *ARHGEF17* | -0.076 | 0.014 | 1.30E-07 | 1.16E-02 | GeneBody | Island | -0.0063 |
| cg04497870 | chr1:41811613 | *HIVEP3* | 0.139 | 0.027 | 1.97E-07 | 1.32E-02 | distal_enhancer |  | 0.0173 |
| cg18693051 | chr19:1161801 | *SBNO2* | 0.169 | 0.032 | 1.64E-07 | 1.32E-02 | distal_enhancer | S_Shelf;N_Shore | 0.0188 |
| cg20864214 | chr11:73343076 | *ARHGEF17* | -0.073 | 0.014 | 1.94E-07 | 1.32E-02 | promoter | Island | -0.0069 |
| cg02462015 | chr4:89283414 | *GPRIN3* | 0.154 | 0.030 | 2.56E-07 | 1.47E-02 | distal_enhancer |  | 0.0145 |
| cg20519581 | chr20:50343307 | *LINC01271* | 0.110 | 0.021 | 2.48E-07 | 1.47E-02 | distal_enhancer | N_Shore | 0.0119 |
| cg15607402 | chr10:114636179 | *ABLIM1* | 0.072 | 0.014 | 2.89E-07 | 1.55E-02 | distal_enhancer | S_Shelf | 0.0063 |
| cg27492584 | chr17:72603168 | *LINC00511* | 0.233 | 0.046 | 3.24E-07 | 1.63E-02 | promoter |  | 0.0146 |
| cg17725001 | chr19:44804013 |  | 0.109 | 0.022 | 5.06E-07 | 2.40E-02 |  |  | 0.0121 |
| cg00050692 | chr2:25302008 | *DNMT3A* | 0.085 | 0.017 | 9.55E-07 | 2.96E-02 | GeneBody |  | 0.0104 |
| cg00284386 | chr8:1838041 | *ARHGEF10* | -0.207 | 0.042 | 8.66E-07 | 2.96E-02 | GeneBody | N_Shelf | -0.0035 |
| cg01301798 | chr1:67723714 | *GADD45A* | 0.087 | 0.018 | 9.92E-07 | 2.96E-02 | distal_enhancer |  | 0.0087 |
| cg01322839 | chr9:98136334 | *TRIM14* | 0.114 | 0.023 | 9.87E-07 | 2.96E-02 | distal_enhancer |  | 0.0075 |
| cg01593826 | chr15:75112684 |  | -0.172 | 0.035 | 6.74E-07 | 2.96E-02 |  |  | -0.0125 |
| cg17325206 | chr15:90852879 | *RN7SL363P* | 0.092 | 0.019 | 9.42E-07 | 2.96E-02 | distal_enhancer |  | 0.0122 |
| cg17760714 | chr12:67647958 | *DYRK2* | 0.130 | 0.026 | 7.97E-07 | 2.96E-02 | prox_enhancer | N_Shore | 0.0017 |
| cg21655515 | chr16:68258201 | *PLA2G15* | 0.162 | 0.033 | 9.13E-07 | 2.96E-02 | GeneBody |  | 0.008 |
| cg22082184 | chr17:4606021 | *LINC01996* | 0.131 | 0.026 | 7.26E-07 | 2.96E-02 | distal_enhancer | N_Shore | 0.0127 |
| cg22325408 | chr19:1042148 | *ABCA7* | -0.100 | 0.020 | 7.37E-07 | 2.96E-02 | exon1 | S_Shore | -0.0108 |
| cg10000558 | chr6:116392362 | *DSE* | 0.105 | 0.022 | 1.34E-06 | 3.72E-02 | GeneBody |  | 0.0129 |
| cg11709399 | chr7:130903767 | *MIR29A* | 0.090 | 0.018 | 1.30E-06 | 3.72E-02 | distal_enhancer |  | 0.009 |
| cg02750385 | chr13:41311418 | *NAA16* | 0.107 | 0.022 | 1.86E-06 | 4.41E-02 | exon1 | Island | 0.0022 |
| cg06464468 | chr11:65646942 | *MIR4489* | -0.076 | 0.016 | 1.72E-06 | 4.41E-02 | distal_enhancer | Island | -0.0069 |
| cg13314489 | chrX:131226838 |  | -0.141 | 0.030 | 1.75E-06 | 4.41E-02 |  |  | -0.0049 |
| cg15294391 | chr10:93309333 | *RPL17P34* | 0.104 | 0.022 | 1.85E-06 | 4.41E-02 | distal_enhancer |  | 0.0123 |
| cg19193420 | chr13:112592994 | *TUBGCP3;LOC124903251* | 0.134 | 0.028 | 1.79E-06 | 4.41E-02 | GeneBody;GeneBody |  | 0.0129 |
| cg10335493 | chr3:46204869 | *CCR1* | -0.165 | 0.035 | 2.12E-06 | 4.88E-02 | GeneBody |  | -0.0027 |
| female-specific analysis |  |  |  |  |  |  |  |  |  |
| cg01188427 | chr1:109934023 | *CSF1* | 0.554 | 0.1 | 3E-08 | 2.44E-02 | distal_enhancer |  | 0.0272 |

**Table 3:** Differentially methylated regions (DMRs) associated with OCD from sex-specific and cross-sex meta-analyses. . The DMRs were identified using ENmix-combp^40,41^ and annotated using GRCh38/hg38. adj: adjusted, chr: chromosome, CTRL: healthy control, diff: difference, n: number of CpGs in DMR, OCD: obsessive-compulsive disorder, regul.: regulatory, TSS: transcription start site.

| location | p | sidak-adj p | n | gene | regul. region | CpG island | median diff<br>(OCD-CTRL) |
| --- | --- | --- | --- | --- | --- | --- | --- |
| cross-sex meta-analysis |  |  |  |  |  |  |  |
| chr11:73342604-73343357 | 2.93E-18 | <2.2e-16 | 8 | *ARHGEF17* | promoter;prox_enhancer;GeneBody | Island;Shore | -0.0108 |
| chr16:57797833-57798398 | 1.13E-13 | 1.61E-10 | 10 | *KIFC3* | TSS200;TSS1500;promoter | Shelf | -0.0054 |
| chr6:33320403-33320823 | 3.72E-13 | 7.13E-10 | 9 | *DAXX* | distal_enhancer | Shelf;Shore | 0.0114 |
| chr19:1395366-1395648 | 5.63E-12 | 1.61E-08 | 4 | *NDUFS7* | GeneBody | Shelf;Shore | -0.0076 |
| chr6:29631235-29631614 | 5.19E-11 | 1.10E-07 | 10 | *GABBR1* | prox_enhancer;TSS200;TSS1500;exon1 | Shelf;Shore | -0.0125 |
| chr11:66334584-66334882 | 2.49E-10 | 6.74E-07 | 5 | *RIN1* | GeneBody | Island;Shore | -0.0065 |
| chr17:72603168-72603399 | 2.15E-10 | 7.49E-07 | 3 | *LINC00511* | promoter;GeneBody |  | 0.0112 |
| chr19:11673832-11674248 | 3.88E-09 | 7.50E-06 | 6 | *ZNF833P;HNRNPA1P10* | TSS200;TSS1500;distal_enhancer;GeneBody;exon1 | Shore;Island | -0.0058 |
| chr6:33277861-33278232 | 3.99E-09 | 8.65E-06 | 15 | *B3GALT4* | exon1;prox_enhancer | Shore | 0.0028 |
| chr16:3012295-3012653 | 4.11E-09 | 9.25E-06 | 7 | *CLDN9* | prox_enhancer;TSS1500 | Shore | -0.0075 |
| chr8:143024153-143024307 | 5.24E-09 | 2.74E-05 | 5 | *LY6E* | distal_enhancer | Shore;Island | 0.0093 |
| chr6:170288238-170288501 | 1.23E-08 | 3.76E-05 | 4 | *FAM120B;DLL1* | distal_enhancer;GeneBody | Island;Shore | -0.0082 |
| chr13:114015296-114015519 | 7.09E-08 | 2.56E-04 | 4 | *C13orf46;RASA3* | distal_enhancer;GeneBody | Island;Shelf;Shore | -0.0052 |
| chr11:6630483-6630758 | 8.85E-08 | 2.59E-04 | 3 | *DCHS1* | GeneBody | Island | -0.0048 |
| chr16:88881388-88881536 | 6.15E-08 | 3.35E-04 | 6 | *CBFA2T3* | GeneBody | Shore;Island;Shelf | -0.0053 |
| chr4:40516007-40516127 | 1.68E-07 | 1.13E-03 | 5 | *RBM47* | promoter |  | 0.0058 |
| chr8:142454076-142454220 | 7.14E-07 | 3.99E-03 | 4 | *BAI1;ADGRB1* | distal_enhancer;GeneBody | Shore;Island | -0.0034 |
| females |  |  |  |  |  |  |  |
| chr11:73342604-73343141 | 1.4E-15 | 2.2E-12 | 7 | *ARHGEF17* | promoter;GeneBody;prox_enhancer | Island | -0.0098 |
| chr1:44807307-44807606 | 1.2E-12 | 3.1E-09 | 5 | *DYNLT4;BTBD19* | promoter;TSS1500 | Shore | 0.0066 |
| chr17:59838304-59838413 | 3.4E-10 | 2.5E-06 | 4 | *MIR21;VMP1* | distal_enhancer;GeneBody |  | 0.0311 |
| chr17:81821611-81822172 | 4.5E-09 | 6.5E-06 | 14 | *MCRIP1* | distal_enhancer | Shelf | 0.0066 |
| chr6:33320403-33320596 | 3E-09 | 1.2E-05 | 7 | *DAXX* | distal_enhancer | Shelf;Shore | 0.0125 |
| chr19:1395366-1395648 | 4.6E-09 | 1.3E-05 | 4 | *NDUFS7* | GeneBody | Shelf;Shore | -0.0067 |
| chr11:66334584-66334882 | 2.8E-08 | 7.6E-05 | 5 | *RIN1* | GeneBody | Island;Shore | -0.0061 |
| chr11:92883153-92883450 | 2.8E-08 | 7.7E-05 | 5 | *EEF1A1P49* | distal_enhancer | Island | -0.0054 |
| chr17:72603168-72603399 | 2.2E-08 | 7.8E-05 | 3 | *LINC00511* | promoter;GeneBody |  | 0.012 |
| chr6:170288238-170288501 | 3.7E-08 | 0.00011 | 4 | *FAM120B;DLL1* | distal_enhancer;GeneBody | Island;Shore | -0.0091 |
| chr6:33277924-33278214 | 5.2E-08 | 0.00014 | 12 | *B3GALT4* | exon1;prox_enhancer | Shore | 0.0046 |
| chr5:179138720-179138937 | 4.4E-08 | 0.00016 | 3 | *ADAMTS2* | GeneBody | Shore;Island | -0.0048 |
| chr1:230332865-230333028 | 1E-07 | 0.0005 | 4 | *PGBD5* | distal_enhancer | Island | -0.0308 |
| chr1:24931014-24931139 | 1.7E-07 | 0.00108 | 9 | *RUNX3* | prox_enhancer | Island | -0.0041 |
| **males** |  |  |  |  |  |  |  |
| chr6:151325405-151325683 | 8.9E-10 | 2.6E-06 | 7 | *AKAP12* | promoter;exon1 | Island;Shore | 0.0028 |
| chr13:30932550-30932827 | 1.3E-09 | 3.8E-06 | 6 | *TEX26;TEX26-AS1* | promoter |  | -0.0208 |
| chr2:21043855-21044341 | 3.1E-09 | 5.1E-06 | 9 | *APOB* | promoter;TSS1500 | Island;Shore | -0.0205 |
| chr16:57798088-57798398 | 3.6E-09 | 9.3E-06 | 7 | *KIFC3* | TSS200;TSS1500;promoter | Shelf | -0.0111 |
| chr15:26629055-26629369 | 1.6E-08 | 4.2E-05 | 6 | *GABRB3* | promoter;exon1 | Island | 0.0288 |
| chr19:2095365-2095645 | 2.6E-08 | 7.4E-05 | 4 | *MOB3A* | prox_enhancer | Island;Shore | 0.0176 |
| chr12:130337711-130337817 | 1.1E-06 | 0.00844 | 4 | *PIWIL1;LOC101927786* | TSS200;TSS1500;promoter | Island | 0.0439 |

In the sex-specific analysis, we identified one DMP and 14 DMRs in females, and seven DMRs in males (**Tables 2 and 3**; Manhattan plots in **Figures S2 and S3**; **Table S1B-C**; **Table S2B**). A significant sex–methylation interaction was observed in six of seven DMRs identified in males (all except the DMR annotated to *KIFC3*; **Table S3**, **Figure S4**). In contrast, no sex interaction remained significant for the loci identified in females after correction for multiple testing (**Table S3**). All DMPs and DMRs identified in the sex-specific analyses are visualized in **Supplementary Files 2 and 3**.

### Sensitivity analyses

We conducted sensitivity analyses using OCD subgroups consisting of non-medicated individuals (females: n = 191; males: n = 69) and individuals without psychiatric comorbidities (females: n = 160; males: n = 67), compared to the full sample of healthy controls. DNA methylation at all DMPs and DMRs remained significantly associated with OCD case-control status across analyses. One male-specific DMR (GABRB3) lost significance in the medication-free subgroup, likely due to reduced sample size rather than medication effects. (**Table S4**).

### Methylation Quantitative Trait Locus (mQTL) Analysis

We performed mQTL analysis to assess whether methylation was influenced by genetic variation. Genotyping data was available for 371 individuals with OCD and 233 healthy controls, all of European ancestry, in the cross-sex meta-analysis. A total of 670 significant mQTLs were identified across 17 DMPs (48.6% of all DMPs) and 10 DMRs (58.5% of all DMRs), with the strongest association observed for a DMR annotated to *ARHGEF17* (adjusted p = 4.22E-54; **Table S5A**). After linkage disequilibrium (LD) clumping, 50 independent mQTLs remained for these DMRs (**Table S6A**).

We then tested the association between these mQTL SNP genotypes and OCD case-control status in the combined sample across both sexes. One mQTL, rs78454796, associated with DNA methylation at cg11415512 annotated to *MUC2*, remained significantly associated with OCD diagnosis after multiple testing correction (adjusted p = 6.30E-35). The G allele was linked to both lower methylation at the DMP and OCD diagnosis, consistent with OCD-associated hypomethylation (**Figure 3A-D**).

**Figure 3:**
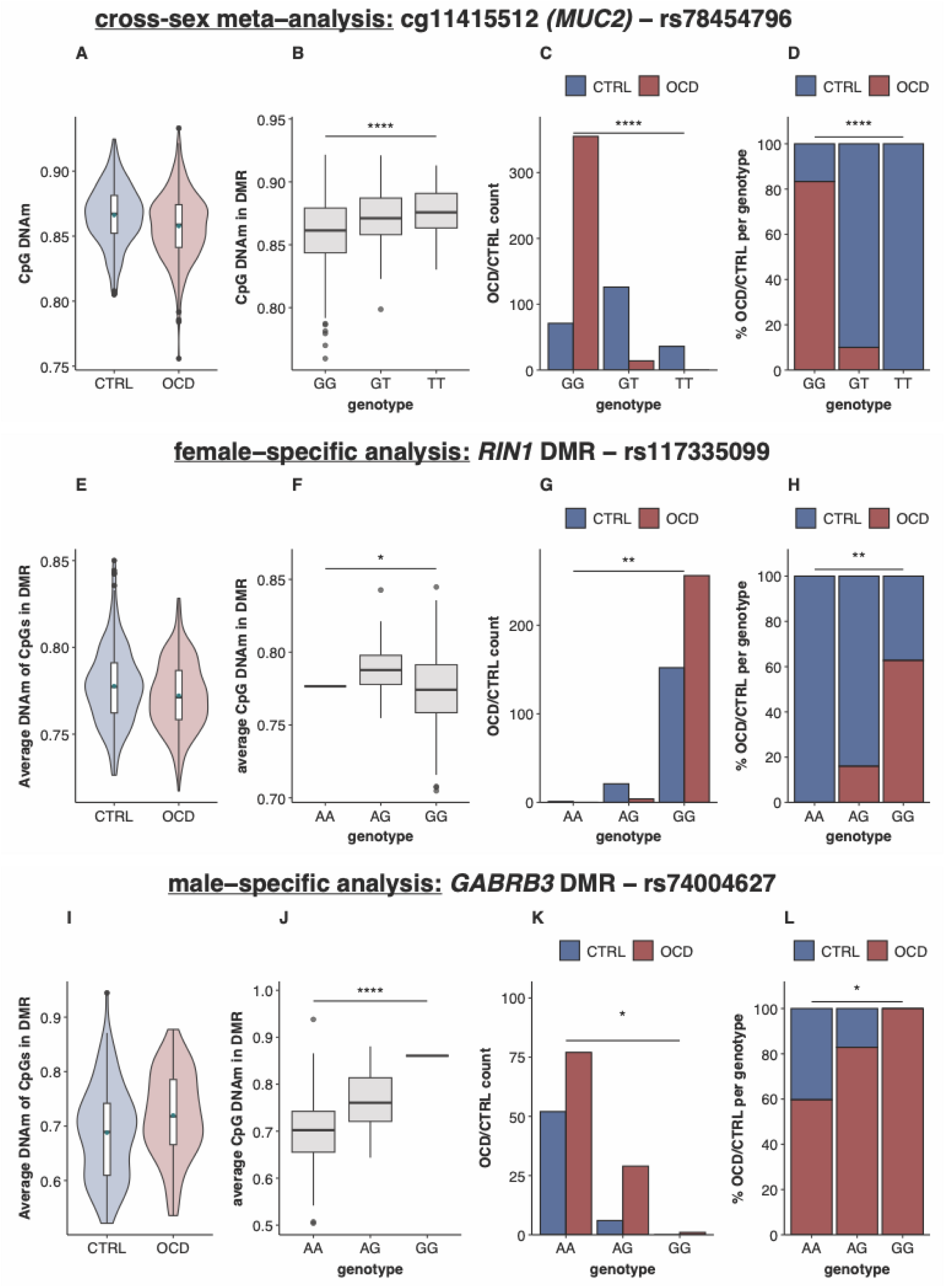
Selected mQTLs associated with both DNA methylation and OCD case–control status. (A, E, **I)** Differences in average DNA methylation between individuals with OCD and healthy controls. **(B, F, J)** Differences in average DNA methylation by genotype of the respective SNP. **(C, G, K)** Counts of OCD cases and healthy controls per genotype of the respective SNP. **(D, H, L)** Percentage of OCD cases and healthy controls per genotype of the respective SNP. CTRL: control, DMP: differentially methylated position, DMR: differentially methylated region, DNAm: DNA methylation, mQTL: methylation quantitative trait locus, ns: not significant, OCD: obsessive-compulsive disorder, SNP: single nucleotide polymorphism. ****: Bonferroni-corrected p ≤ 0.0001, **: Bonferroni-corrected p ≤ 0.01, *: Bonferroni-corrected p ≤ 0.05.

Additionally, seven other mQTLs showed nominal associations with OCD case-control status but did not survive multiple testing correction (**Table S6A**).

In the sex-specific analyses, 316 mQTLs were identified in females (264 individuals with OCD, 175 controls) across 10 DMRs (71.4% of all female-specific DMRs). The strongest association was observed for a DMR annotated to *PGBD5* (adjusted p = 5.42E-44; **Table S5B**). After LD clumping, 22 independent mQTLs remained (**Table S6B**). One mQTL SNP influencing DNA methylation in a DMR annotated to *RIN1* - rs117335099 - was also significantly associated with OCD case-control status after multiple testing correction (adjusted p = 1.93E-03; **Figure 3E-H**). Three additional independent mQTLs showed nominal associations with case-control status, though they did not remain significant after multiple testing correction.

In males (107 individuals with OCD, 58 controls), 128 mQTLs were detected across five DMRs (71.4% of all male-specific DMRs). The strongest mQTL association was found for a DMR annotated to *AKAP12* (adjusted p = 1.27E-15; **Table S5C**). After pruning, seven independent mQTLs remained (**Table S6C**). One mQTL SNP associated with DNA methylation in a DMR annotated to *GABRB3* - rs74004627 - was also significantly associated with OCD diagnosis (adjusted p = 1.87E-02; **Figure 3I-L**).

We identified SNPs in LD (R^2^ ≥ 0.7) with mQTL SNPs that were significantly associated with OCD diagnosis after multiple testing correction. In the male-specific analysis, the lead mQTL SNP rs74004627 had 24 SNPs in LD, all of which showed nominally significant associations with OCD case-control status (p values ranging from 2.67E-03 to 1.08E-02; **Table S7**). For the mQTLs identified in the cross-sex meta-analysis and the female-specific analysis, no LD SNPs were available.

### Gene Ontology (GO)

No GO term remained significant after multiple testing correction (**Table S8**).

### GWAS Atlas

We assessed whether genes annotated to OCD-associated DMRs and DMPs from the cross-sex meta-analysis overlapped with prior GWAS findings (p value ≤ 5E-08) using the PheWAS catalog^47^. For the identified genes of the cross-sex meta-analysis, 487 associations were identified in previous GWAS.

The most frequent categories with significant GWAS hits among the identified genes were immunological and metabolic traits (**Figure 4**). Interestingly, five genes (*CBFA2T3, DAXX, GABBR1, HEMK1, PLA2G15*) were associated with psychiatric traits, two genes (*GABBR1* and *ABCA7*) with neurological traits, and two (*BAI1, PLA2G15*) with cognitive traits. *GABBR1*, for instance, was linked to neuroticism, mental health visits, and fatigue.

**Figure 4:**
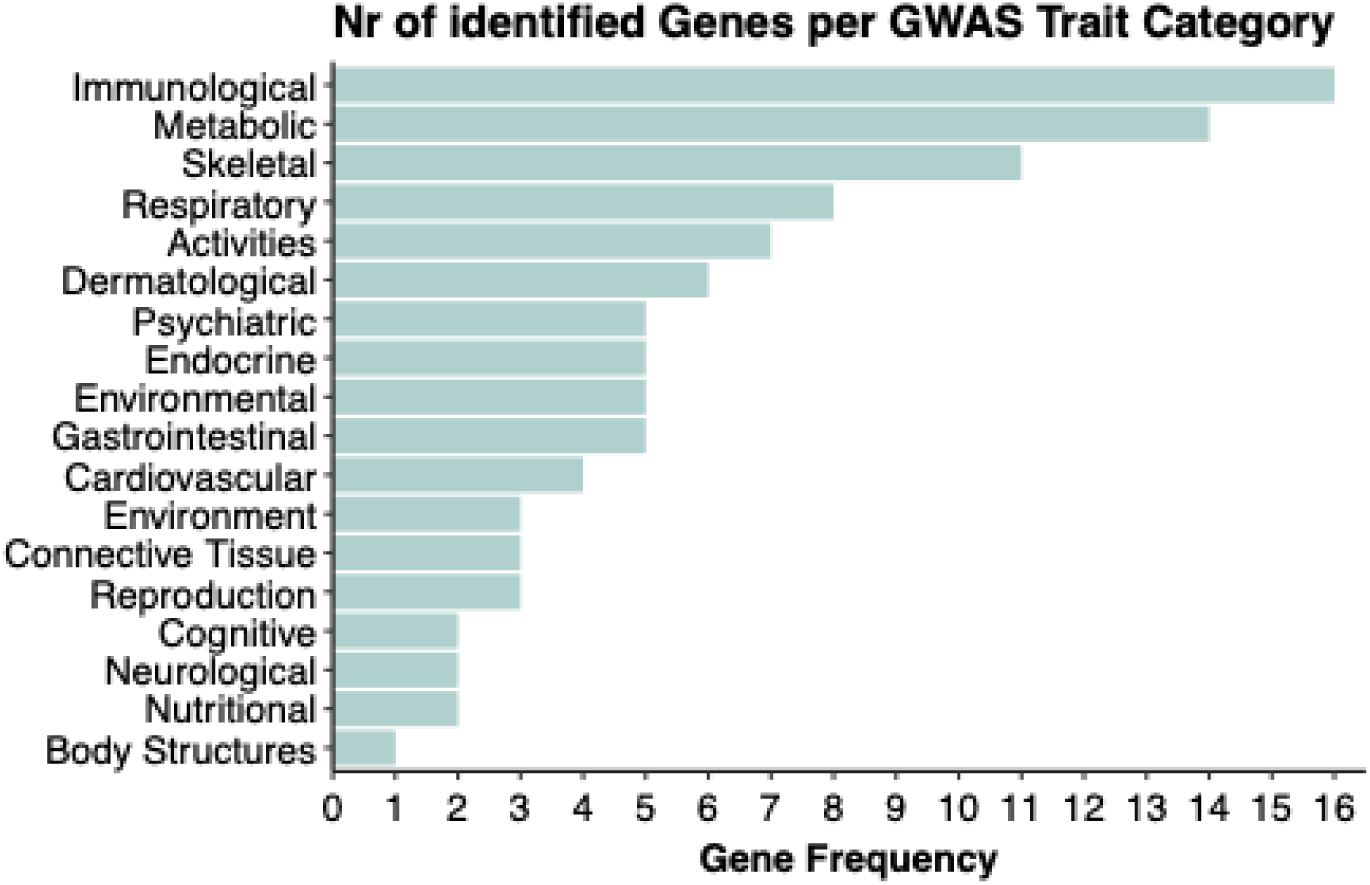
Number of genes annotated to OCD-associated DMRs and DMPs with GWAS hits by category. Genes annotated to DMRs and DMPs from the cross-sex meta-analysis. Extracted from the PheWAS catalog of the GWAS Atlas ^47^. DMP: differentially methylated position, DMR: differentially methylated region, GWAS: genome-wide association study, Nr: number

Details on the GWAS associations, including those from sex-specific analyses, are provided in **Table S9**.

### Saliva-brain correlation

To assess brain relevance of saliva methylation signals, we examined saliva-brain DNA methylation correlations using previously published data^48^.

At the DMP level (**Table S10A**), only one unannotated CpG showed a significant correlation between saliva and brain. Six DMPs exhibited a medium absolute correlation (|ρ| > 0.3), while nine DMPs had no available data due to differences in array coverage.

At the DMR level (**Table S10B**), 17 DMRs contained at least one CpG with a significant saliva-brain correlation (eight from the cross-sex meta-analysis, four from the female-specific analysis, and five from the male-specific analysis). Eleven DMRs included at least one CpG with a high absolute correlation (|ρ| > 0.5), with several showing multiple CpGs with medium to high correlations. Fifteen DMRs had at least one CpG with a medium correlation (|ρ| > 0.3). One DMR lacked data for all its CpGs. Of note, three DMRs showed strong, significant correlations across all CpGs within the region: *PGBD5* (female-specific), as well as *GABRB3* and *PIWIL1;LOC101927786* (both male-specific).

### Cell Type Proportion Analyses

Saliva cell type proportions varied substantially across samples (**Figure S5**). After multiple testing correction, significant differences between individuals with OCD and healthy controls were observed for all analyzed cell types except one (**Figure 5**; **Table S11A**): neutrophils (adjusted p = 8.80E-06), monocytes (adjusted p = 1.19E-05), CD4+ T cells (adjusted p = 1.18E-05), NK cells (adjusted p = 7.07E-05), epithelial cells (adjusted p = 7.54E-05), B cells (adjusted p = 2.89E-04), and fibroblasts (adjusted p = 0.051).

**Figure 5:**
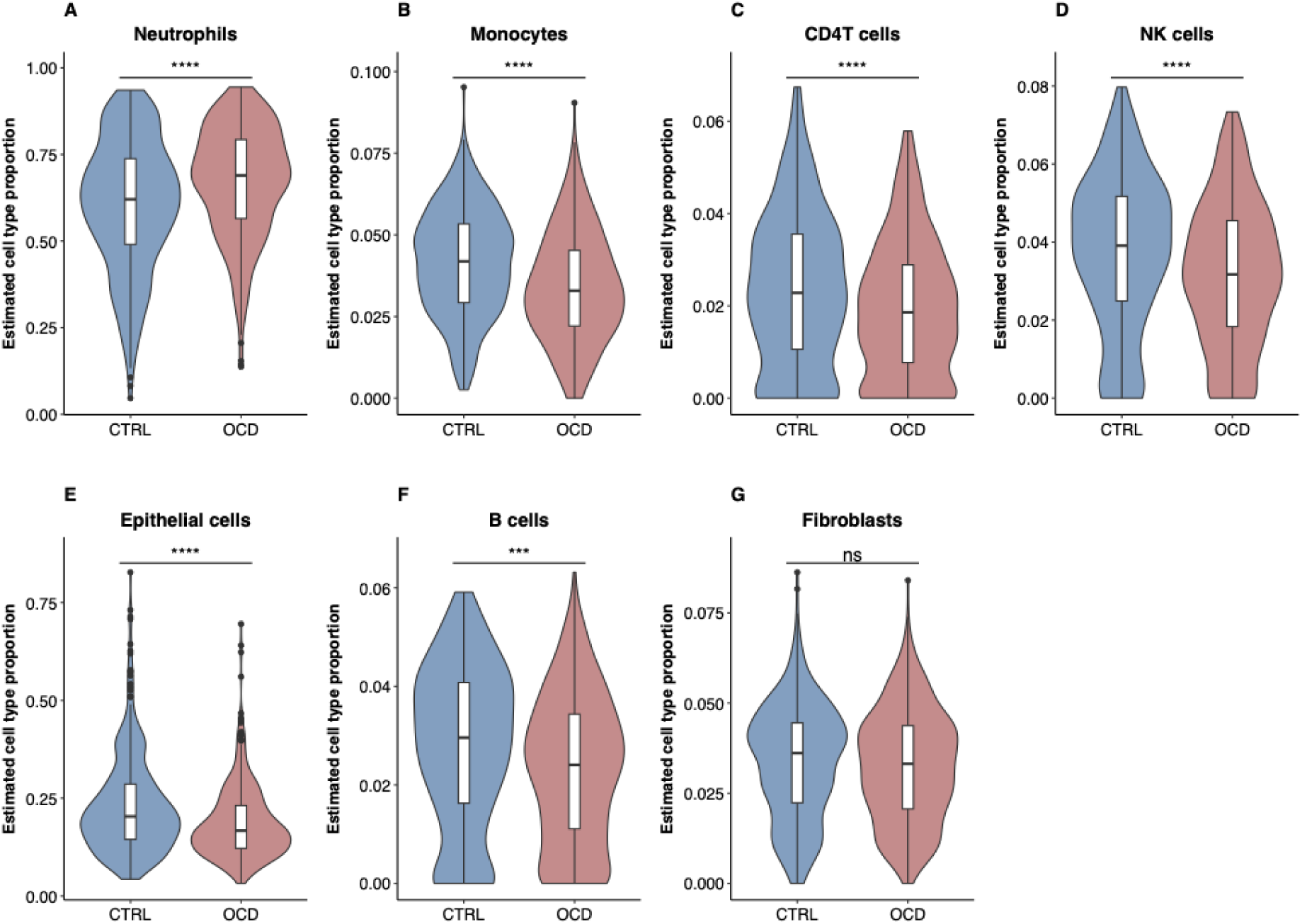
Differences in estimated saliva cell type proportions between individuals with OCD and healthy controls. NK: natural killer, OCD: obsessive-compulsive disorder

However, smoking had a stronger overall effect on estimated cell type proportions than OCD case status, which was accounted for in the analysis (**Table S11A**). The effect of OCD case status on cell type proportions did not differ between males and females (**Table S11B**).

To determine whether these associations were driven by psychoactive medication use (in about one third of OCD cases), we repeated the analysis including only unmedicated individuals with OCD. In this subset, all analyzed cell types remained significantly associated with OCD case status, and fibroblasts also reached significance (**Figure S6**, **Table S11C**), indicating that the observed differences are not attributable to medication use.

## Discussion

We report the largest methylome-wide association study (MWAS) of OCD to date, using saliva-derived DNA methylation profiles. We identified 35 differentially methylated positions (DMPs) and 17 differentially methylated regions (DMRs) associated with OCD, plus additional sex-specific DMRs. Integration with genetic variation revealed that some of these DNA methylation differences are genetically influenced, while others may reflect environmental influences or changes associated with having the disease status. Importantly, sex-specific analyses followed by a cross-sex meta-analysis uncovered patterns that would be missed by traditional sex-adjusted models, highlighting the importance of considering sex as a biological variable in epigenetic research. Sensitivity analyses indicated that the findings were not driven by psychiatric comorbidities or psychoactive medication use.

Our findings suggest that DNA methylation differences associated with OCD may be shaped by both genetic and non-genetic factors. mQTL analysis showed that nearly half of the DMPs and over half of the DMRs were influenced by nearby genetic variation. At several of these loci - annotated to *GABRB3, RIN1,* and *MUC2* - genetic variants were associated with both methylation levels and OCD diagnosis, suggesting that these variants may contribute to OCD risk by modulating DNA methylation and gene regulation. Although these mQTLs were not among the genome-wide significant hits in the most recent large-scale OCD GWAS^4^, the convergence of genetic and epigenetic signals highlights them as promising targets for further investigation and underscores the relevance of gene-epigenome interactions in OCD. Conversely, other methylation differences appear independent of genetic background, suggesting potential influences from environmental exposures, early developmental factors, or the disease state itself. These findings support a multifactorial model in which genetic, epigenetic, and environmental factors all contribute to OCD.

Sex-specific analyses identified one significant DMP and 14 DMRs in females, and seven DMRs in males. Most of the DMRs identified in males showed significant sex-by-diagnosis interactions, indicating that certain DNA methylation associations with OCD may be sex-specific. Sex-specific analyses identified several genes previously associated with OCD or related phenotypes in genetic and epigenetic studies, including *TEX26*^49^, *AKAP12*^50,51^, and *PIWIL1*^18^. In addition, the cross-sex meta-analysis identified a DMP on the X chromosome that would have been missed using traditional sex-adjusted models, which typically exclude sex chromosomes. Our study underscores the importance of incorporating sex as a biological variable in molecular research on psychiatric disorders. Sex-stratified approaches not only enhance signal detection but also uncover potentially distinct pathways of risk between the sexes.

Several differentially methylated loci mapped to genes relevant to OCD neurobiology. Of particular interest are *GABBR1* and *GABRB3*, encoding subunits of GABA receptors critical for inhibitory neurotransmission - a process thought to be reduced in OCD^52^. Genetic variants in *GABBR1* have been previously associated with OCD^53^ as well as related traits such as neuroticism and depression^54–56^, and there is preliminary evidence of differential methylation of this gene in OCD^17^, *GABRB3*, identified in the male-specific analysis, has also been associated with several psychiatric and neurodevelopmental conditions^57–59^.

We also observed methylation differences at a DMP annotated to *GPRIN3*, a regulator of dopaminergic signaling in the striatum, consistent with models of striatal dysfunction in OCD^60^. Additional differences were identified in genes related to neurodevelopment and neuroplasticity, including *BAI1 (ADGRB1), ARHGEF10, ZNRF1, ABLIM1, DYRK2*, *KIFC3*, *DCHS1, RIN1, PGBD5, RUNX3, and PIWIL1*. These genes are involved in axonal integrity, dendritic growth, myelination, and synaptic organization, supporting a neurodevelopmental basis for OCD. Several of the identified genes also play roles in gene regulation, such as *DAXX* and *DNMT3A*, previously associated with psychiatric^61–63^ and neurodevelopmental disorders^64^, respectively. *MIR29A* and *MIR21* encode microRNAs implicated in neuronal development and glial functioning^65–67^, respectively. Collectively, these findings align with current knowledge of molecular pathways involved in OCD.

Our findings also suggest immune involvement. Three DMRs (annotated to *GABBR1*, *DAXX*, and *B3GALT4*) mapped to the major histocompatibility complex (MHC) region on chromosome 6^68^, which contains numerous immune-related genes and was also implicated in the most recent large OCD GWAS^4^. Given the complex regulatory landscape of the MHC, these methylation signals may reflect effects on nearby immune genes. Additional differentially methylated loci were annotated to genes such as *CSF1*, *TRIM14*, *RUNX3*, *DLL1*, and *HIVEP3*, all known regulators of immune responses, and many of the associated loci showed immune-related GWAS hits in the PheWAS catalog^47^. Furthermore, we observed a higher proportion of estimated immune cells - particularly neutrophils - in OCD cases compared to controls, independent of smoking or medication use. These cell-type differences were statistically controlled for in all MWAS analyses to mitigate confounding. Taken together, these results align with growing evidence of immune dysregulation in OCD^69^, including altered inflammatory markers^70^, increased neuroinflammation^71^, and elevated rates of autoimmune comorbidities^69^.

This study was conducted using DNA extracted from saliva. While saliva is not the primary tissue of interest for mental disorders, peripheral tissues can still identify clinically useful biomarkers. Moreover, saliva may capture systemic processes such as immune or inflammatory activity, which are relevant to OCD. It may also capture DNA methylation patterns that reflect those in brain tissue. To assess this, we examined correlations between DNA methylation in saliva and brain using reference dataset^48^ While most DMPs showed low cross-tissue correlation, a substantial proportion of DMRs contained CpGs with moderate (|ρ| > 0.3) to high (|ρ| > 0.5) correlation. Three DMRs demonstrated strong and significant correlations across all their CpGs, annotated to *PGBD5 (females)*, *GABRB3* (males), and *PIWIL1;LOC101927786 (males)*. These stronger correlations observed at the regional level may reflect broader regulatory patterns conserved across tissues. However, these correlations should be interpreted with caution, particularly where correlations are low, as the reference dataset was derived from a limited number of brain samples spanning multiple regions^48^.

This study has several strengths: it is the largest MWAS of OCD, incorporated a sex-stratified design that enabled the identification of both shared and sex-specific signals, integrated genotype data, and applied rigorous quality control.

Limitations include: I) The statistical power remained limited for detecting the typically small effect sizes, particularly in the sex-specific analyses, which had smaller sample sizes. II) The study sample mostly comprised individuals of European ancestry, which may restrict generalizability. III) Although known confounders were accounted for, we cannot fully exclude the influence of unmeasured factors (e.g., sample timing, BMI) or residual confounding from medication use or comorbidities, despite sensitivity analyses showing no effects. IV) DNA extraction was conducted at two different sites, potentially introducing batch effects. However, case samples were distributed across both locations, control probe principal components were used to adjust for batch variation, and QQ plots showed no inflation - suggesting minimal impact. V) Cell type proportions were estimated rather than directly measured; however, prior research has demonstrated a strong correlation (r = 0.72) between estimated saliva and buccal cell proportions and cytological staining^72^, supporting the reliability of these estimates.

Thus, our findings should be replicated in independent datasets, ideally including diverse populations and combined with functional studies to determine how the identified DNA methylation differences influence gene expression. Longitudinal studies with repeated measurements could help clarify the temporal dynamics of these DNA methylation marks, including whether they change in response to therapeutic interventions.

In conclusion, we identified differential DNA methylation associated with OCD, including both shared and sex-specific signals. These differences involve genes related to neurotransmission, synaptic plasticity, neurodevelopment, gene regulation, and immune function, and appear to be influenced by both genetic and non-genetic factors. Our findings support a multifactorial model of OCD, to which a range of factors likely contribute. As more datasets become available, the identification of OCD risk profiles that integrate genetic, epigenetic, and environmental factors could help advance clinically relevant tools for improved risk stratification.

## Data Availability

All scripts are available on GitHub (https://github.com/KiraHoeffler/OCD_MWAS). Due to ethical restrictions and data privacy regulations, individual-level data cannot be shared.

## Acknowledgements

We are deeply grateful to all study participants for their contribution. We thank Helene Nilsen, Marie Bjorøy, and the participating psychotherapists for their support in sample and data collection. We are also grateful to Louise Grevle and Lisa Vårdal for sample preparation and coordinating sample shipments, and we acknowledge the contribution of the biobank infrastructure maintained by Per Morten Knappskog, Stefan Johansson, and their team. We further thank Nadine Fricker at Life & Brain for excellent communication and service and Lea Zillich for her valuable advice. This research was made possible by the generous financial support of the Trond Mohn Foundation. KDH received a fellowship from the University of Bergen, and AKS and SLH were supported by a Bedrehelse grant (#273446). The collection of control samples was funded by grants from the Research Council of Norway and Helse Vest. Grammatical revisions to this manuscript were assisted by GPT-4o (OpenAI, San Francisco, United States).

## Author contributions

Study design: KDH, SLH

Data collection: SLH, GK, BH, TOE, KH, JJC, JH, NORDiC

Data analysis: KDH, AKS, MWH, AH

Data interpretation: KDH, SLH, AKS, TK, KJR

Manuscript writing: KDH, SLH and TK

All authors read, edited and approved the manuscript.

## Nordic OCD and Related Disorders Consortium (NORDiC)

Julia Bäckman^12^, Long Long Chen^12^, James J. Crowley^5,11^, Elles de Schipper^12^, Diana Pascal^12^, Jan Haavik^2,9^, Kristen Hagen^2,6,8^, Matthew W. Halvorsen^5^, Bjarne Hansen^2,7^, Kira D. Höffler^1,2,3,4^, Fred Johansson^12,13^, Elinor K. Karlsson^14,15^, Gerd Kvale^10^, Paul Lichtenstein^16^, Kerstin Lindblad-Toh^15,17^, Manuel Mattheisen^18-21^, David Mataix-Cols^12^, Christian Rück^12^, Thorstein Olsen Eide^2,6,7^, Nora I. Strom^12,18,19,22,23^, John Wallert^12^

^12^Centre for Psychiatry Research, Department of Clinical Neuroscience, Karolinska Institutet and Stockholm Health Care Services, Region Stockholm, Stockholm, Sweden.

^13^Department of Health Promoting Science, Sophiahemmet University, Stockholm, Sweden.

^14^Department of Bioinformatics and Integrative Biology, University of Massachusetts Medical School, Worcester, MA, USA.

^15^Broad Institute of MIT and Harvard, Cambridge, MA, USA.

^16^Department of Medical Epidemiology and Biostatistics, Karolinska Institutet, Stockholm, Sweden.

^17^Science for Life Laboratory, Department of Medical Biochemistry and Microbiology, Uppsala University, 751 32, Uppsala, Sweden.

^18^Institute of Psychiatric Phenomics and Genomics (IPPG), University Hospital, LMU Munich, Munich, Germany.

^19^Department of Biomedicine, Aarhus University, Aarhus, Denmark.

^20^Department of Community Health and Epidemiology, Dalhousie University, Halifax, Nova Scotia, Canada.

^21^Faculty of Computer Science, Dalhousie University, Halifax, Nova Scotia, Canada.

^22^Department of Psychology, Humboldt-Universität zu Berlin, Berlin, Germany. ^23^University Hospital of Psychiatry and Psychotherapy, University of Bern, Bern, Switzerland.

## References

1. Ruscio, A. M., Stein, D. J., Chiu, W. T. & Kessler, R. C. The Epidemiology of Obsessive-Compulsive Disorder in the National Comorbidity Survey Replication. Molecular psychiatry 15, 53 (2010).

2. Stein, D. J. et al. Obsessive-compulsive disorder. Nat Rev Dis Primers 5, 52 (2019).

3. Mataix-Cols, D. et al. Heritability of Clinically Diagnosed Obsessive-Compulsive Disorder Among Twins. JAMA Psychiatry 81, 631–632 (2024).

4. Strom, N. I. et al. Genome-wide analyses identify 30 loci associated with obsessive– compulsive disorder. Nat Genet 1–13 (2025) doi:10.1038/s41588-025-02189-z.

5. Brander, G., Pérez-Vigil, A., Larsson, H. & Mataix-Cols, D. Systematic review of environmental risk factors for Obsessive-Compulsive Disorder: A proposed roadmap from association to causation. Neurosci Biobehav Rev 65, 36–62 (2016).

6. Cappi, C. et al. Epigenetic evidence for involvement of the oxytocin receptor gene in obsessive-compulsive disorder. BMC Neurosci 17, 79 (2016).

7. D’Addario, C. et al. Regulation of oxytocin receptor gene expression in obsessive– compulsive disorder: a possible role for the microbiota-host epigenetic axis. Clin Epigenetics 14, 47 (2022).

8. Park, C. I., Kim, H. W., Jeon, S., Kang, J. I. & Kim, S. J. Reduced DNA methylation of the oxytocin receptor gene is associated with obsessive-compulsive disorder. Clin Epigenetics 12, 101 (2020).

9. Schiele, M. A. et al. Oxytocin Receptor Gene DNA Methylation: A Biomarker of Treatment Response in Obsessive-Compulsive Disorder? Psychother Psychosom 90, 57–63 (2021).

10. Seo, J. H., Kim, S. T., Jeon, S., Kang, J. I. & Kim, S. J. Sex-dependent association of DNA methylation of HPA axis-related gene FKBP5 with obsessive-compulsive disorder. Psychoneuroendocrinology 158, 106404 (2023).

11. D’Addario, C. et al. Exploring the role of BDNF DNA methylation and hydroxymethylation in patients with obsessive compulsive disorder. J Psychiatr Res 114, 17–23 (2019).

12. D’Addario, C. et al. In Search for Biomarkers in Obsessive-Compulsive Disorder: New Evidence on Saliva as a Practical Source of DNA to Assess Epigenetic Regulation. Curr Med Chem 29, 5782–5791 (2022).

13. Campos-Martin, R. et al. Epigenome-wide analysis identifies methylome profiles linked to obsessive-compulsive disorder, disease severity, and treatment response. Mol Psychiatry 28, 4321–4330 (2023).

14. de Oliveira, K. C. et al. Brain areas involved with obsessive-compulsive disorder present different DNA methylation modulation. BMC Genomic Data 22, 45 (2021).

15. Goodman, S. J. et al. Obsessive-compulsive disorder and attention-deficit/hyperactivity disorder: distinct associations with DNA methylation and genetic variation. Journal of Neurodevelopmental Disorders 12, 23 (2020).

16. Guo, L. et al. Epigenome-wide DNA methylation analysis of whole blood cells derived from patients with GAD and OCD in the Chinese Han population. Transl Psychiatry 12, 465 (2022).

17. Nissen, J. B. et al. DNA Methylation at the Neonatal State and at the Time of Diagnosis: Preliminary Support for an Association with the Estrogen Receptor 1, Gamma-Aminobutyric Acid B Receptor 1, and Myelin Oligodendrocyte Glycoprotein in Female Adolescent Patients with OCD. Front Psychiatry 7, 35 (2016).

18. Rodriguez, N. et al. Integrative DNA Methylation and Gene Expression Analysis of Cognitive Behavioral Therapy Response in Children and Adolescents with Obsessive-Compulsive Disorder; a Pilot Study. Pharmgenomics Pers Med 14, 757–766 (2021).

19. Schiele, M. A. et al. Epigenome-wide DNA methylation in obsessive-compulsive disorder. Transl Psychiatry 12, 1–7 (2022).

20. Yue, W. et al. Genome-wide DNA methylation analysis in obsessive-compulsive disorder patients. Sci Rep 6, 31333 (2016).

21. Mathes, B. M., Morabito, D. M. & Schmidt, N. B. Epidemiological and Clinical Gender Differences in OCD. Curr Psychiatry Rep 21, 36 (2019).

22. Mathis, M. A. de et al. Gender differences in obsessive-compulsive disorder: a literature review. Braz J Psychiatry 33, 390–399 (2011).

23. American Psychiatric Association. DSM-5: Diagnostic And Statistical Manual Of Mental Disorders. (American Psychiatric Publishing, Washington, D.C, 2013).

24. Sheehan, D. V. et al. The Mini-International Neuropsychiatric Interview (M.I.N.I.): the development and validation of a structured diagnostic psychiatric interview for DSM-IV and ICD-10. J Clin Psychiatry 59 Suppl 20, 22-33;quiz 34–57 (1998).

25. Goodman, W. K. et al. The Yale-Brown Obsessive Compulsive Scale: I. Development, use, and reliability. Archives of General Psychiatry 46, 1006–1011 (1989).

26. Johansson, S. et al. Genetic analyses of dopamine related genes in adult ADHD patients suggest an association with the DRD5-microsatellite repeat, but not with DRD4 or SLC6A3 VNTRs. Am J Med Genet B Neuropsychiatr Genet 147B, 1470–1475 (2008).

27. Irgens, L. M. The Medical Birth Registry of Norway. Epidemiological research and surveillance throughout 30 years. Acta Obstetricia et Gynecologica Scandinavica 79, 435–439 (2000).

28. Wickham, H. Ggplot2: Elegant Graphics for Data Analysis. (Springer, New York, NY, 2009). doi:10.1007/978-0-387-98141-3.

29. Lehne, B. et al. A coherent approach for analysis of the Illumina HumanMethylation450 BeadChip improves data quality and performance in epigenome-wide association studies. Genome Biol 16, 37 (2015).

30. Ritchie, M. E. et al. limma powers differential expression analyses for RNA-sequencing and microarray studies. Nucleic Acids Res 43, e47 (2015).

31. Du, P. et al. Comparison of Beta-value and M-value methods for quantifying methylation levels by microarray analysis. BMC Bioinformatics 11, 587 (2010).

32. Philibert, R., Dogan, M., Beach, S. R. H., Mills, J. A. & Long, J. D. AHRR methylation predicts smoking status and smoking intensity in both saliva and blood DNA. Am J Med Genet B Neuropsychiatr Genet 183, 51–60 (2020).

33. Dawes, K. et al. The relationship of smoking to cg05575921 methylation in blood and saliva DNA samples from several studies. Sci Rep 11, 21627 (2021).

34. Höffler, K. D., et al. Optimizing Genetic Ancestry Adjustment in DNA Methylation Studies: A Comparative Analysis of Approaches. Preprint at 10.21203/rs.3.rs-6580295/v1 (2025).

35. Turner, S. D. qqman: an R package for visualizing GWAS results using Q-Q and manhattan plots. Journal of Open Source Software 3, 731 (2018).

36. Fox, J. & Weisberg, S. An R Companion to Applied Regression. (Sage, Thousand Oaks CA, 2019).

37. van Iterson, M., van Zwet, E. W., BIOS Consortium & Heijmans, B. T. Controlling bias and inflation in epigenome- and transcriptome-wide association studies using the empirical null distribution. Genome Biol 18, 19 (2017).

38. Tesfaye, M. et al. Sex effects on DNA methylation affect discovery in epigenome-wide association study of schizophrenia. Mol Psychiatry 1–11 (2024) doi:10.1038/s41380-024-02513-9.

39. Schwarzer, G., Carpenter, J. R. & Rücker, G. Meta-Analysis with R. (Springer International Publishing, Cham, 2015). doi:10.1007/978-3-319-21416-0.

40. Xu, Z., Niu, L. & Taylor, J. A. The ENmix DNA methylation analysis pipeline for Illumina BeadChip and comparisons with seven other preprocessing pipelines. Clinical Epigenetics 13, 216 (2021).

41. Pedersen, B. S., Schwartz, D. A., Yang, I. V. & Kechris, K. J. Comb-p: software for combining, analyzing, grouping and correcting spatially correlated P-values. Bioinformatics 28, 2986–2988 (2012).

42. Mallik, S. et al. An evaluation of supervised methods for identifying differentially methylated regions in Illumina methylation arrays. Brief Bioinform 20, 2224–2235 (2019).

43. Phipson, B., Maksimovic, J. & Oshlack, A. missMethyl: an R package for analyzing data from Illumina’s HumanMethylation450 platform. Bioinformatics 32, 286–288 (2016).

44. Li, J. & Ji, L. Adjusting multiple testing in multilocus analyses using the eigenvalues of a correlation matrix. Heredity 95, 221–227 (2005).

45. 1000 Genomes Project Consortium et al. A global reference for human genetic variation. Nature 526, 68–74 (2015).

46. Halvorsen, M. W. et al. A burden of rare copy number variants in obsessive-compulsive disorder. Mol Psychiatry 30, 1510–1517 (2025).

47. Watanabe, K. et al. A global overview of pleiotropy and genetic architecture in complex traits. Nat Genet 51, 1339–1348 (2019).

48. Braun, P. R. et al. Genome-wide DNA methylation comparison between live human brain and peripheral tissues within individuals. Transl Psychiatry 9, 47 (2019).

49. Strom, N. I. et al. Genome-wide association study identifies new locus associated with OCD. 2021.10.13.21261078 Preprint at 10.1101/2021.10.13.21261078 (2021).

50. Lin, G. N. et al. De novo mutations identified by whole-genome sequencing implicate chromatin modifications in obsessive-compulsive disorder. Science Advances 8, eabi6180 (2022).

51. Zhai, R. et al. Rhesus monkeys exhibiting spontaneous ritualistic behaviors resembling obsessive-compulsive disorder. Natl Sci Rev 10, nwad312 (2023).

52. Richter, M. A. et al. Evidence for cortical inhibitory and excitatory dysfunction in obsessive compulsive disorder. Neuropsychopharmacology 37, 1144–1151 (2012).

53. Zai, G. et al. Evidence for the gamma-amino-butyric acid type B receptor 1 (GABBR1) gene as a susceptibility factor in obsessive-compulsive disorder. Am J Med Genet B Neuropsychiatr Genet 134B, 25–29 (2005).

54. Nagel, M. et al. Meta-analysis of genome-wide association studies for neuroticism in 449,484 individuals identifies novel genetic loci and pathways. Nat Genet 50, 920–927 (2018).

55. Luciano, M. et al. Association analysis in over 329,000 individuals identifies 116 independent variants influencing neuroticism. Nat Genet 50, 6–11 (2018).

56. Fatemi, S. H., Folsom, T. D. & Thuras, P. D. Deficits in GABA(B) receptor system in schizophrenia and mood disorders: a postmortem study. Schizophr Res 128, 37–43 (2011).

57. Buxbaum, J. D. et al. Association between a GABRB3 polymorphism and autism. Mol Psychiatry 7, 311–316 (2002).

58. Hodges, L. M. et al. Evidence for linkage and association of GABRB3 and GABRA5 to panic disorder. Neuropsychopharmacology 39, 2423–2431 (2014).

59. Johannesen, K. M. et al. Structural mapping of GABRB3 variants reveals genotype-phenotype correlations. Genet Med 24, 681–693 (2022).

60. Burguière, E., Monteiro, P., Mallet, L., Feng, G. & Graybiel, A. M. Striatal circuits, habits, and implications for obsessive-compulsive disorder. Curr Opin Neurobiol 0, 59–65 (2015).

61. Strom, N., I., et al. Genome-wide association study of major anxiety disorders in 122,341 European-ancestry cases identifies 58 loci and highlights GABAergic signaling. medRxiv 2024.07.03.24309466 (2024) doi:10.1101/2024.07.03.24309466.

62. Sheng, G., Demers, M., Subburaju, S. & Benes, F. M. Differences in the circuitry-based association of copy numbers and gene expression between the hippocampi of patients with schizophrenia and the hippocampi of patients with bipolar disorder. Arch Gen Psychiatry 69, 550–561 (2012).

63. Ruzicka, W. B., Subburaju, S. & Benes, F. M. Circuit- and Diagnosis-Specific DNA Methylation Changes at γ-Aminobutyric Acid-Related Genes in Postmortem Human Hippocampus in Schizophrenia and Bipolar Disorder. JAMA Psychiatry 72, 541–551 (2015).

64. Christian, D. L. et al. DNMT3A Haploinsufficiency Results in Behavioral Deficits and Global Epigenomic Dysregulation Shared across Neurodevelopmental Disorders. Cell Rep 33, 108416 (2020).

65. Lippi, G. et al. Targeting of the Arpc3 actin nucleation factor by miR-29a/b regulates dendritic spine morphology. J Cell Biol 194, 889–904 (2011).

66. Duan, P. et al. miR-29a modulates neuronal differentiation through targeting REST in mesenchymal stem cells. PLoS One 9, e97684 (2014).

67. Miguel-Hidalgo, J. J. et al. MicroRNA-21: expression in oligodendrocytes and correlation with low myelin mRNAs in depression and alcoholism. Prog Neuropsychopharmacol Biol Psychiatry 79, 503–514 (2017).

68. Horton, R. et al. Gene map of the extended human MHC. Nat Rev Genet 5, 889– 899 (2004).

69. Endres, D. et al. Immunological causes of obsessive-compulsive disorder: is it time for the concept of an ‘autoimmune OCD’ subtype? Transl Psychiatry 12, 5 (2022).

70. Gray, S. M. & Bloch, M. H. Systematic review of proinflammatory cytokines in obsessive-compulsive disorder. Curr Psychiatry Rep 14, 220–228 (2012).

71. Attwells, S. et al. Inflammation in the Neurocircuitry of Obsessive-Compulsive Disorder. JAMA Psychiatry 74, 833–840 (2017).

72. Wong, Y. T. et al. A comparison of epithelial cell content of oral samples estimated using cytology and DNA methylation. Epigenetics 17, 327–334 (2022).

