## Supplementary Materials for "Methylome-Wide Association Study of Obsessive-Compulsive Disorder"

#### Authors

Kira D. Höffler[1,2,3,4*], Anne-Kristin Stavrum[1,2,3], Matthew W. Halvorsen[5], Thorstein Olsen Eide[2,6,7], Kristen Hagen[2,6,8], André Høberg[9], Nordic OCD and Related Disorders Consortium (NORDiC), Gerd Kvale[2,10], James J. Crowley[5,11], Jan Haavik[2,9], Kerry J. Ressler[4], Bjarne Hansen[2,7], Torsten Klengel[4], Stephanie Le Hellard[1,2,3*]

<u>Nordic OCD and Related Disorders Consortium (NORDiC)</u>

Julia Bäckman[12], Long Long Chen[12], James J. Crowley[5,11], Elles de Schipper[12], Diana Pascal[12], Jan Haavik[2,9], Kristen Hagen[2,6,8], Matthew W. Halvorsen[5], Bjarne Hansen[2,7], Kira D. Höffler[1,2,3,4], Fred Johansson[12,13], Elinor K. Karlsson[14,15], Gerd Kvale[10], Paul Lichtenstein[16], Kerstin Lindblad-Toh[15,17], Manuel Mattheisen[18-21], David Mataix-Cols[12], Christian Rück[12], Thorstein Olsen Eide[2,6,7], Nora I. Strom[12,18,19,22,23], John Wallert[12]

#### Affiliations

[1]Department for Clinical Medicine, University of Bergen, Bergen, Norway

[2]Bergen Center for Brain Plasticity, Haukeland University Hospital, Bergen, Norway

[3]Dr. Einar Martens Research Group for Biological Psychiatry, Department of Medical Genetics, Haukeland University Hospital, Bergen, Norway

[4]Department of Psychiatry, McLean Hospital, Harvard Medical School, Belmont, MA, USA

[5]Department of Genetics, University of North Carolina at Chapel Hill, Chapel Hill, NC, USA

[6]Department of Psychiatry, Møre og Romsdal Hospital Trust, Molde, Norway

[7]Center for Crisis Psychology, University of Bergen, Bergen, Norway

[8]Department of Mental Health, Norwegian University of Science and Technology, Trondheim, Norway
[9]Department of Biomedicine, University of Bergen, Bergen, Norway
[10]Department of Clinical Psychology, University of Bergen, Norway
[11]Department of Psychiatry, University of North Carolina at Chapel Hill, Chapel Hill, NC, USA
[12]Centre for Psychiatry Research, Department of Clinical Neuroscience, Karolinska Institutet and Stockholm Health Care Services, Region Stockholm, Stockholm, Sweden.
[13]Department of Health Promoting Science, Sophiahemmet University, Stockholm, Sweden.
[14]Department of Bioinformatics and Integrative Biology, University of Massachusetts Medical School, Worcester, MA, USA.
[15]Broad Institute of MIT and Harvard, Cambridge, MA, USA.
[16]Department of Medical Epidemiology and Biostatistics, Karolinska Institutet, Stockholm, Sweden.
[17]Science for Life Laboratory, Department of Medical Biochemistry and Microbiology, Uppsala University, 751 32, Uppsala, Sweden.
[18]Institute of Psychiatric Phenomics and Genomics (IPPG), University Hospital, LMU Munich, Munich, Germany.
[19]Department of Biomedicine, Aarhus University, Aarhus, Denmark.
[20]Department of Community Health and Epidemiology, Dalhousie University, Halifax, Nova Scotia, Canada.
[21]Faculty of Computer Science, Dalhousie University, Halifax, Nova Scotia, Canada.
[22]Department of Psychology, Humboldt-Universität zu Berlin, Berlin, Germany.
[23]University Hospital of Psychiatry and Psychotherapy, University of Bern, Bern, Switzerland.

### Supplementary Methods

#### Quality Control

The RGset was background-corrected using the bgcorrect.illumina() function from the *minfi*[1] package. Filtering, normalization, and imputation were performed separately for autosomes, the X chromosome in females, and XY chromosomes in males. We applied a detection p value threshold of 10E-16[2] for autosomes and the X chromosome in females, and 10E-5 for XY chromosomes in males (less strict due to the haploid nature of the sex chromosomes in males).

Probes were excluded if they:

1) had low bead counts (>5% of samples with counts <3)
2) had more than 5% missing data
3) were cross-reactive probes
4) were SNP (single nucleotide polymorphism) probes (up to 10 bp from the CpG site and MAF of 0.01)[3,4]
5) were flagged probes or probes with mapping inaccuracies in Illumina product files).

A total of 805,163 CpG probes remained after quality control.

Samples were excluded if they:

1) had sex mismatches between reported and sex predicted from DNA methylation data using the getSex() function from the minfi package[1]
2) were outliers in the minfi sex plot[1]
3) had genotype mismatches between repeated samples[5]
4) had low bisulfite conversion rates (<80%)[6]
5) had more than 5% missing data
6) were clear outliers in the beta value distributions

Quantile normalization was applied to the filtered intensity values, followed by BMIQ[7] normalization on beta values for autosomes. BMIQ was not applied to the sex chromosomes, as it assumes a beta value distribution that does not fit these chromosomes. K-nearest neighbor imputation[8] was used for missing values.

Replicated probes were handled similarly to the rmPosReps() function of the DMRcate[9] package, based on Peters et al. (2024)[4], with precision prioritized over sensitivity and weighted averages instead of standard averages calculated. CpGs with overlapping SNPs and MAF 0.05 were quality-controlled similarly to sex chromosomes in females and later used for genetic ancestry adjustment, as described elsewhere[10]. For detailed information, please refer to the uploaded scripts on GitHub (https://github.com/KiraHoeffler/OCD_MWAS).

**Estimation of Cell Type Proportions**

Saliva cell type proportions were estimated using HEpiDISH[11] with the RPC method. The *centEpiFibFatIC.m* dataset (excluding fat) served as the primary reference, while the *centBloodSub.m* dataset was used as the secondary reference[12].

**Annotation**

As 65.3% of CpG sites on the EPICv2 array were not annotated in the Illumina manifest, CpG sites were reannotated in a stepwise manner, proceeding to the next step only if no annotation was obtained in the previous one:

1. Regulatory Region Proximity: CpGs were first annotated to regulatory regions and their respective genes based on ENCODE data (downloaded from https://screen.wenglab.org/downloads, April 2025)[13].

2. Proximity to TSS or Exon 1: CpGs located within 2,000 base pairs upstream of a gene's TSS or within exon 1 were annotated to that gene using RefSeq gene annotations obtained from the UCSC Genome Browser[14].

3. Gene Body Location: CpGs located within the gene body were annotated using RefSeq gene annotations (Perez et al., 2025)[14].

CpG island annotations were extracted from the Illumina EPIC v2 manifest. The adapted annotation, with genes annotated for 87.6% of CpGs, is provided in **Supplementary File 1**.

#### Calculation of Odds Ratios (ORs)

Individuals were categorized into the top 20% and bottom 20% of average DNA methylation levels at each DMR. Fisher's exact test was used to compute ORs and their p values.

#### DNA Methylation Values for Result Visualization

For result visualization, M-values were adjusted for age, cell type proportions, smoking, sex (for the cross-sex meta-analysis), and the same number of control probe and ancestry PCs used in the main model, using the removeBatchEffect() function from the limma package[15]. The adjusted M-values were then converted to beta values for plotting.

#### Sensitivity Analyses

In subset analyses - including non-medicated individuals and those without psychiatric comorbidities - the associations between average M-values across CpGs within DMRs, as well as M-values of DMPs, and OCD case-control status were evaluated using standard linear models, adjusting for the same covariates as in the main analysis.

#### Methylation Quantitative Trait Loci (mQTL) Analyses

The liftover tool[16] was used to convert coordinates from hg19 to hg38, samples were filtered based on available genotyping data, and SNPs were filtered on availability in all included datasets. For each DMP and DMR, SNPs located within 50,000 base pairs upstream and downstream - as well as within the DMR itself, when applicable - were selected. Association testing between each SNP and the average M values of the corresponding DMR was performed using a linear model, incorporating the same covariates as the main MWAS model but adjusting for ten genotyping PCs instead of two ancestry PCs. To identify significant mQTLs, FDR correction was applied across all SNPs from all loci. Case-control status was tested for association with each significant mQTL using a logistic regression model, adjusting for sex, age, and ten genotyping PCs.

To identify independent mQTLs, linkage disequilibrium (LD) pruning was performed using PLINK[17] with an LD $R^2$ threshold of 0.2, and a window size of 250 kilobases. From each LD block, the mQTL with the lowest p value for association with case-control status was selected. FDR correction was applied to the case-control association p values to account for the number of independent mQTLs tested.

SNPs in LD ($R^2 \geq 0.7$ within a 500 kb window) with mQTL SNPs that were significantly associated with OCD case-control status after correction for multiple testing were identified using PLINK[17]. We then extracted association results for these LD-SNPs to determine whether they showed similar association patterns as the lead SNPs.

##### GWAS Associations

For genes annotated to the identified DMPs and DMRs, significant GWAS hits (p value ≤ 5E-08) were identified using the PheWAS tool from the Atlas of GWAS Summary Statistics, release 3[18], including GWAS results from before 2020.

##### Saliva-Brain Correlation

Spearman's rank correlation coefficients (rho) and p-values for brain–saliva tissue pairs were obtained from data provided by Dr. Shizhong Han, based on the study by Braun et al. (2019)[19] for the DMPs and the CpGs within the identified DMRs.

#### Supplementary Figures

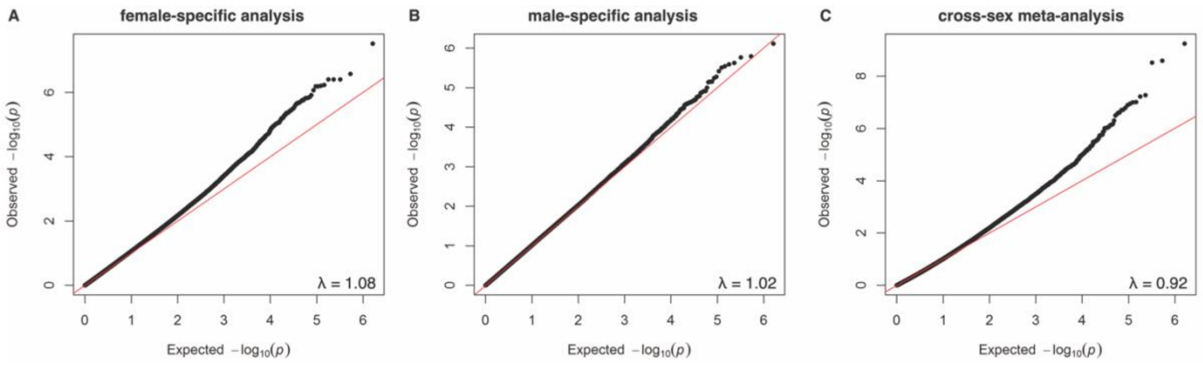


**Figure S1: Quantile-quantile plots.**

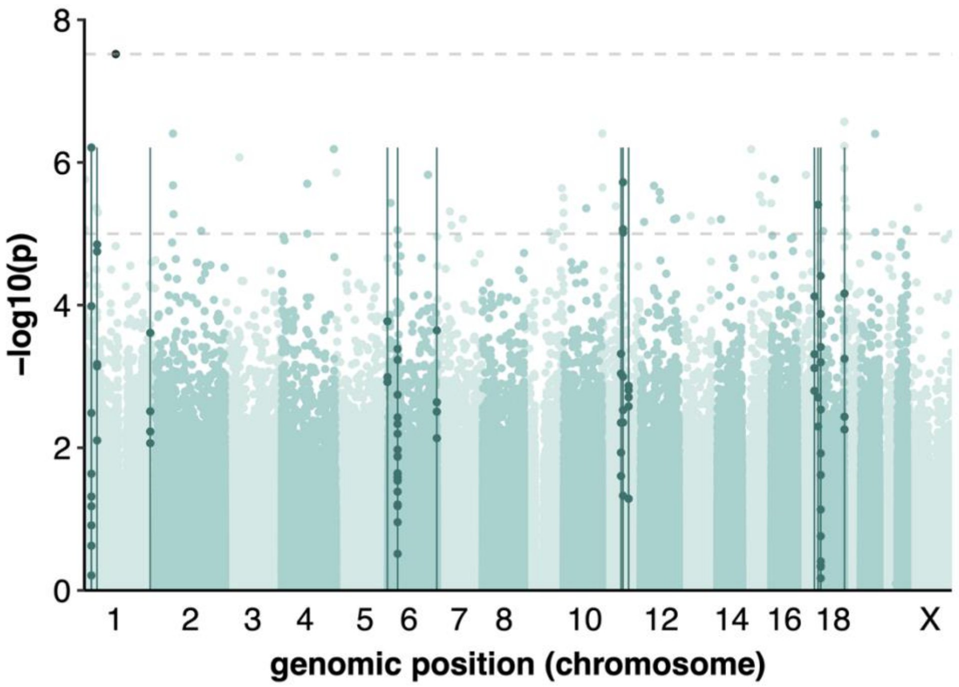


**Figure S2: Manhattan plot of the sex-specific analysis in females.** Differentially methylated positions (dots above the upper dashed line) and regions (vertical lines) are highlighted in dark green.

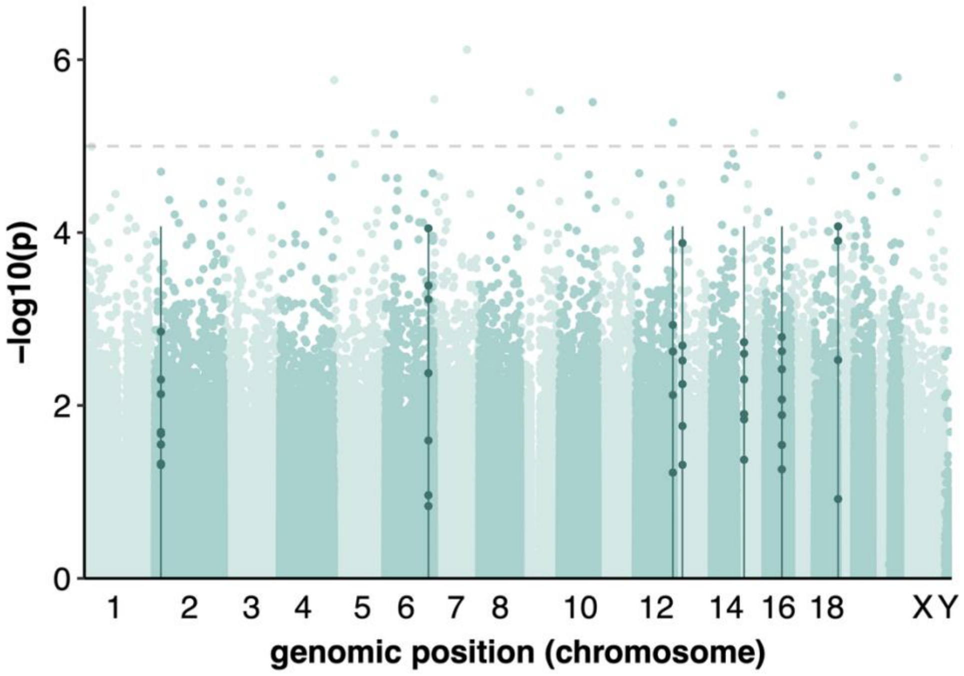


**Figure S3: Manhattan plot of the sex-specific analysis in males.** Differentially methylated positions (dots above the upper dashed line) and regions (vertical lines) are highlighted in dark green.

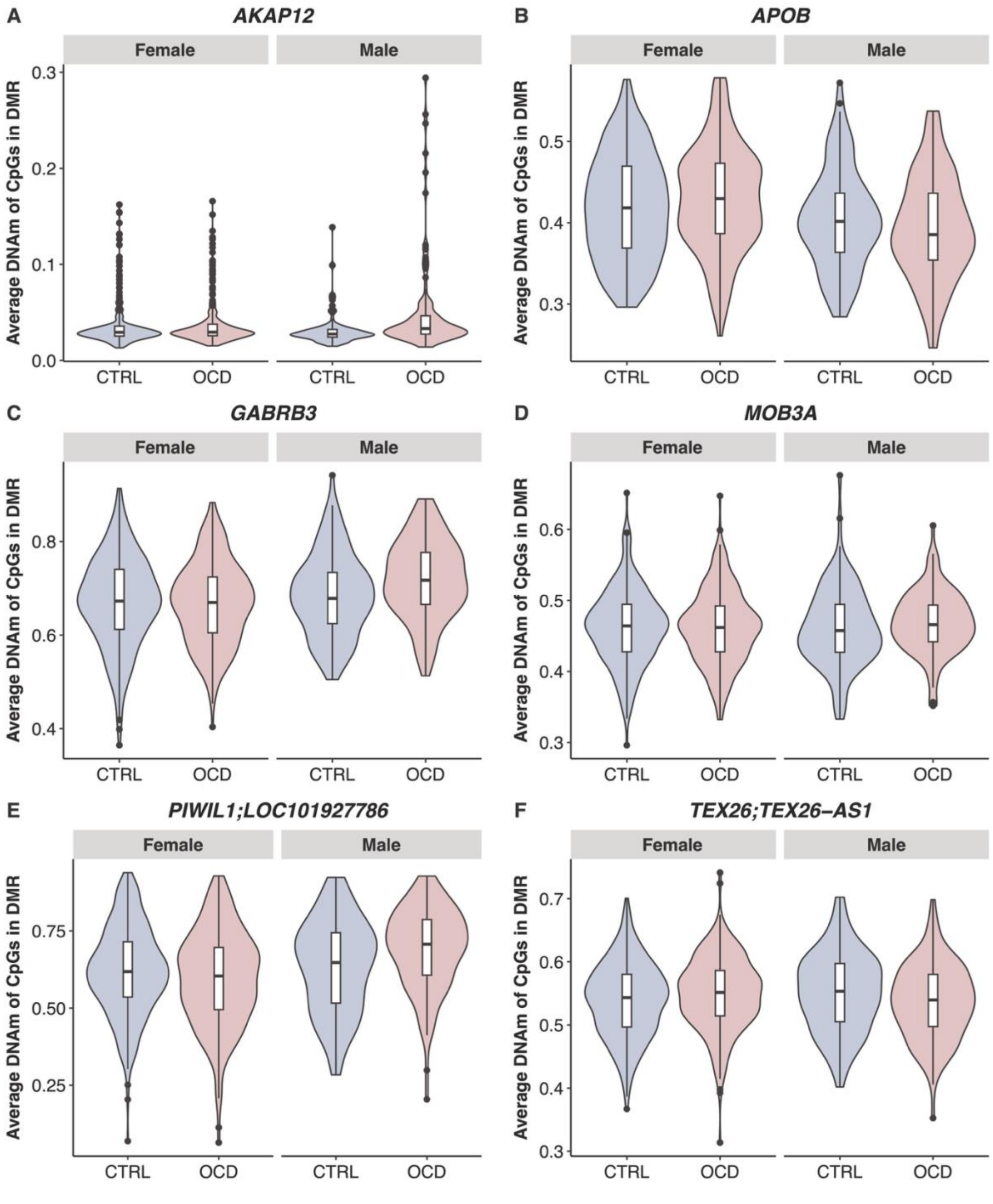


**Figure S4: DMRs with significant interaction between sex and the DNA methylation-response relationship.** CTRL: healthy control, DMR: differentially methylated region, DNAm: DNA methylation, OCD: obsessive-compulsive disorder

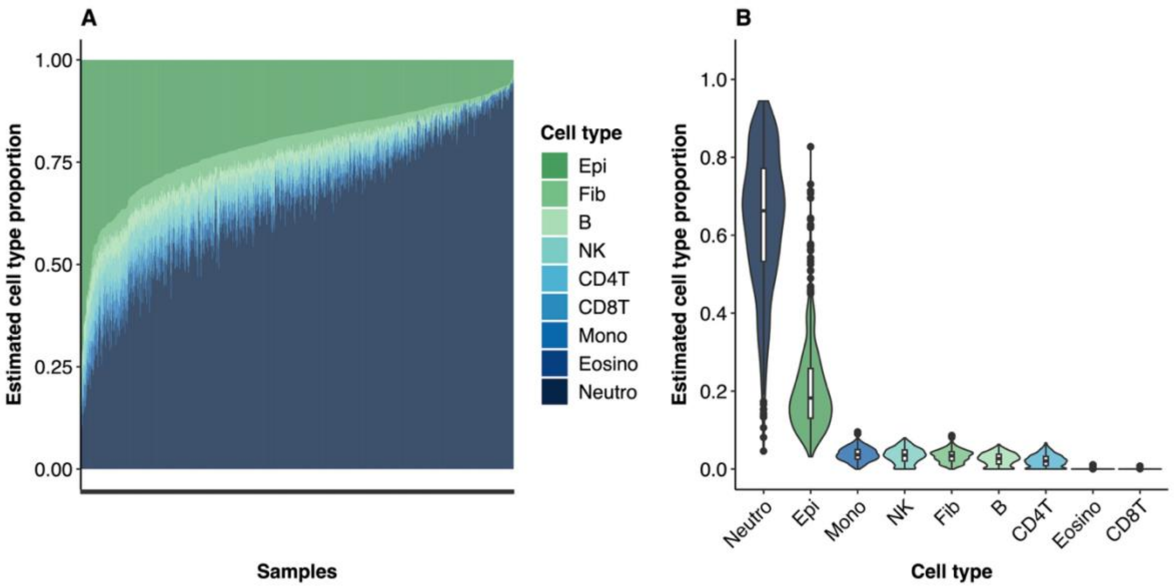


**Figure S5: Variability in estimated saliva cell type proportions across samples.** (A) Stacked bar plot showing the estimated cell type proportions for each sample. (B) Violin plot with overlaid box plots displaying the distribution of estimated cell type proportions for each cell type. Cell type proportions were estimated using HepiDish[11]. B: B cells, CD4T: CD4+ T lymphocytes, CD8T: CD8+ T lymphocytes, Epi: epithelial cells, Eosino: eosinophiles, Fib: fibroblasts, T lymphocytes, Epi: epithelial cells, Eosino: eosinophiles, Fib: fibroblasts, Mono: monocytes, Neutro: neutrophiles, NK: natural killer cells.

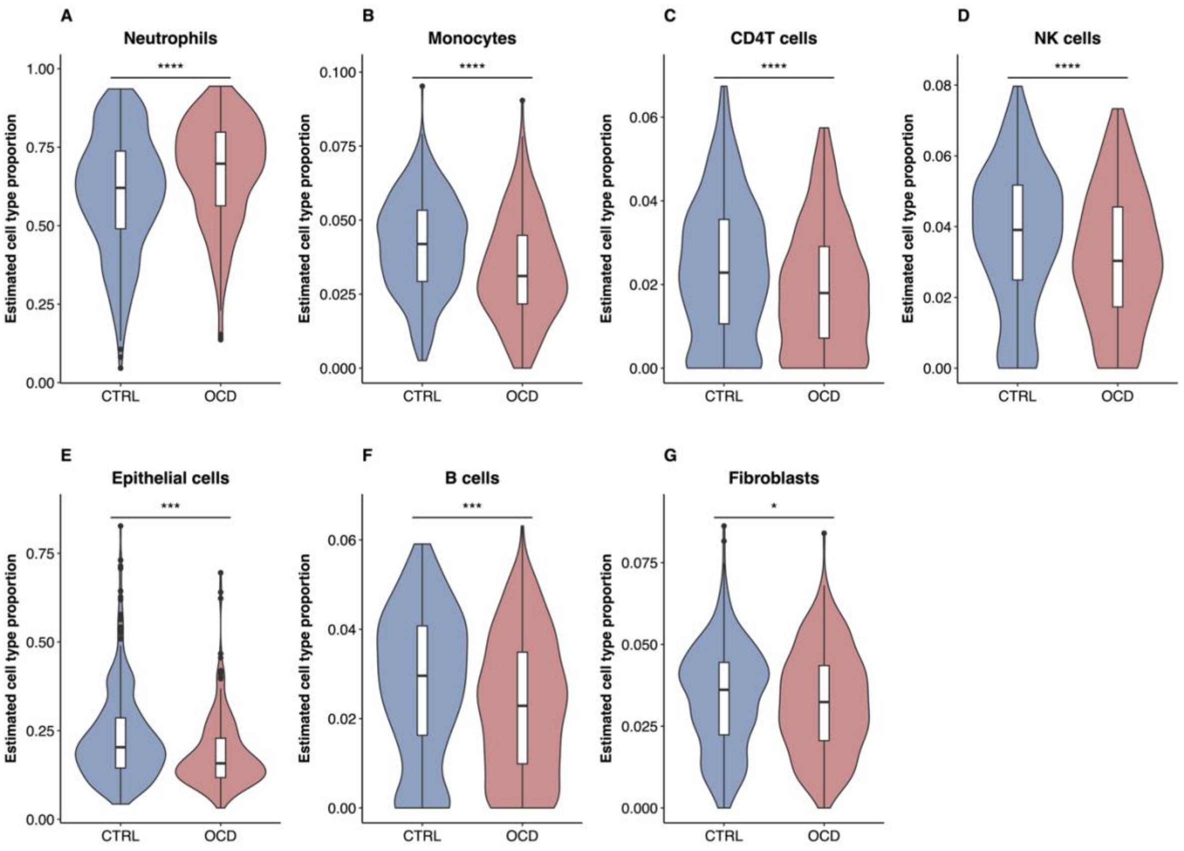


**Figure S6: Differences in estimated saliva cell type proportions between individuals with OCD who did not take psychoactive medication and healthy controls.** NK: natural killer, OCD: obsessive-compulsive disorder

### Supplementary Tables

**Table S1: Differentially methylated positions (DMPs) associated with OCD – extended information. A)** cross-sex meta-analysis, **B)** female-specific analysis. The DMPs were annotated using GRCh38/hg38. adjusted: adjusted, avg: average, CI: confidence interval, CTRL: healthy control, diff: difference, IQR: interquartile range, OCD: obsessive-compulsive disorder, OR: odds ratio, prox: proximal, SE: standard error.

**Table S2: Differentially methylated regions (DMRs) associated with OCD – extended information. A)** cross-sex meta-analysis, **B)** female-specific analysis, **C)** male-specific analysis. The DMRs were annotated using GRCh38/hg38. adjusted: adjusted, avg: average, CI: confidence interval, CTRL: healthy control, diff: difference, IQR: interquartile range, nprobe: number of CpGs in DMR, OCD: obsessive-compulsive disorder, OR: odds ratio, prox: proximal, TSS: transcription start site.

**Table S3: Interaction between sex and the DNA methylation-response relationship.** SE: standard error, BH: Benjamini-Hochberg correction

**Table S4: Sensitivity analyses.** DNA methylation at the identified differentially methylated positions (DMPs) and regions (DMRs) was compared between **(A)** OCD subgroups of non-medicated individuals and **(B)** individuals without psychiatric comorbidities, each relative to the full sample of healthy controls. FDR: false discovery rate-adjusted, SE: standard error.

**Table S5: Significant mQTLs before pruning.** Results from the **(A)** cross-sex meta-analysis, **(B)** female-specific analysis, and **(C)** male-specific analysis. Case: OCD case, Ctrl: healthy control, DMR: differentially methylated region, DNAm: DNA methylation, mQTL: methylation quantitative trait locus.

**Table S6: Significant mQTLs after pruning.** Results from the **(A)** cross-sex meta-analysis, **(B)** female-specific analysis, and **(C)** male-specific analysis. Case: OCD case, Ctrl: healthy control, DMR: differentially methylated region, DNAm: DNA methylation, mQTL: methylation quantitative trait locus.

**Table S7: Associations of SNPs in linkage disequilibrium with rs74004627 from the male-specific mQTL analysis.** Case: OCD case; Ctrl: healthy control; DMR: differentially methylated region; DNAm: DNA methylation; mQTL: methylation quantitative trait locus.

**Table S8: Top five most enriched gene ontology biological process terms.** No term remained significant after multiple testing correction.

**Table S9: GWAS hits for the genes annotated to OCD-associated DMRs.** Extracted from the PheWAS catalog[18], filtered on associations with p ≤ 5E-08. DMR: differentially methylated region, GWAS: genome-wide association study, OCD: obsessive-compulsive disorder.

**Table S10: Saliva-brain correlation.** Saliva-brain DNA methylation correlations for significant **(A)** DMPs and **(B)** CpGs within significant DMRs were extracted using previously published data (Braun et al. 2019). High absolute correlation (|ρ| > 0.5) highlighted in dark green, medium absolute correlation (|ρ| > 0.5) in light green, low absolute correlation (|ρ| > 0.3) in blue, and missing data in purple. chr: chromosome, DMP: differentially methylated position, DMR: differentially methylated region.

**Table S11: Estimated cell type proportions**. **(A)** Association of case-control status, sex, age, and smoking CpG DNA methylation with cell type proportions. **(B)** Analysis to assess whether the effect of case-control status on cell type proportions differed by sex. **(C)** Repeating the association analysis in individuals with OCD who did not take psychoactive medication. B: B cells, CD4T: CD4+ T lymphocytes, Epi: epithelial cells, Fib: fibroblasts, Mono: monocytes, Neutro: neutrophiles, NK: natural killer cells.

### Supplementary Files

**Supplementary File 1: Adapted annotation for the Illumina EPICv2 array.**

**Supplementary File 2: Visualization of differentially methylated positions (DMPs) from the cross-sex meta-analysis. (A)** Box-violin plots showing DNA methylation differences between individuals with OCD and controls. **(B)** Density plots of methylation levels, with OCD odds ratios (ORs) calculated for the highest and lowest 20% of methylation values (indicated by vertical lines). Significant ORs are marked with an asterisk (*). chr: chromosome, CTRL: healthy control, DNAm: DNA methylation, OCD: obsessive-compulsive disorder.

**Supplementary File 3: Visualization of differentially methylated regions (DMRs) from the cross-sex meta-analysis. (A)** Box-violin plots showing DNA methylation differences between individuals with OCD and controls. **(B)** Density plots of methylation levels, with OCD odds ratios (ORs) calculated for the highest and lowest 20% of methylation values (indicated by vertical lines). Significant ORs are marked with an asterisk (*). **(C)** Difference in median DNA methylation between individuals with OCD and healthy controls for each CpG site (red dots) within the DMR. chr: chromosome, CTRL: healthy control, DMR: differentially methylated region. DNAm: DNA methylation, OCD: obsessive-compulsive disorder.

#### References


1. Aryee, M. J. *et al.* Minfi: a flexible and comprehensive Bioconductor package for the analysis of Infinium DNA methylation microarrays. *Bioinformatics* **30**, 1363–1369 (2014).
2. Lehne, B. *et al.* A coherent approach for analysis of the Illumina HumanMethylation450 BeadChip improves data quality and performance in epigenome-wide association studies. *Genome Biol* **16**, 37 (2015).
3. Kaur, D. *et al.* Comprehensive evaluation of the Infinium human MethylationEPIC v2 BeadChip. *Epigenetics Communications* **3**, 6 (2023).
4. Peters, T. J. *et al.* Characterisation and reproducibility of the HumanMethylationEPIC v2.0 BeadChip for DNA methylation profiling. *BMC Genomics* **25**, 251 (2024).
5. Heiss, J. A. & Just, A. C. Identifying mislabeled and contaminated DNA methylation microarray data: an extended quality control toolset with examples from GEO. *Clinical Epigenetics* **10**, 73 (2018).
6. Pidsley, R. *et al.* A data-driven approach to preprocessing Illumina 450K methylation array data. *BMC Genomics* **14**, 293 (2013).
7. Teschendorff, A. E. *et al.* A beta-mixture quantile normalization method for correcting probe design bias in Illumina Infinium 450 k DNA methylation data. *Bioinformatics* **29**, 189–196 (2013).
8. Tian, Y. *et al.* ChAMP: updated methylation analysis pipeline for Illumina BeadChips. *Bioinformatics* **33**, 3982–3984 (2017).
9. Peters, T. J. *et al.* De novo identification of differentially methylated regions in the human genome. *Epigenetics & Chromatin* **8**, 6 (2015).
10. Höffler, K. D. *et al.* Optimizing Genetic Ancestry Adjustment in DNA Methylation Studies: A Comparative Analysis of Approaches. Preprint at https://doi.org/10.21203/rs.3.rs-6580295/v1 (2025).
11. Zheng, S. C. *et al.* A Novel Cell-Type Deconvolution Algorithm Reveals Substantial Contamination by Immune Cells in Saliva, Buccal and Cervix. *Epigenomics* **10**, 925–940 (2018).

12. Zheng, S. C., Breeze, C. E., Beck, S. & Teschendorff, A. E. Identification of differentially methylated cell types in epigenome-wide association studies. *Nat Methods* **15**, 1059–1066 (2018).
13. Moore, J. E. *et al.* Expanded encyclopaedias of DNA elements in the human and mouse genomes. *Nature* **583**, 699–710 (2020).
14. Perez, G. *et al.* The UCSC Genome Browser database: 2025 update. *Nucleic Acids Res* **53**, D1243–D1249 (2025).
15. Ritchie, M. E. *et al.* limma powers differential expression analyses for RNA-sequencing and microarray studies. *Nucleic Acids Res* **43**, e47 (2015).
16. Hinrichs, A. S. *et al.* The UCSC Genome Browser Database: update 2006. *Nucleic Acids Res* **34**, D590-598 (2006).
17. Purcell, S. *et al.* PLINK: a tool set for whole-genome association and population-based linkage analyses. *Am J Hum Genet* **81**, 559–575 (2007).
18. Watanabe, K. *et al.* A global overview of pleiotropy and genetic architecture in complex traits. *Nat Genet* **51**, 1339–1348 (2019).
19. Braun, P. R. *et al.* Genome-wide DNA methylation comparison between live human brain and peripheral tissues within individuals. *Transl Psychiatry* **9**, 47 (2019).
