## Supplementary File 2 for "Methylome-Wide Association Study of Obsessive-Compulsive Disorder"

### Differentially methylated positions (DMPs)

A) Box-violin plots showing DNA methylation differences between individuals with OCD and controls.

B) Density plots of methylation levels, with OCD odds ratios (ORs) calculated for the highest and lowest 20% of methylation values (indicated by vertical lines). Significant ORs are marked with an asterisk (*).

chr: chromosome, CTRL: healthy control, DMP: differentially methylated position. DNAm: DNA methylation, OCD: obsessive-compulsive disorder.

### sex-stratified meta-analysis

### cg00050692 – DNMT3A

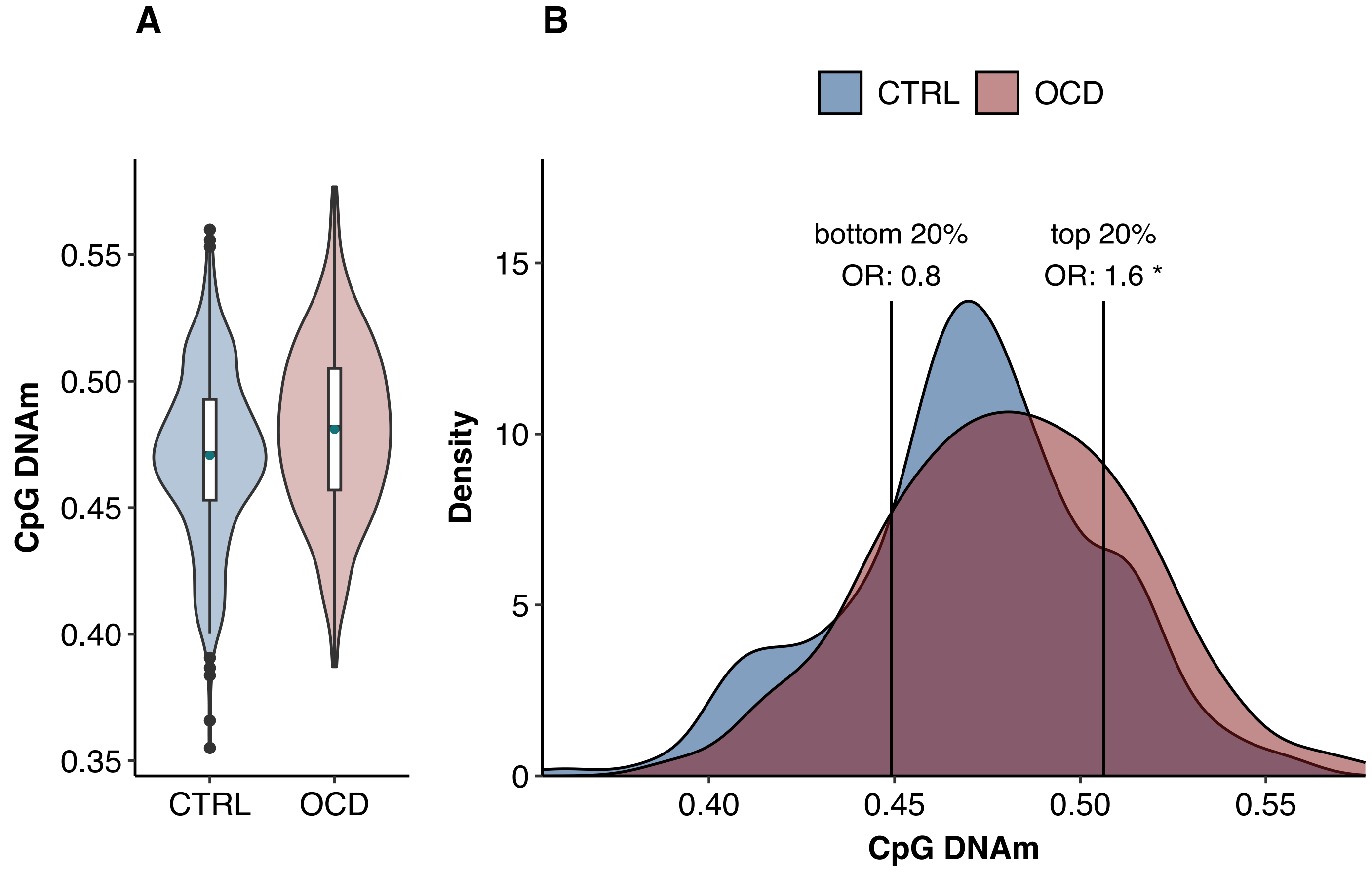

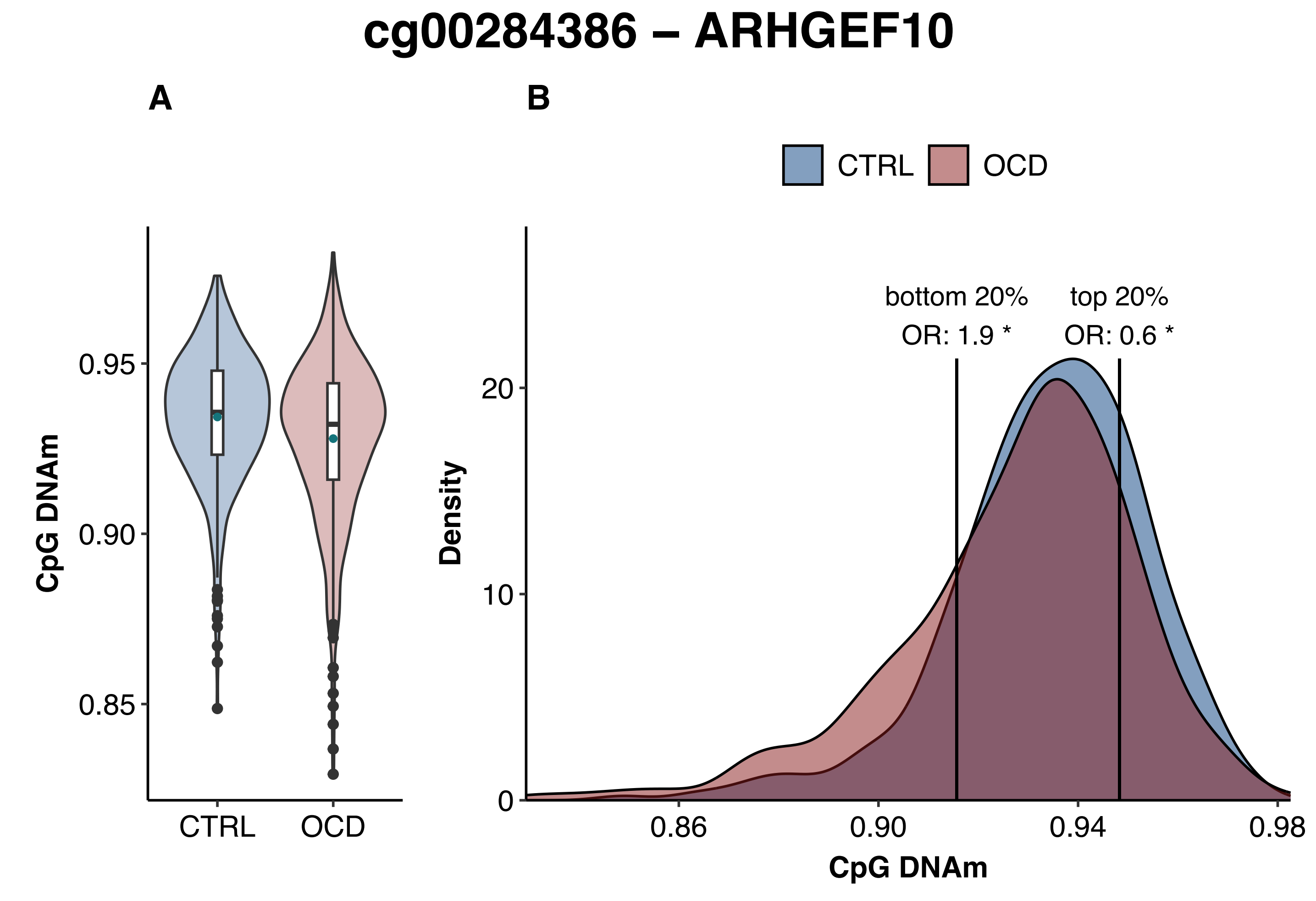
cg00284386 – ARHGEF10
A
B
CTRL
OCD
CpG DNAm
0.95
0.90
0.85
CTRL
OCD
bottom 20%
OR: 1.9 *
top 20%
OR: 0.6 *
Density
20
10
0
0.86
0.90
0.94
0.98
CpG DNAm

### cg01188427 – CSF1

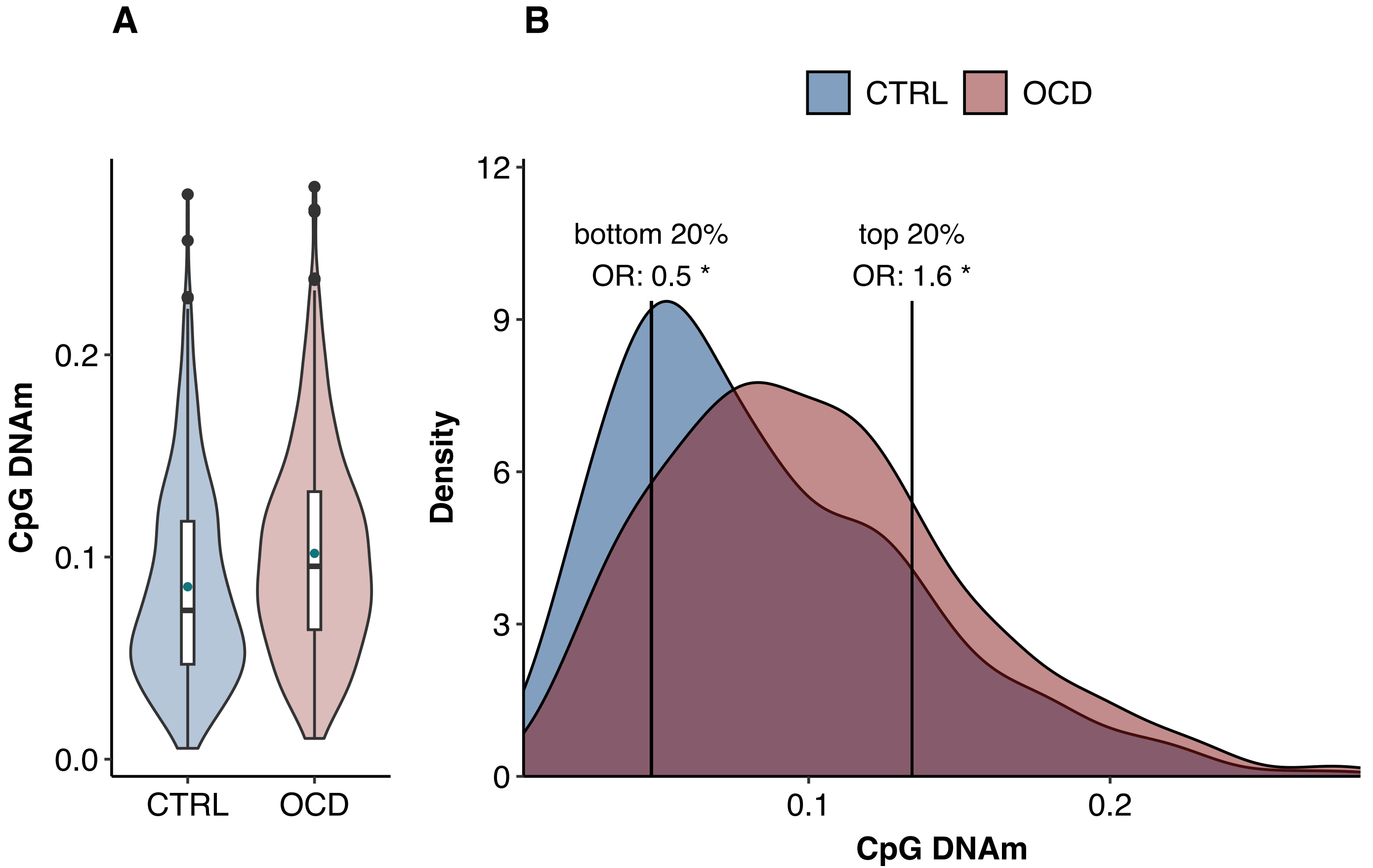

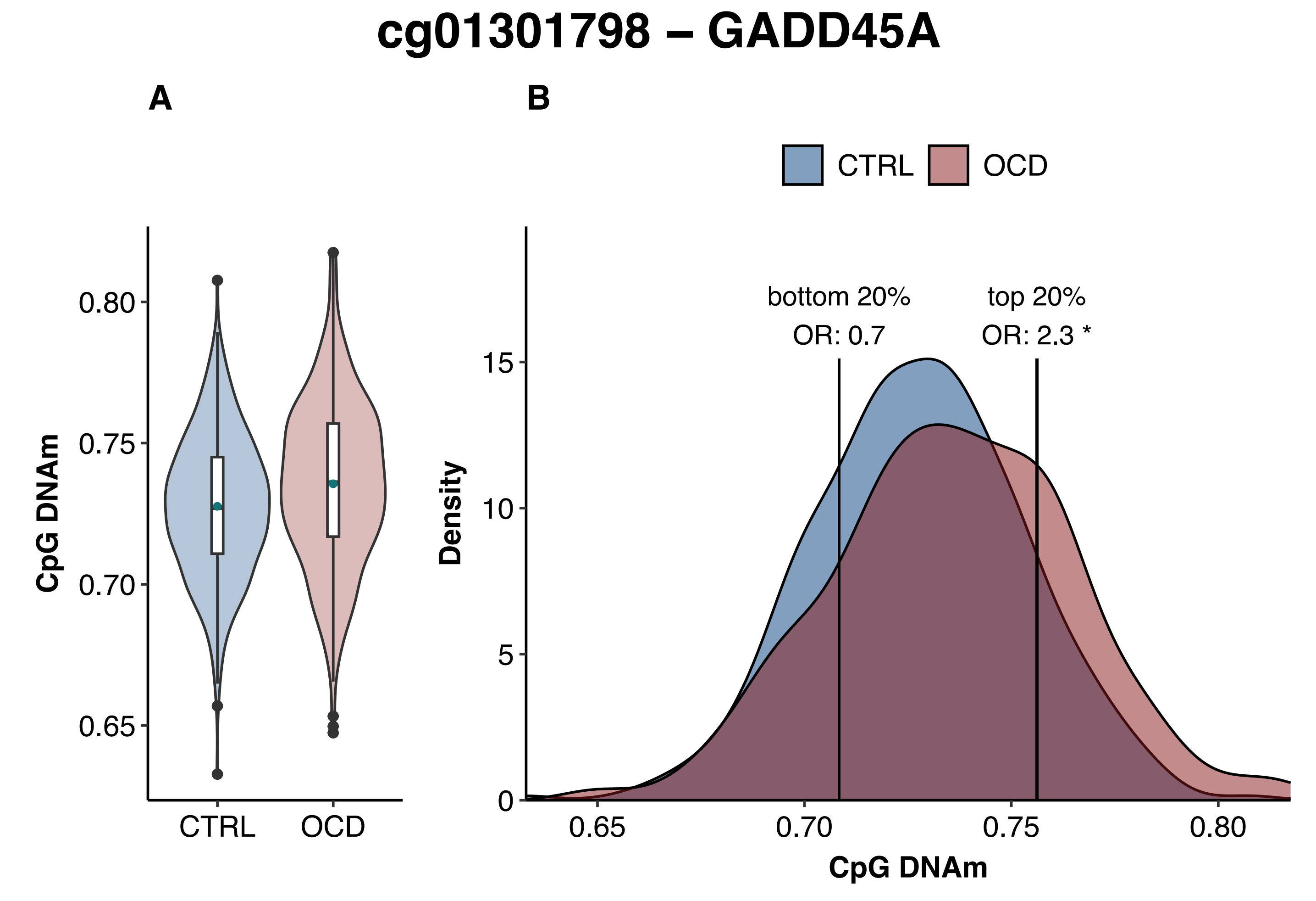

cg01301798 – GADD45A
A
B
CTRL
OCD
CpG DNAm
0.80
0.75
0.70
0.65
CTRL
OCD
bottom 20%
OR: 0.7
top 20%
OR: 2.3 *
Density
15
10
5
0
0.65
0.70
0.75
0.80
CpG DNAm

### cg01322839 – TRIM14

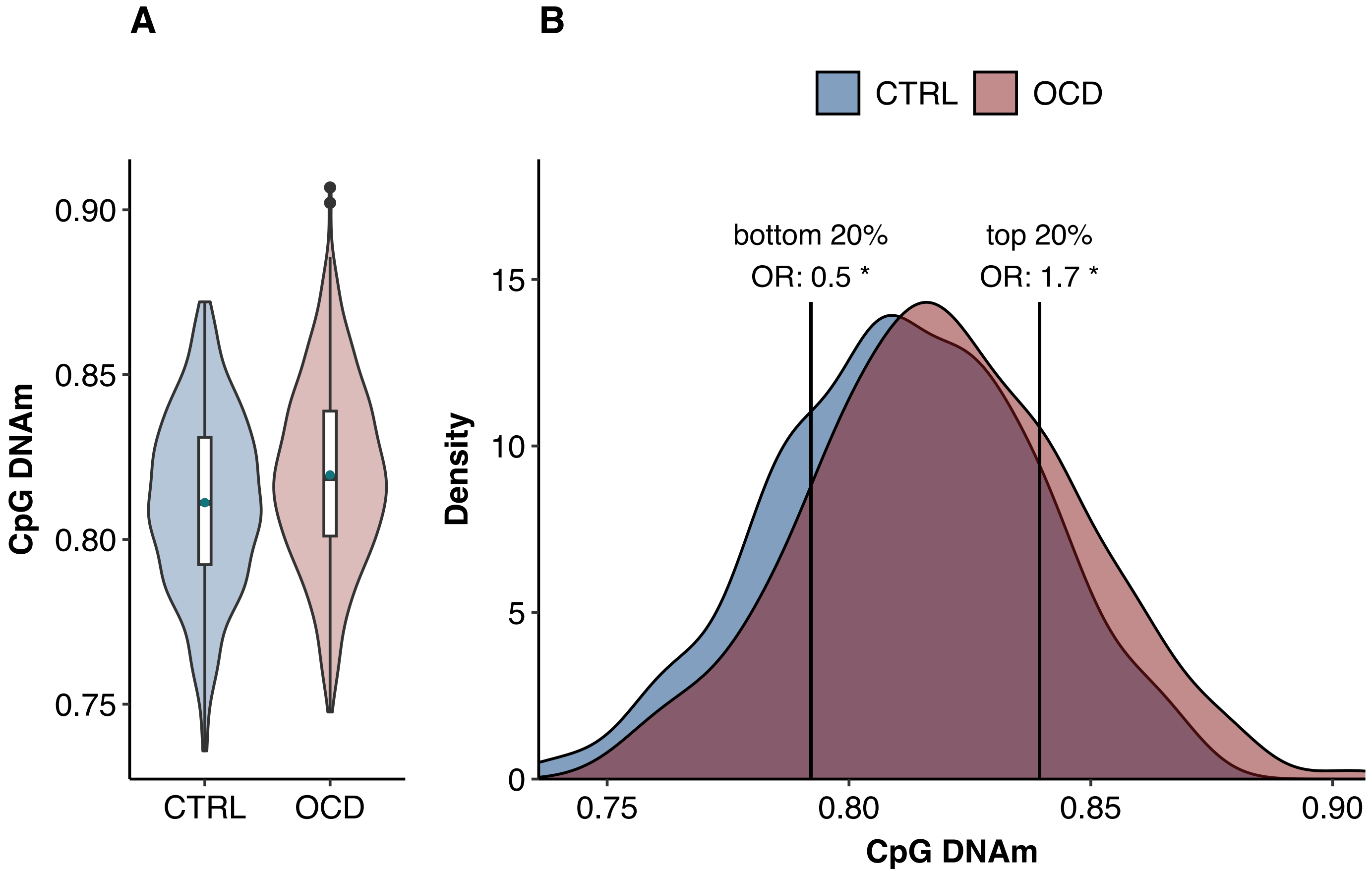

### cg01593826 – UNANNOTATED

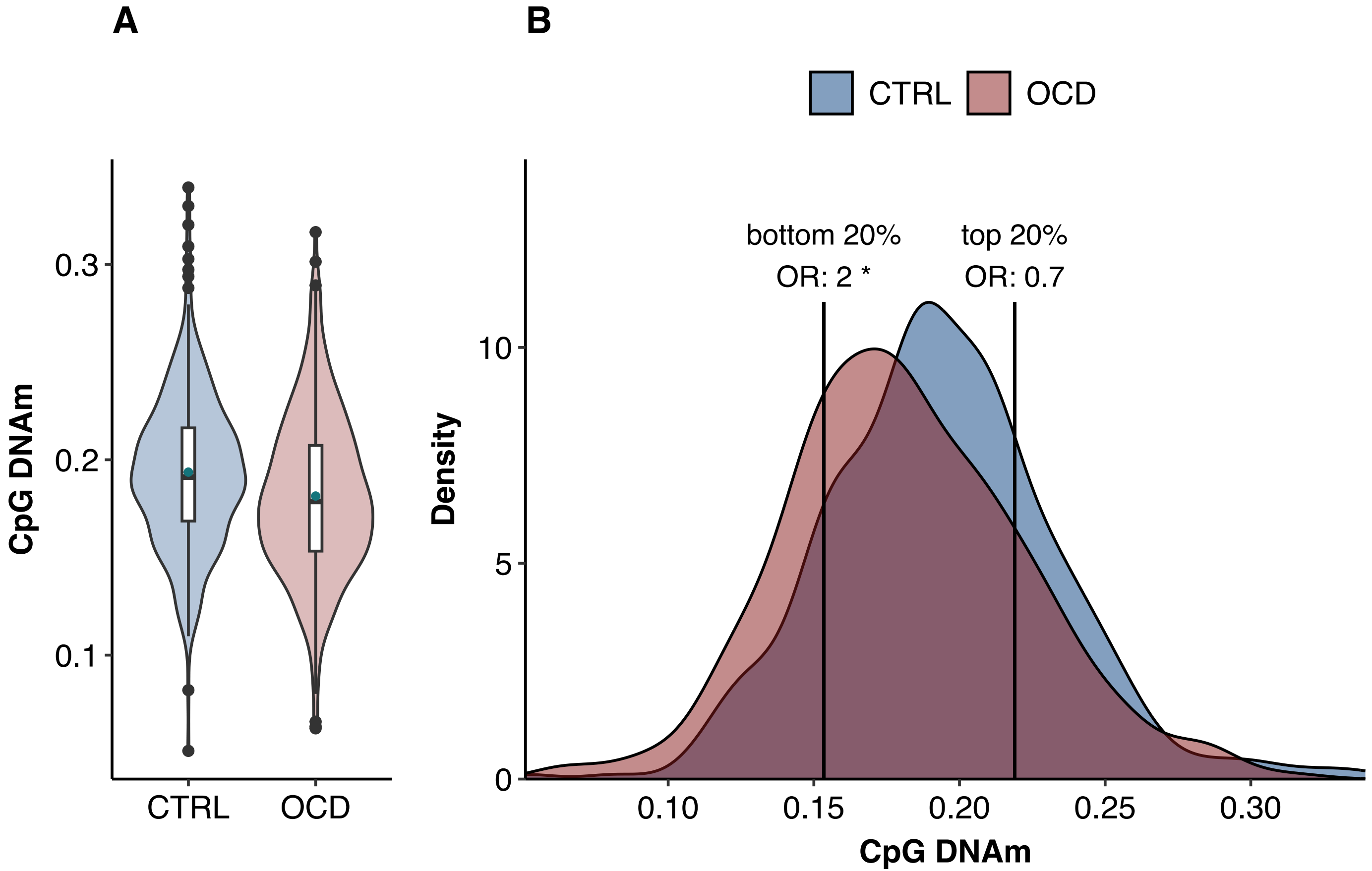

### cg02456934 – ZNRF1

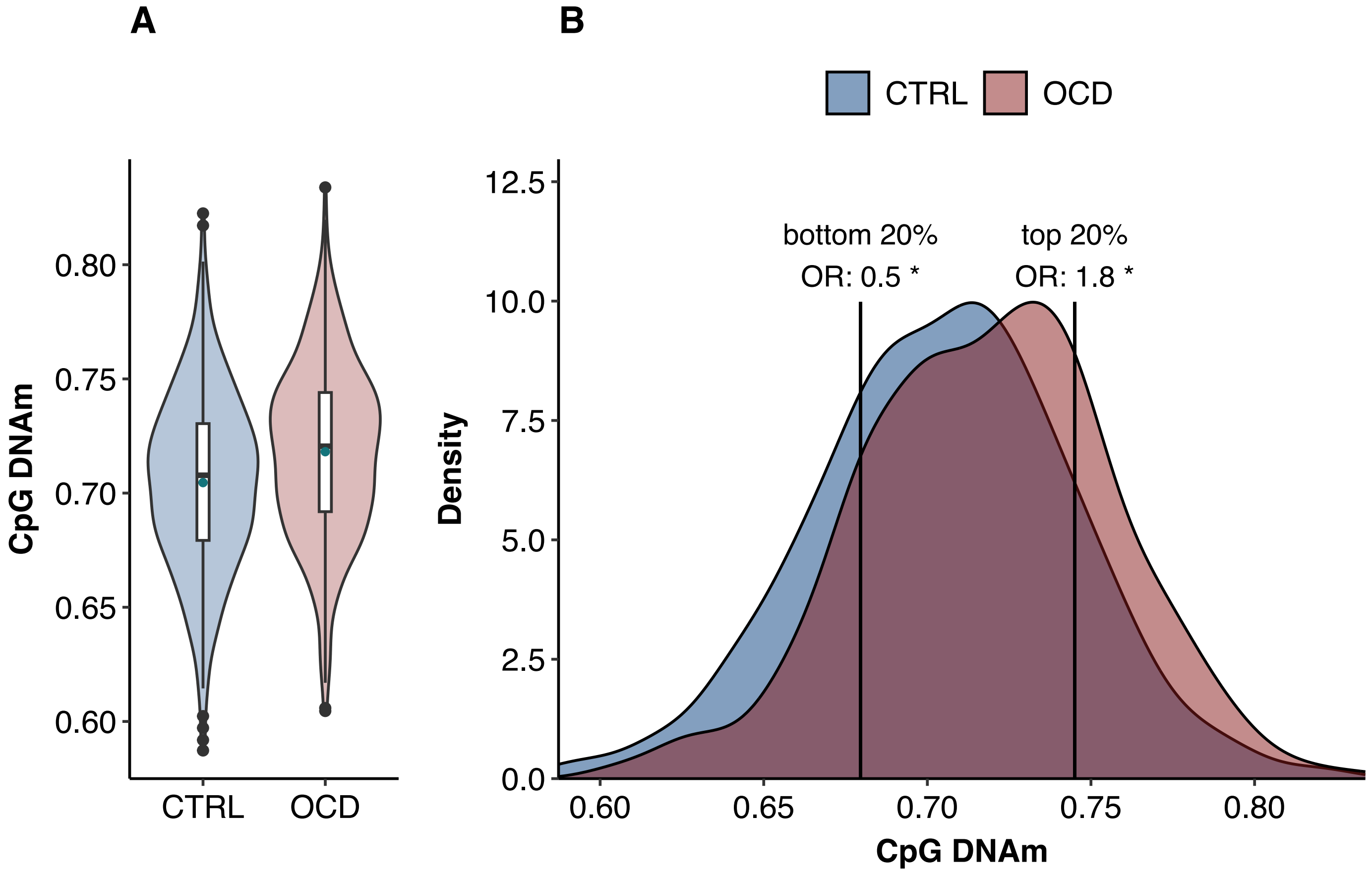

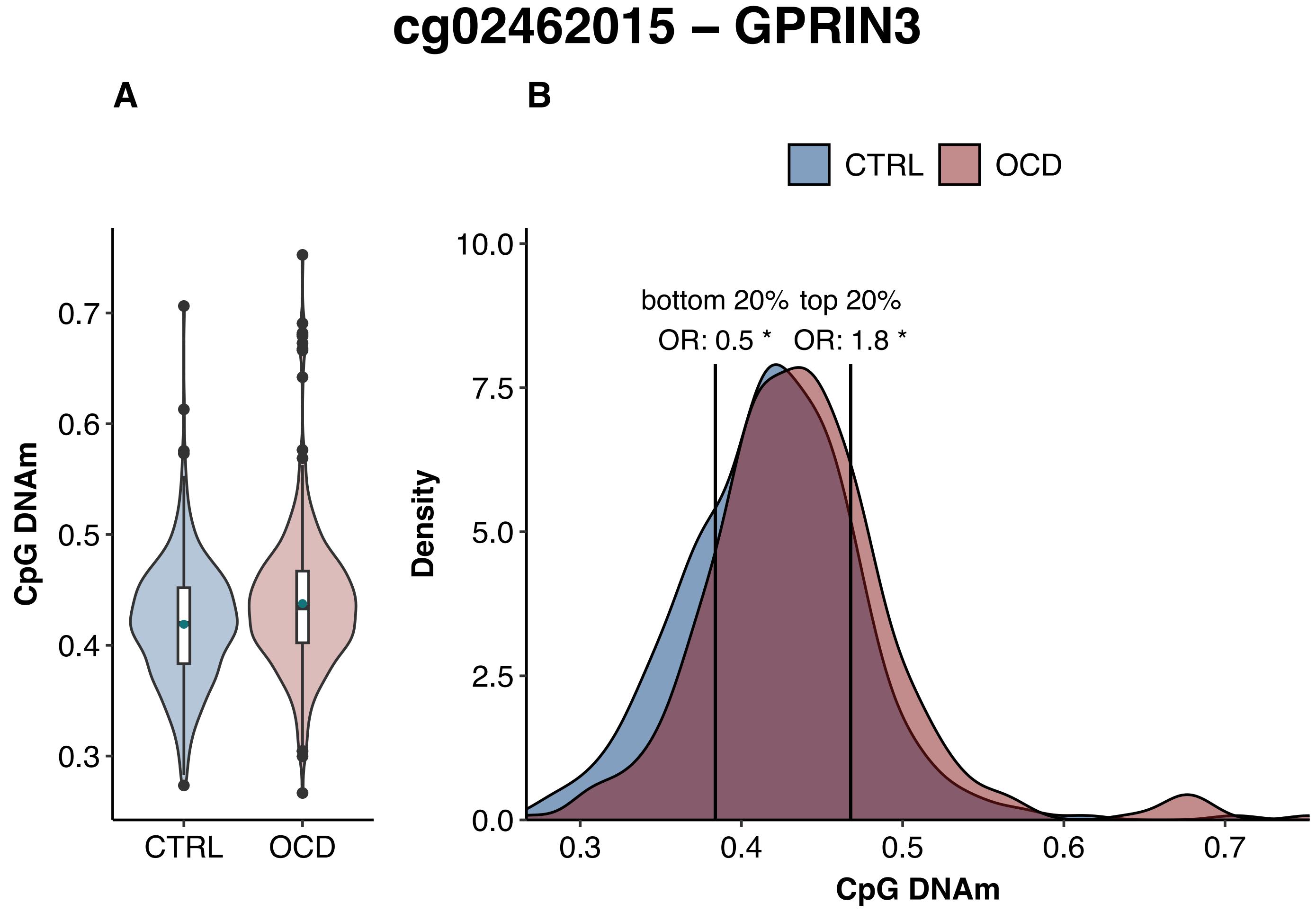
cg02462015 – GPRIN3
A
B
CTRL
OCD
CpG DNAm
0.7
0.6
0.5
0.4
0.3
CTRL
OCD
Density
10.0
7.5
5.0
2.5
0.0
bottom 20%
top 20%
OR: 0.5 *
OR: 1.8 *
0.3
0.4
0.5
0.6
0.7
CpG DNAm

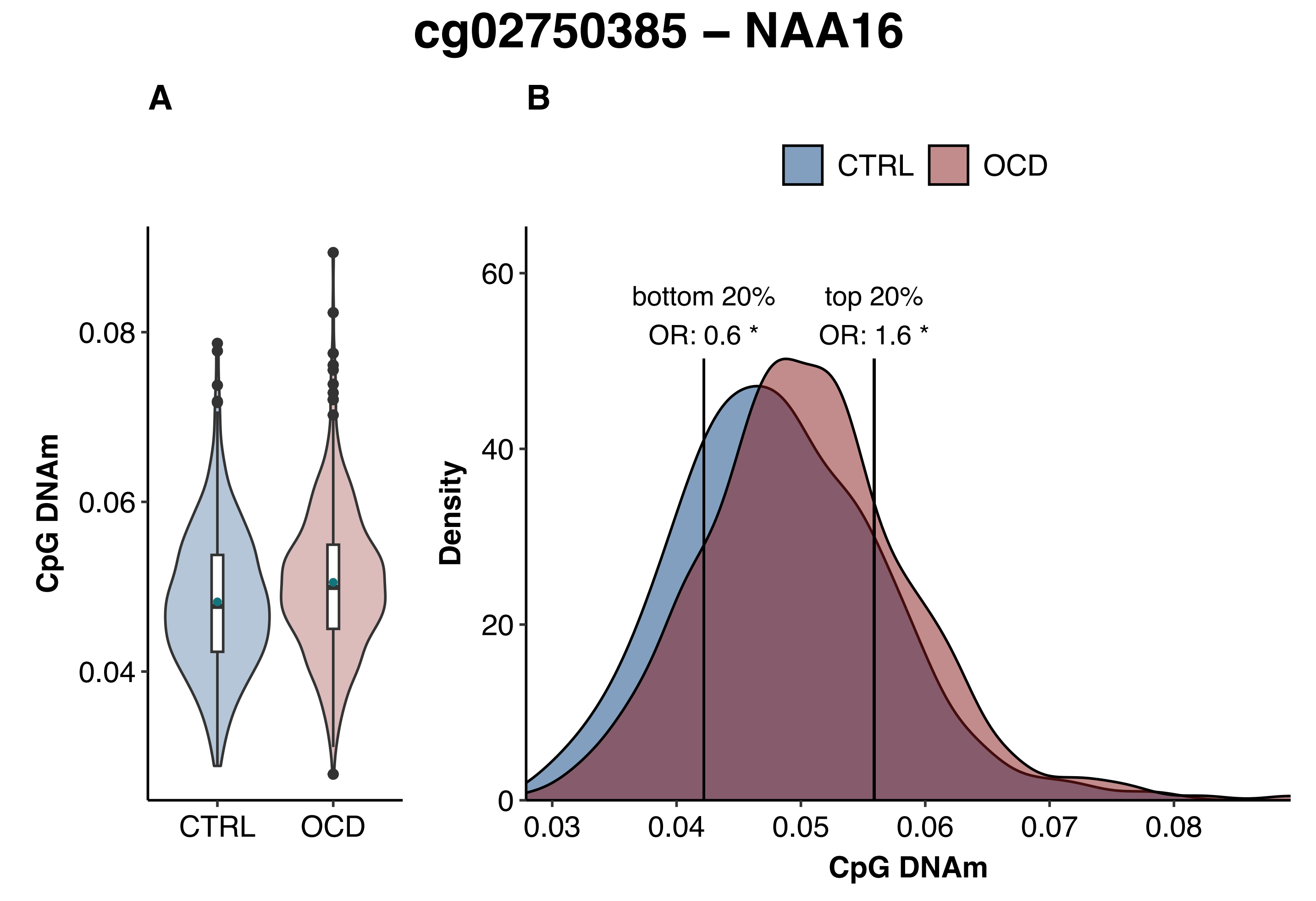
cg02750385 – NAA16
A
B
CTRL
OCD
CpG DNAm
0.08
0.06
0.04
CTRL
OCD
Density
60
40
20
0
bottom 20%
OR: 0.6 *
top 20%
OR: 1.6 *
0.03
0.04
0.05
0.06
0.07
0.08
CpG DNAm

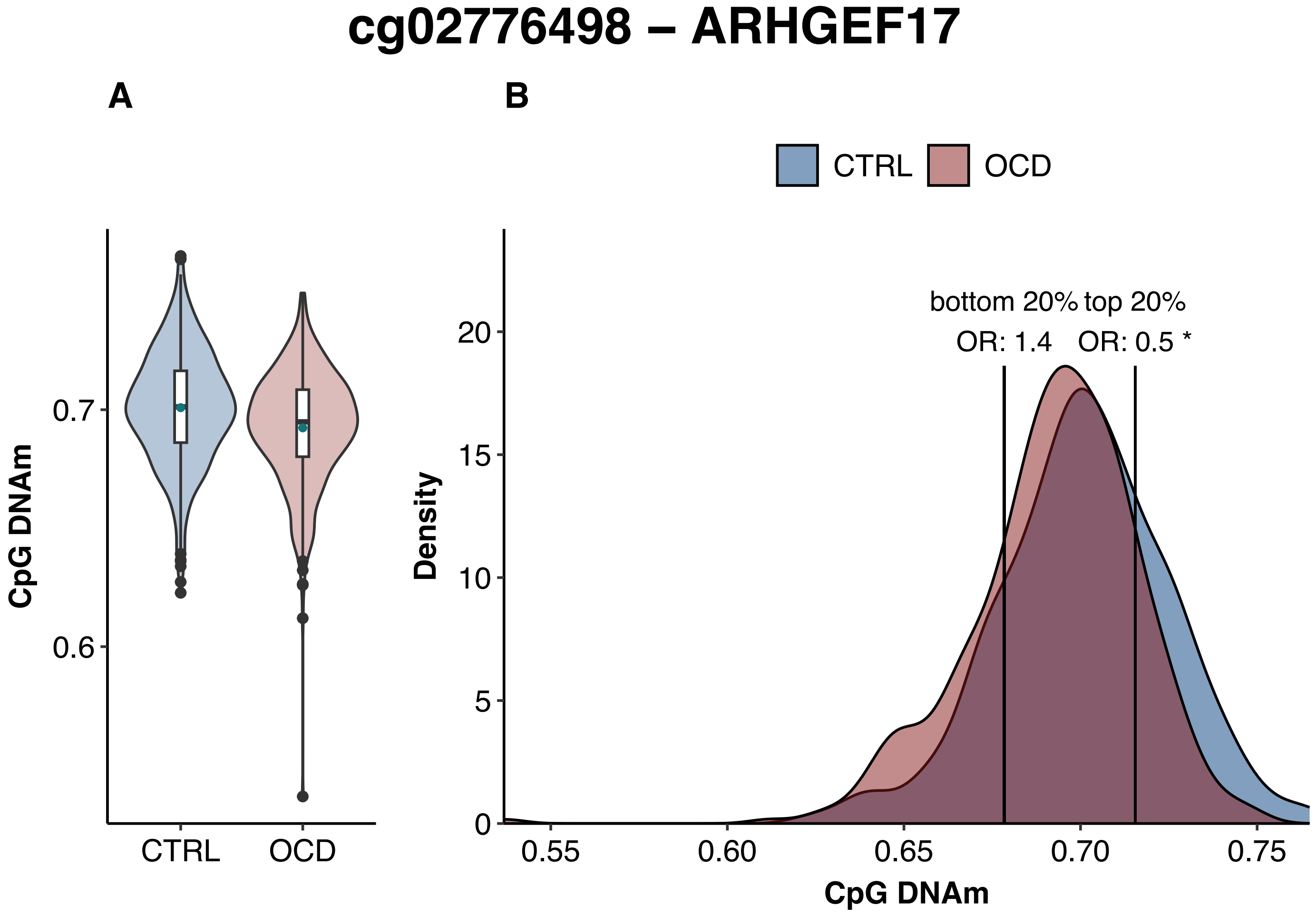

cg02776498 – ARHGEF17
A
B
CTRL
OCD
CpG DNAm
0.7
0.6
CTRL
OCD
bottom 20%
top 20%
OR: 1.4
OR: 0.5 *
Density
20
15
10
5
0
0.55
0.60
0.65
0.70
0.75
CpG DNAm

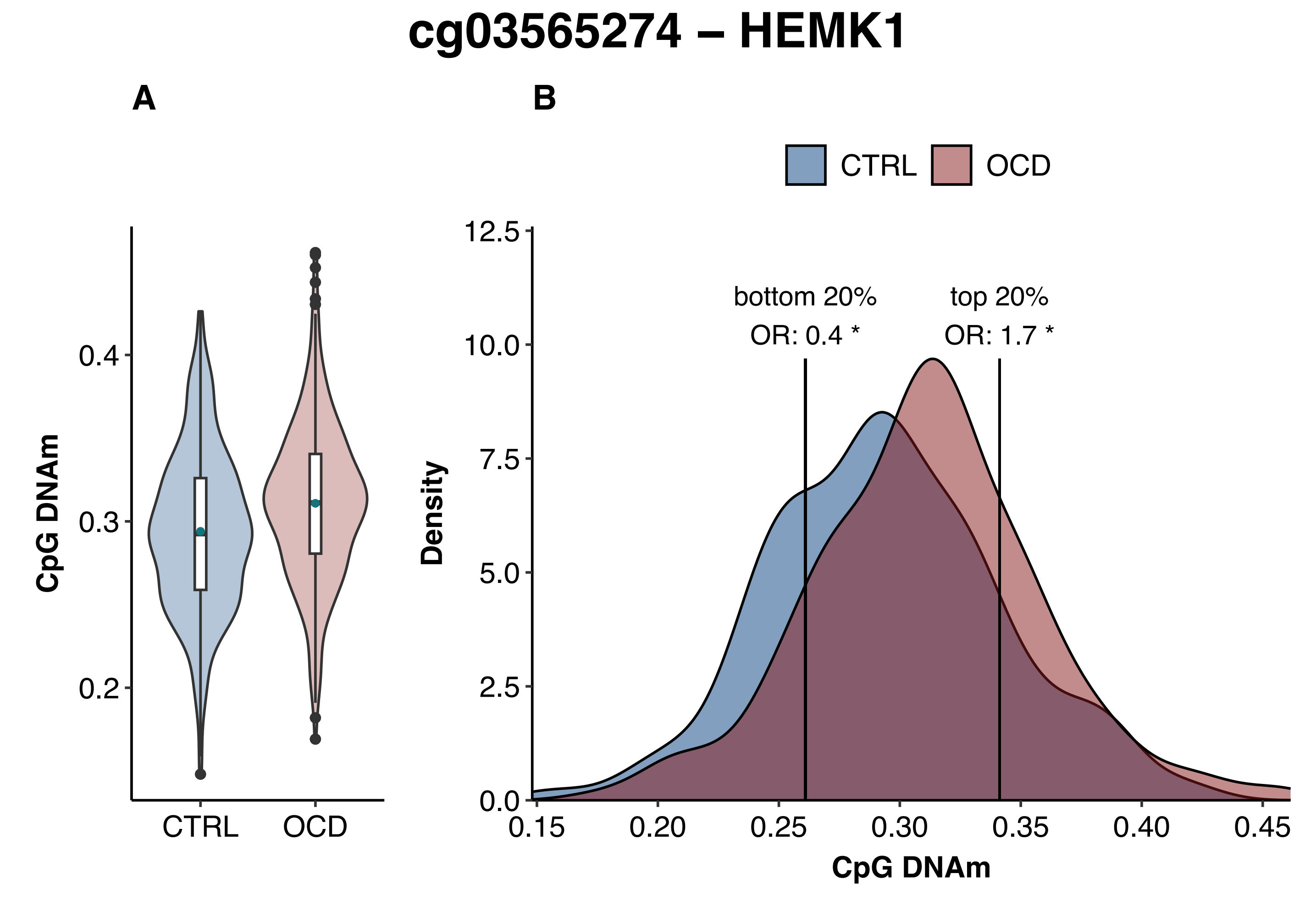
cg03565274 – HEMK1
A
B
CTRL
OCD
CpG DNAm
0.4
0.3
0.2
CTRL
OCD
bottom 20%
OR: 0.4 *
top 20%
OR: 1.7 *
Density
12.5
10.0
7.5
5.0
2.5
0.0
0.15
0.20
0.25
0.30
0.35
0.40
0.45
CpG DNAm

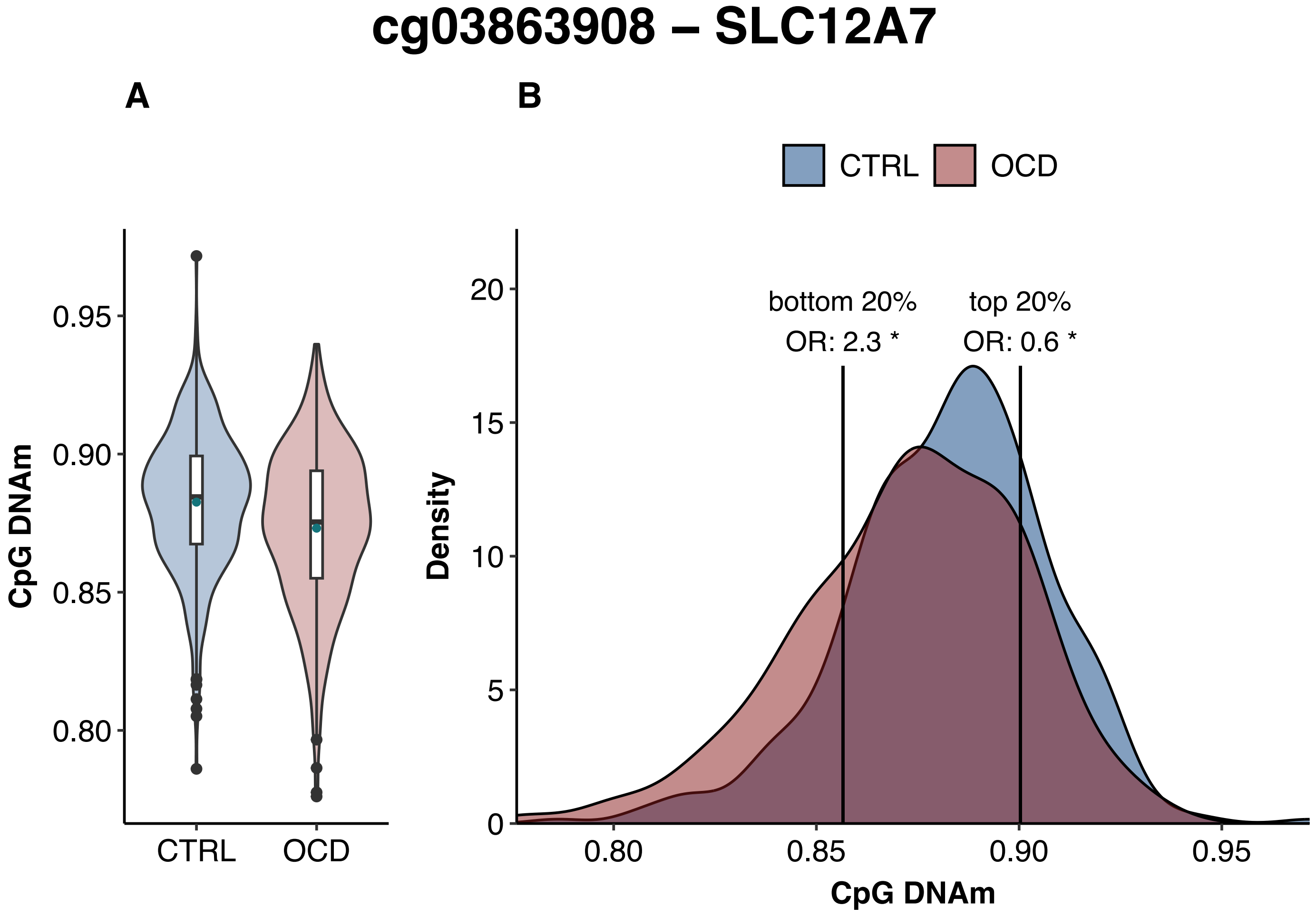
cg03863908 – SLC12A7
A
B
CTRL
OCD
CpG DNAm
0.95
0.90
0.85
0.80
CTRL
OCD
Density
20
15
10
5
0
bottom 20%
OR: 2.3 *
top 20%
OR: 0.6 *
0.80
0.85
0.90
0.95
CpG DNAm

### cg04497870 – HIVEP3

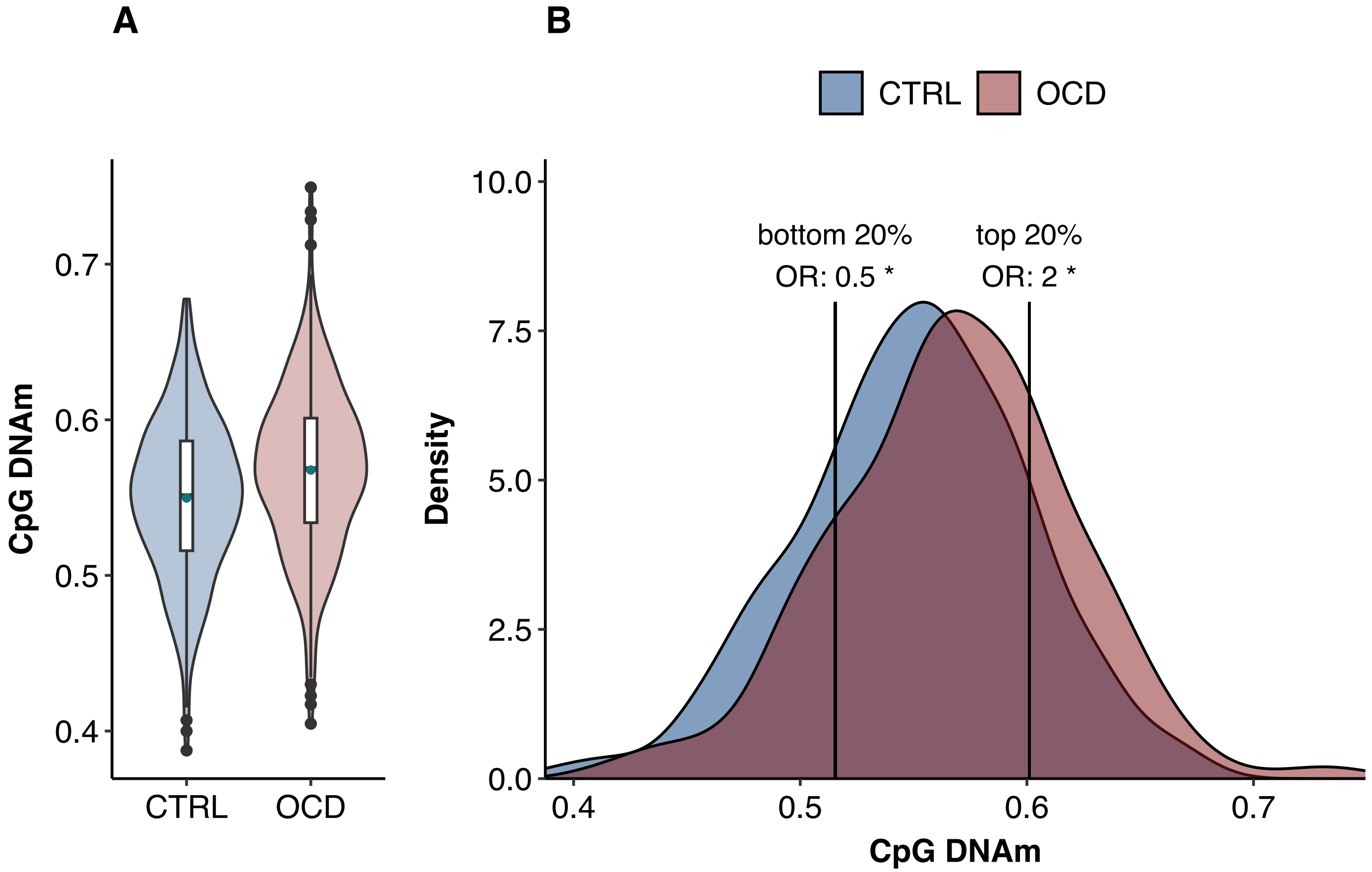

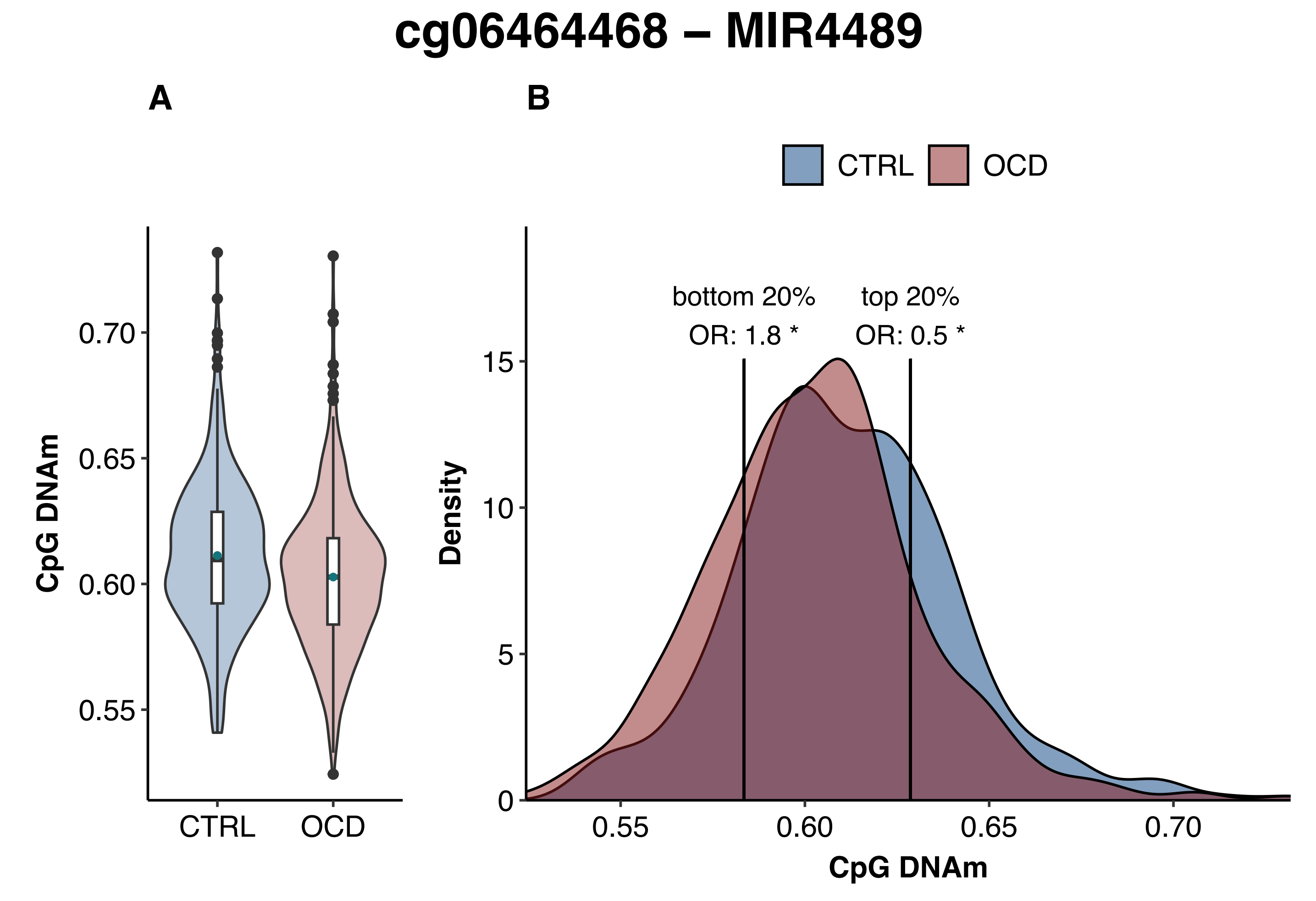
cg06464468 – MIR4489
A
B
CTRL
OCD
CpG DNAm
0.70
0.65
0.60
0.55
CTRL
OCD
bottom 20%
OR: 1.8 *
top 20%
OR: 0.5 *
Density
15
10
5
0
0.55
0.60
0.65
0.70
CpG DNAm

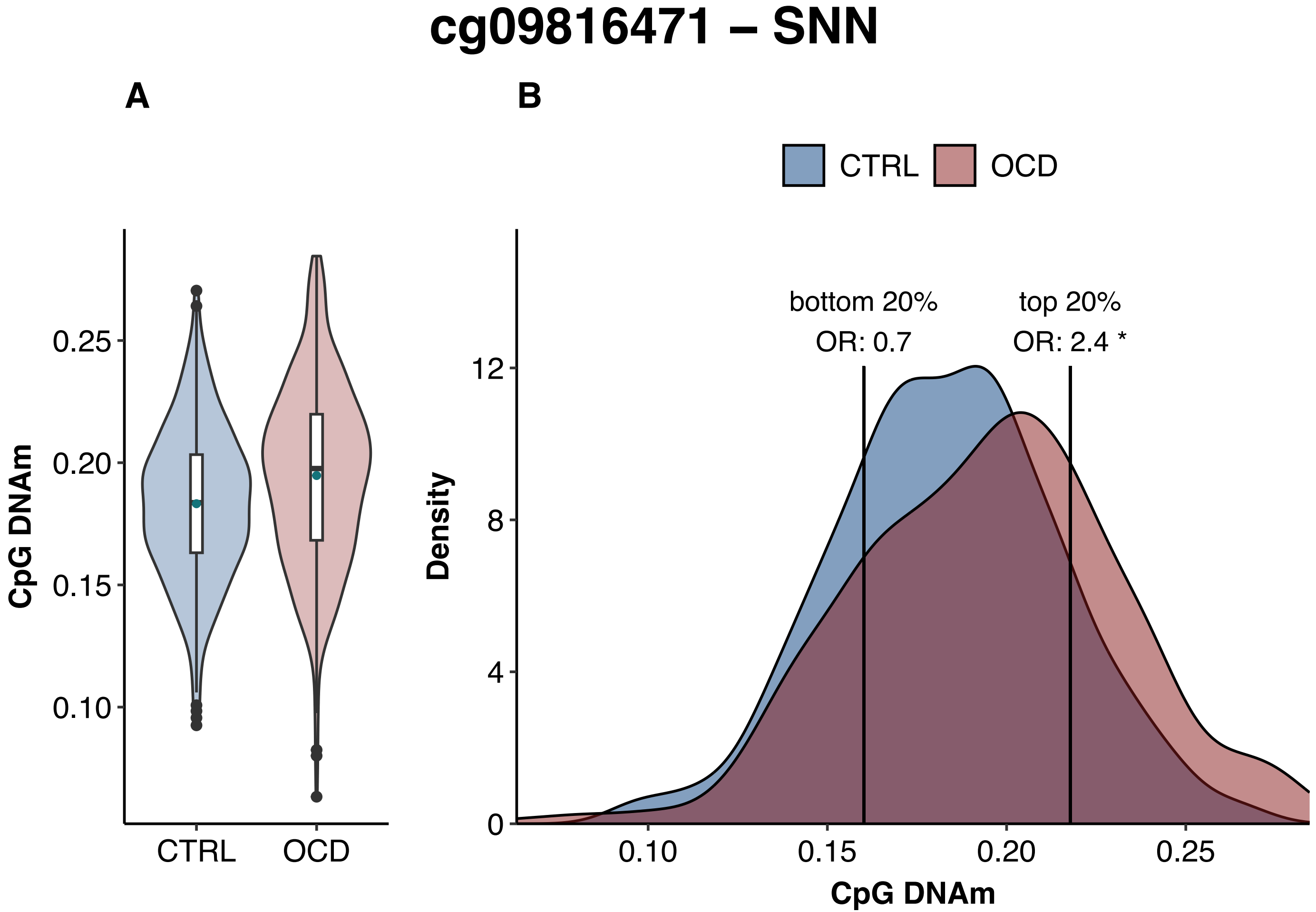

cg09816471 – SNN
A
B
CTRL
OCD
CpG DNAm
0.25
0.20
0.15
0.10
CTRL
OCD
bottom 20%
OR: 0.7
top 20%
OR: 2.4 *
Density
12
8
4
0
0.10
0.15
0.20
0.25
CpG DNAm

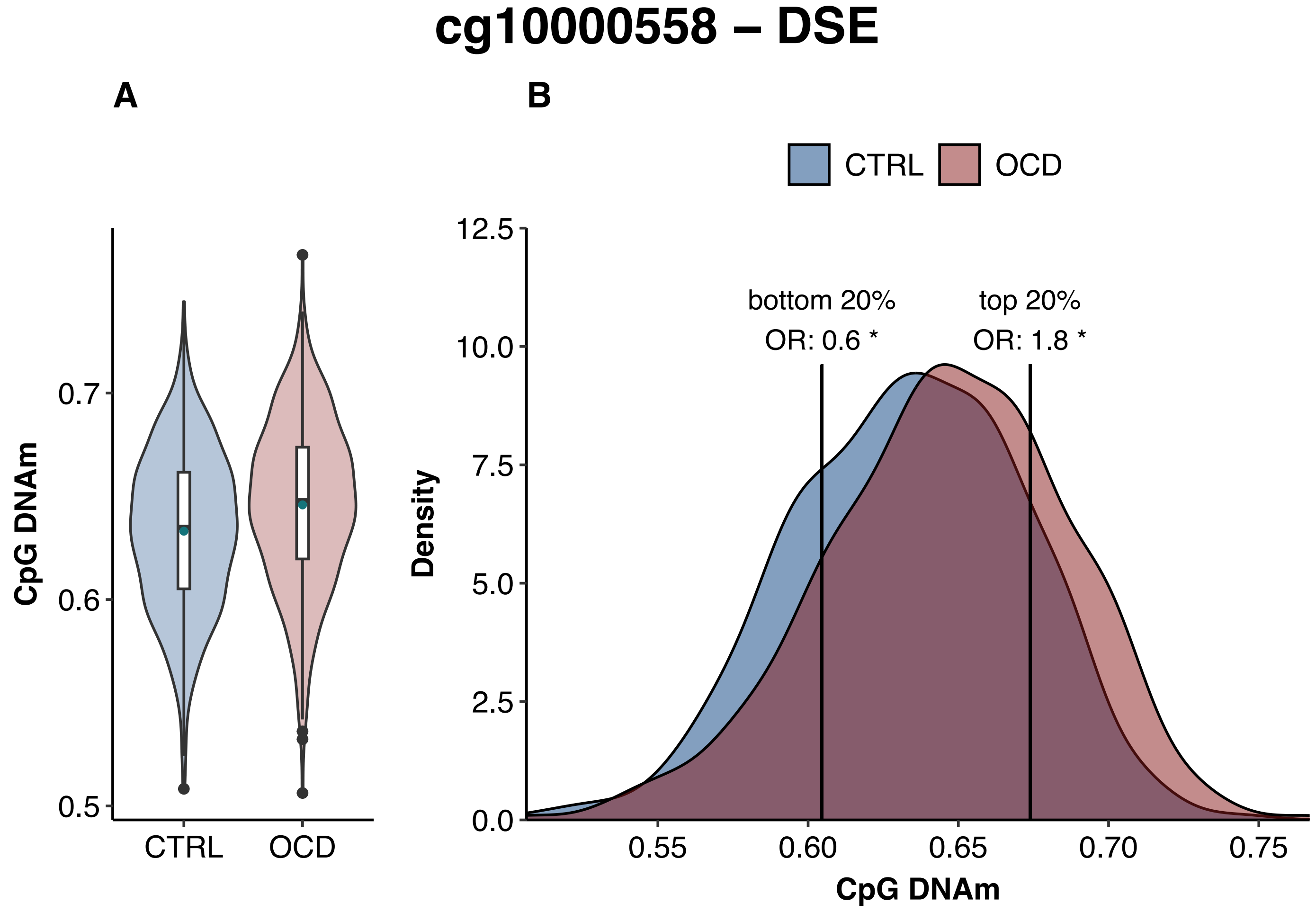
cg10000558 – DSE
A
B
CTRL
OCD
CpG DNAm
0.7
0.6
0.5
CTRL
OCD
Density
12.5
10.0
7.5
5.0
2.5
0.0
bottom 20%
OR: 0.6 *
top 20%
OR: 1.8 *
0.55
0.60
0.65
0.70
0.75
CpG DNAm

### cg10335493 – CCR1

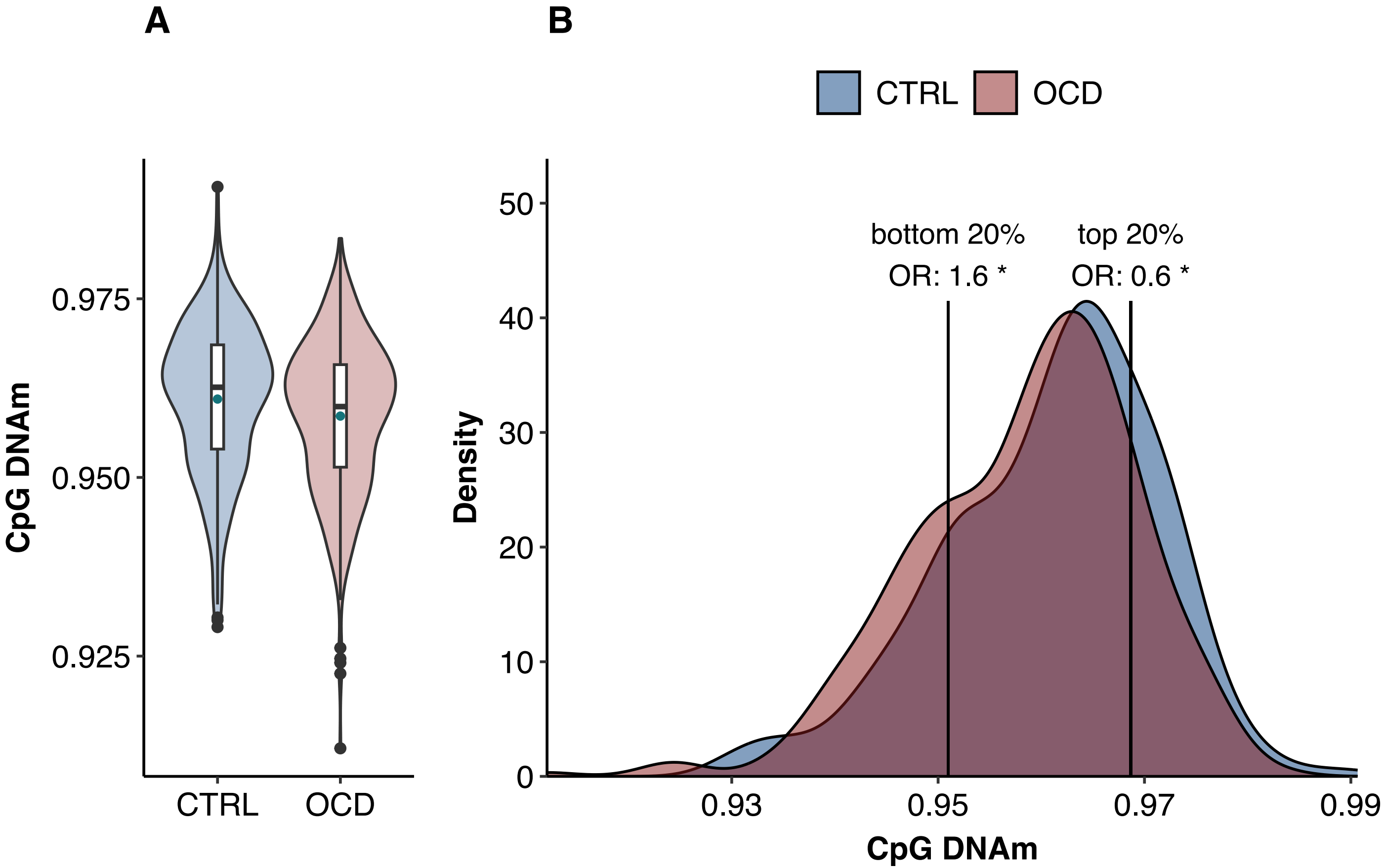

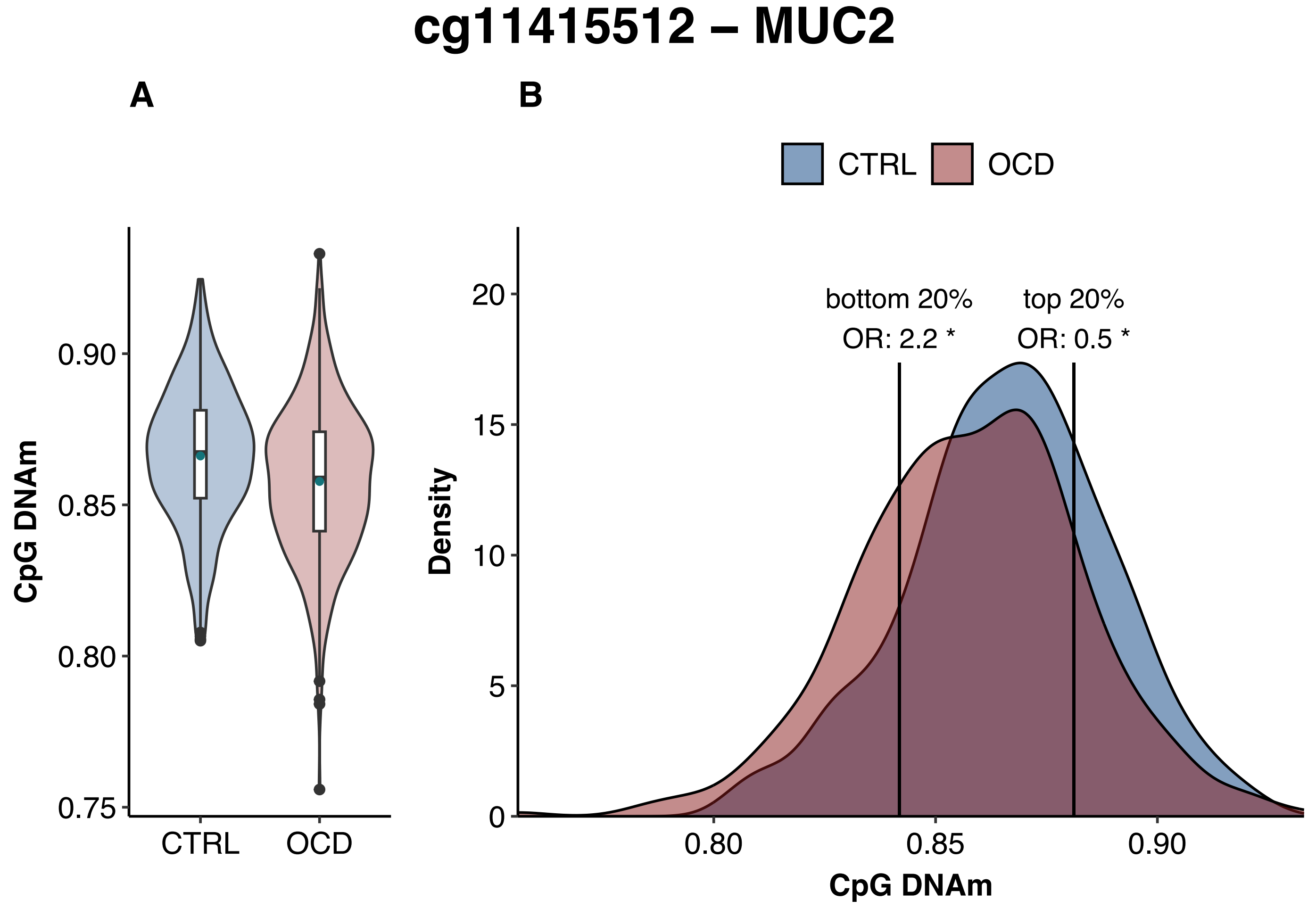
cg11415512 – MUC2
A
B
CTRL
OCD
CpG DNAm
0.90
0.85
0.80
0.75
CTRL
OCD
Density
20
15
10
5
0
bottom 20%
OR: 2.2 *
top 20%
OR: 0.5 *
0.80
0.85
0.90
CpG DNAm

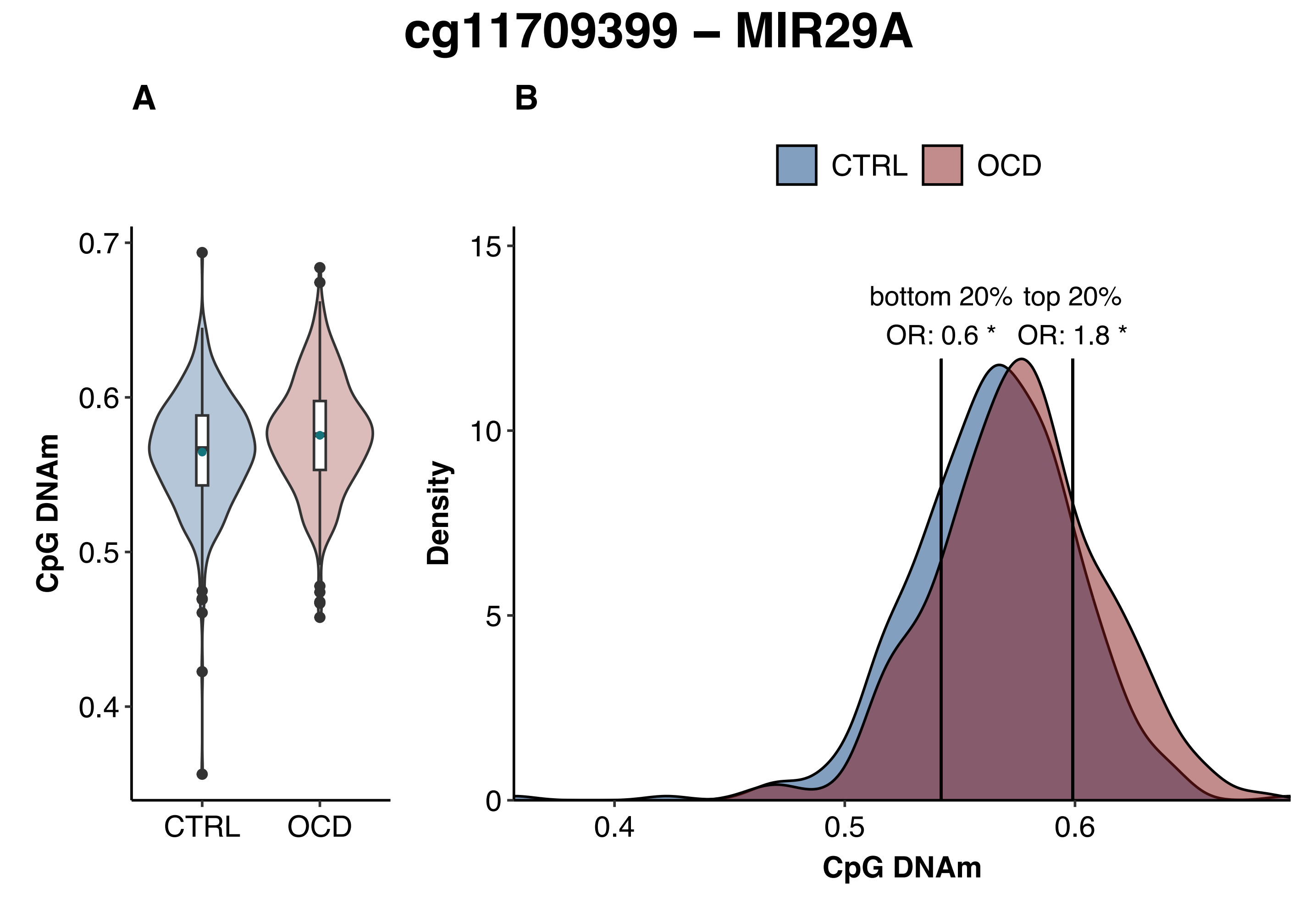
cg11709399 – MIR29A
A
B
CTRL
OCD
CpG DNAm
0.7
0.6
0.5
0.4
CTRL
OCD
Density
15
10
5
0
bottom 20%
OR: 0.6 *
top 20%
OR: 1.8 *
0.4
0.5
0.6
CpG DNAm

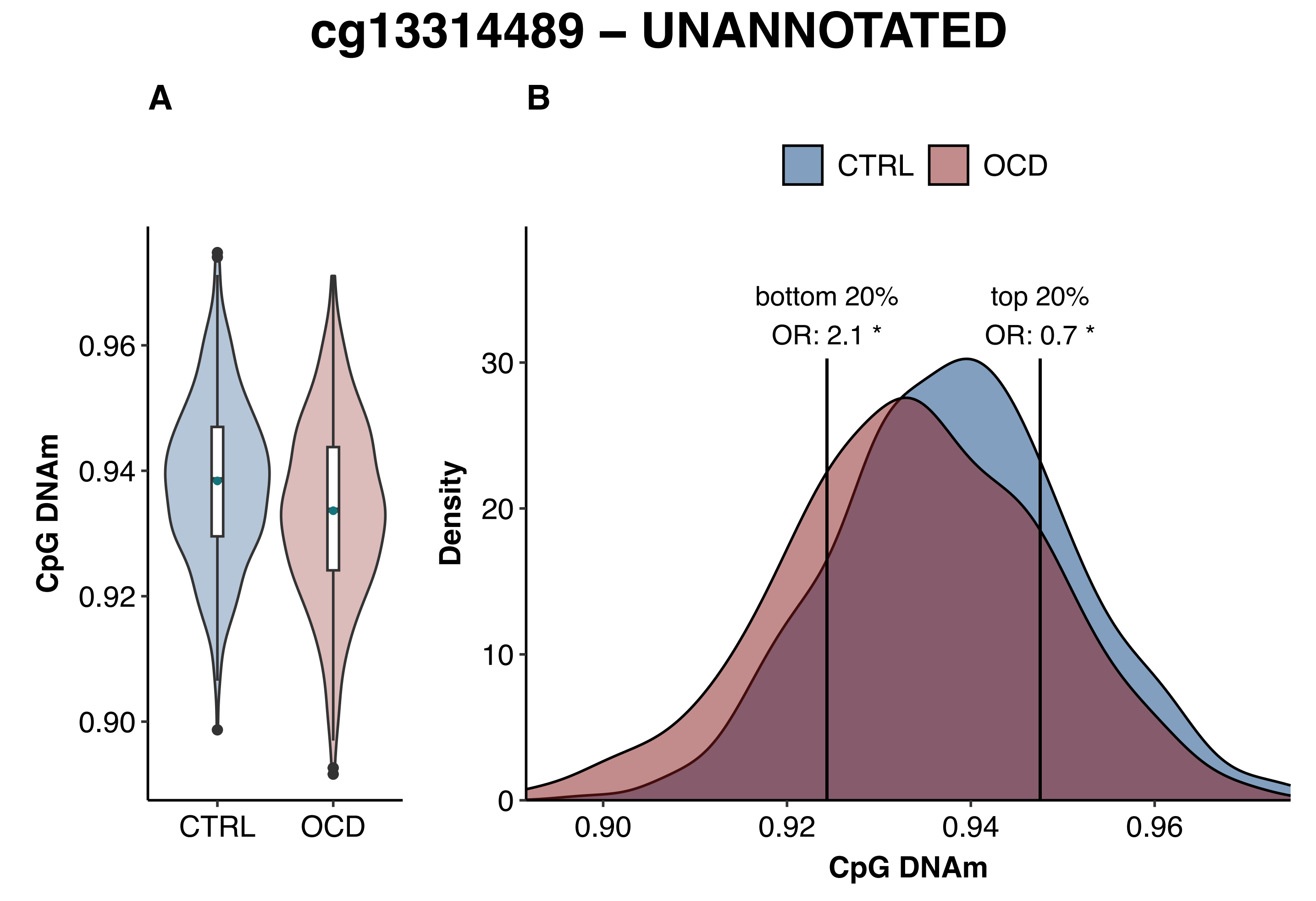

cg13314489 – UNANNOTATED
A
B
CTRL
OCD
CpG DNAm
0.96
0.94
0.92
0.90
CTRL
OCD
bottom 20%
OR: 2.1 *
top 20%
OR: 0.7 *
Density
30
20
10
0
0.90
0.92
0.94
0.96
CpG DNAm

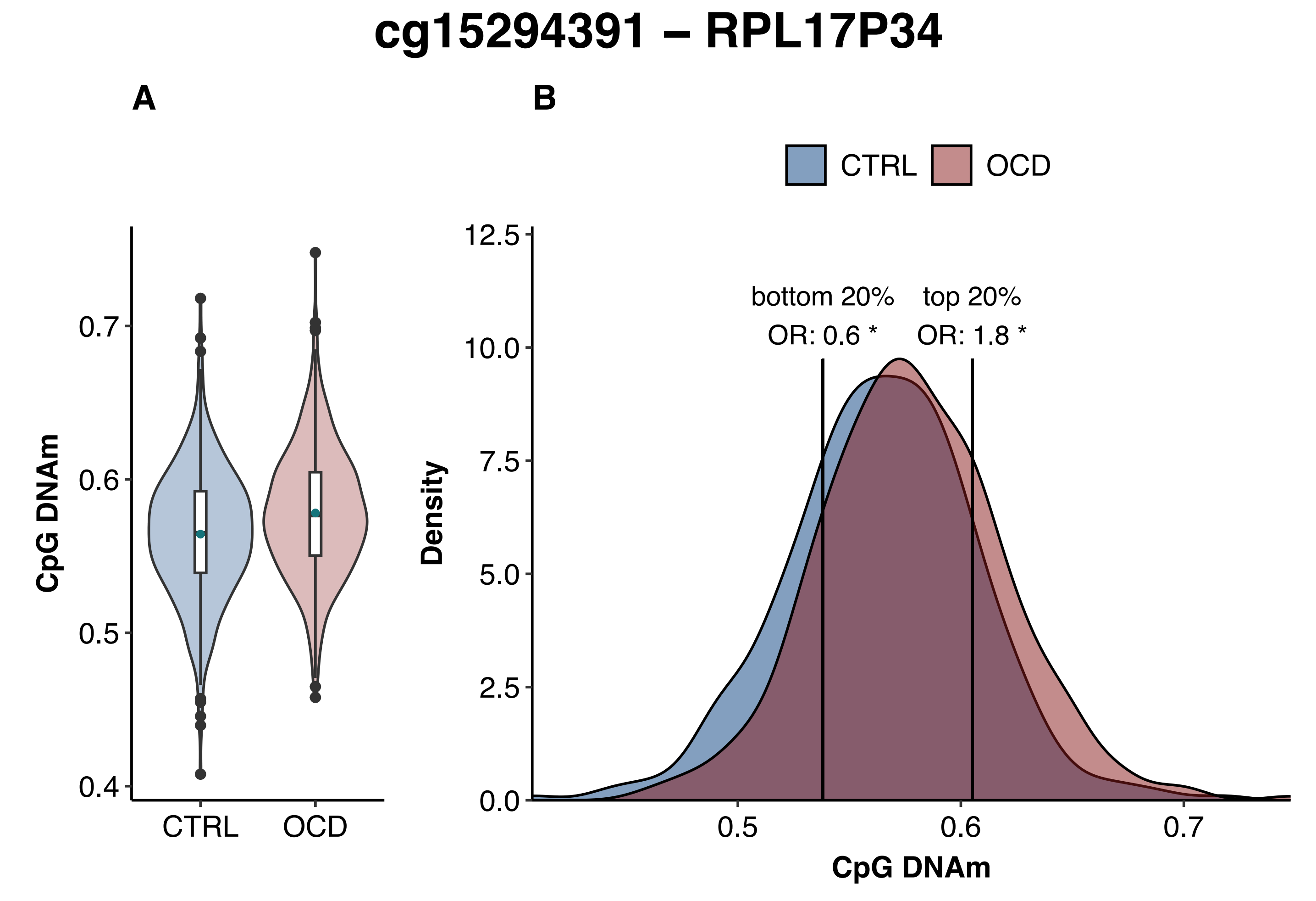
cg15294391 – RPL17P34
A
B
CTRL
OCD
bottom 20%
OR: 0.6 *
top 20%
OR: 1.8 *
CpG DNAm
Density
0.4
0.5
0.6
0.7
0.0
2.5
5.0
7.5
10.0
12.5
CTRL
OCD

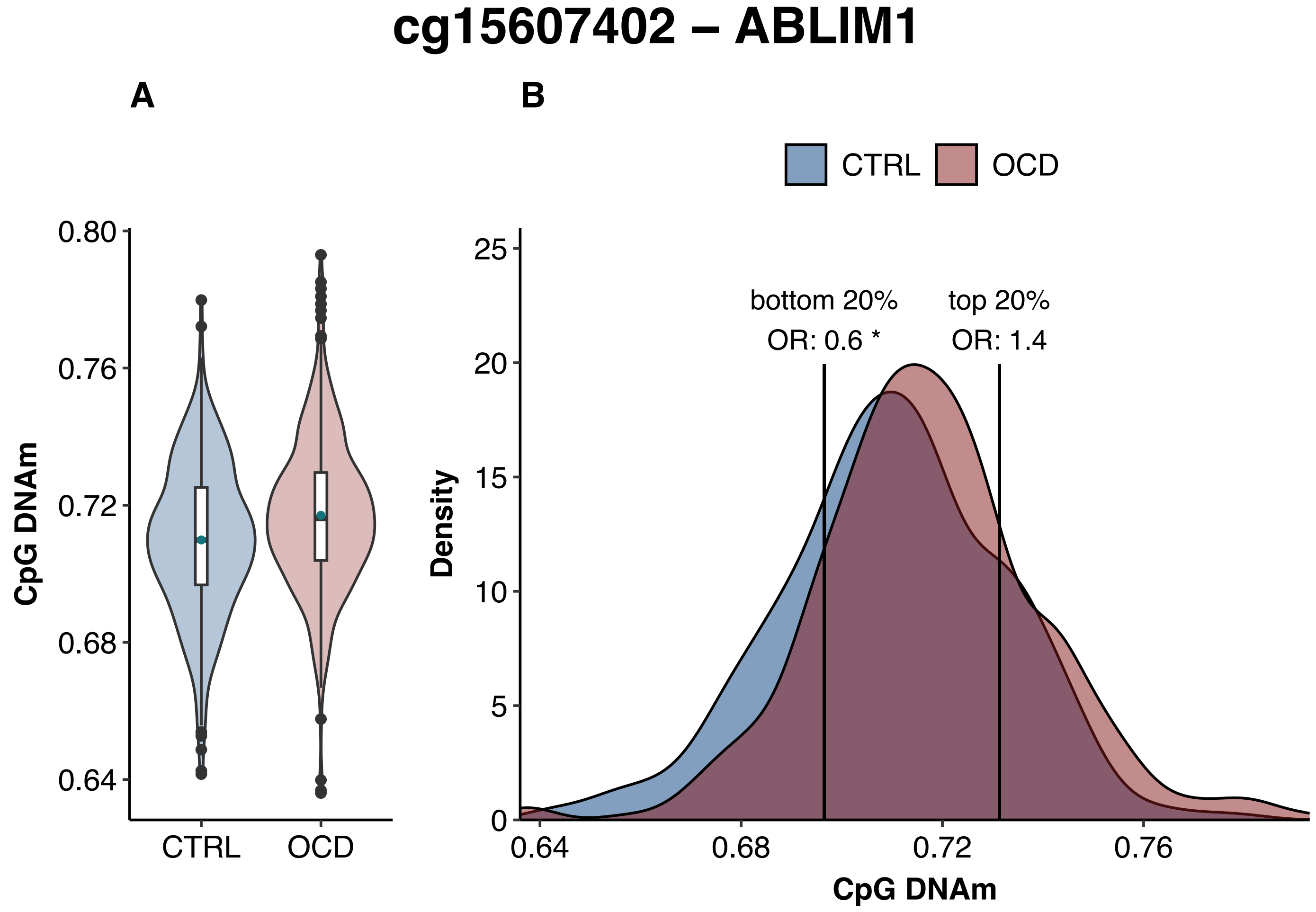

cg15607402 – ABLIM1
A
B
CTRL
OCD
0.80
0.76
0.72
0.68
0.64
CpG DNAm
CTRL
OCD
25
20
15
10
5
0
Density
bottom 20%
OR: 0.6 *
top 20%
OR: 1.4
0.64
0.68
0.72
0.76
CpG DNAm

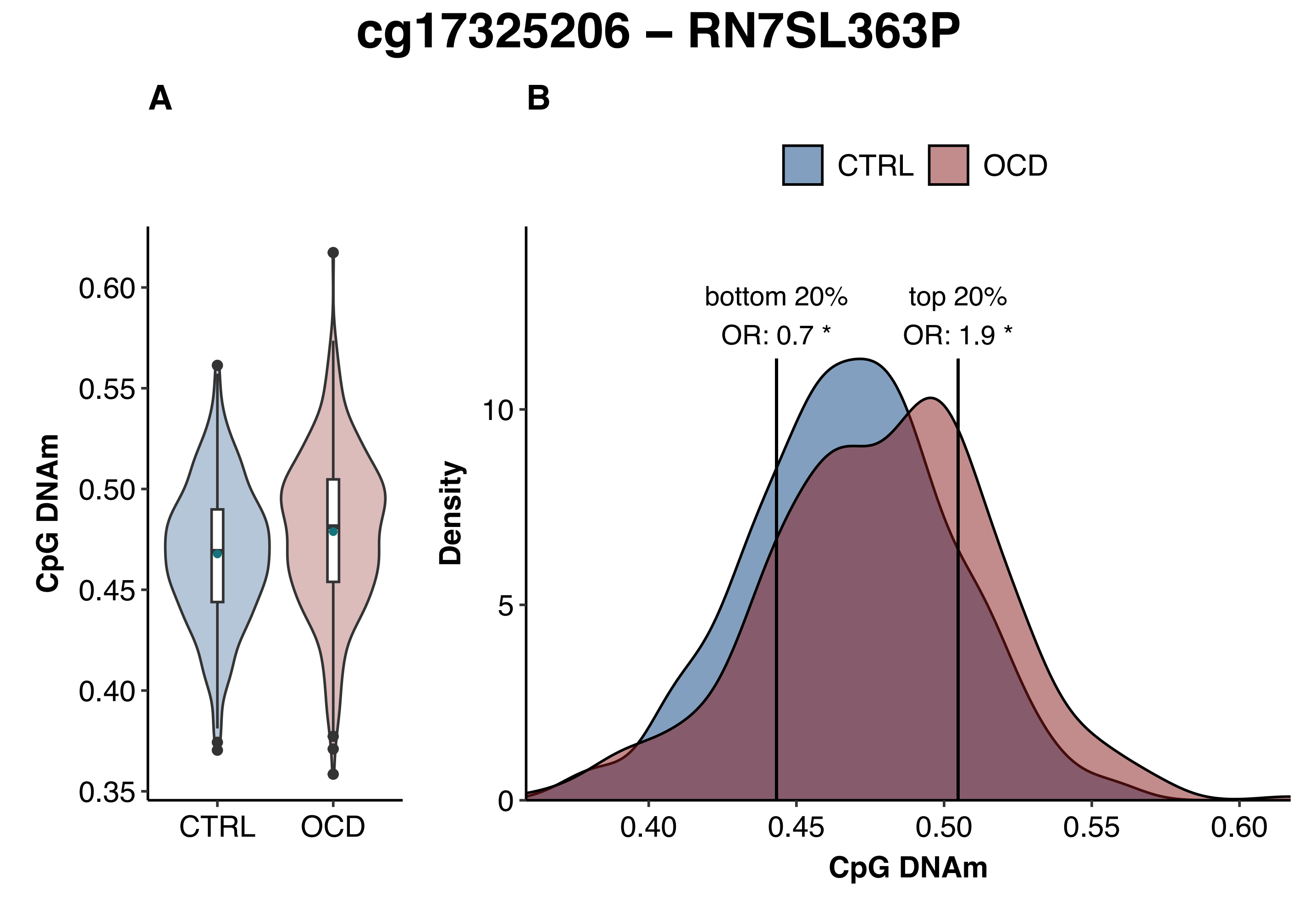
cg17325206 – RN7SL363P
A
B
CTRL
OCD
0.60
0.55
0.50
0.45
0.40
0.35
CpG DNAm
CTRL
OCD
bottom 20%
OR: 0.7 *
top 20%
OR: 1.9 *
10
5
0
Density
0.40
0.45
0.50
0.55
0.60
CpG DNAm

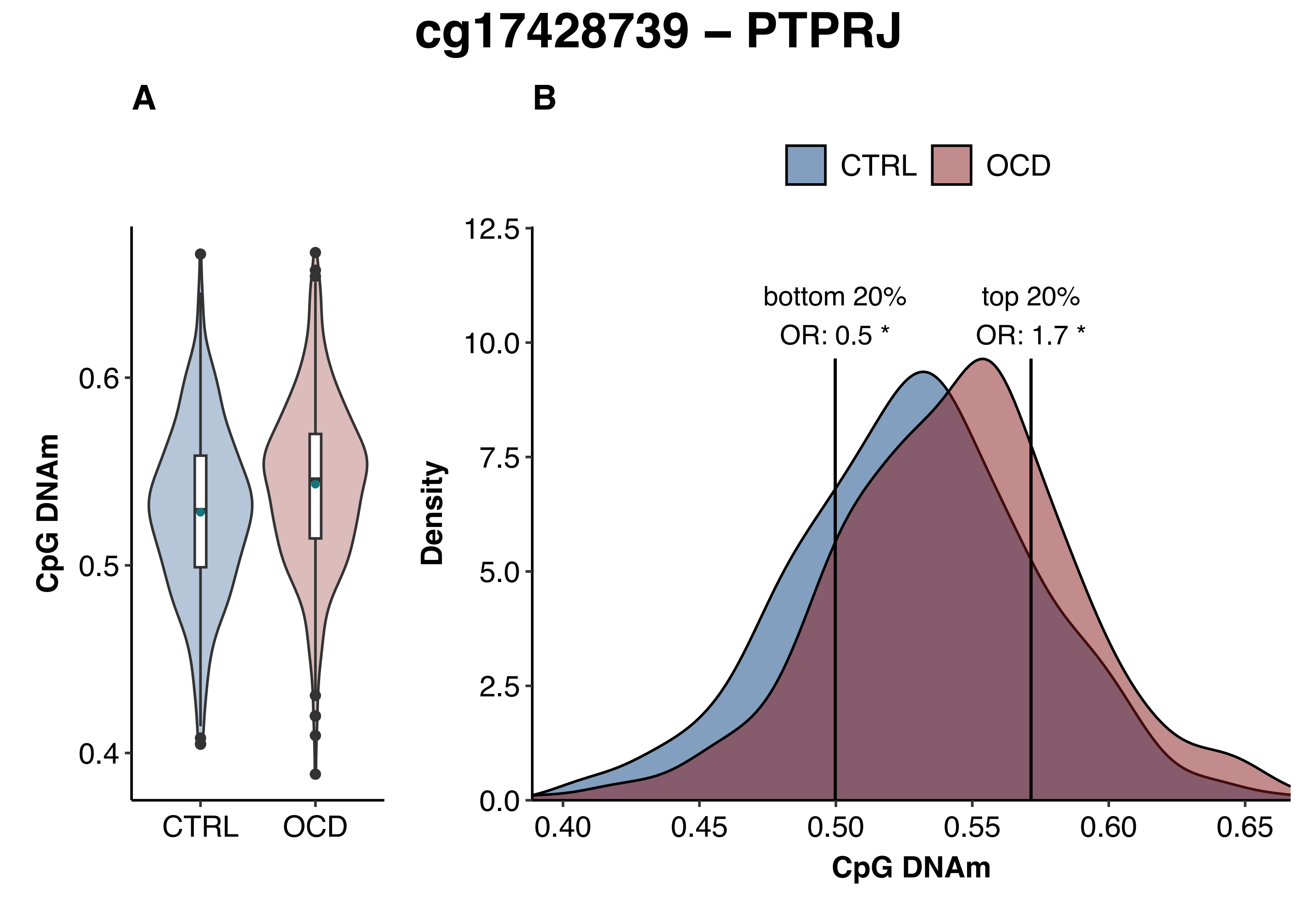
cg17428739 – PTPRJ
A
B
CTRL
OCD
CpG DNAm
0.6
0.5
0.4
CTRL
OCD
Density
12.5
10.0
7.5
5.0
2.5
0.0
bottom 20%
OR: 0.5 *
top 20%
OR: 1.7 *
0.40
0.45
0.50
0.55
0.60
0.65
CpG DNAm

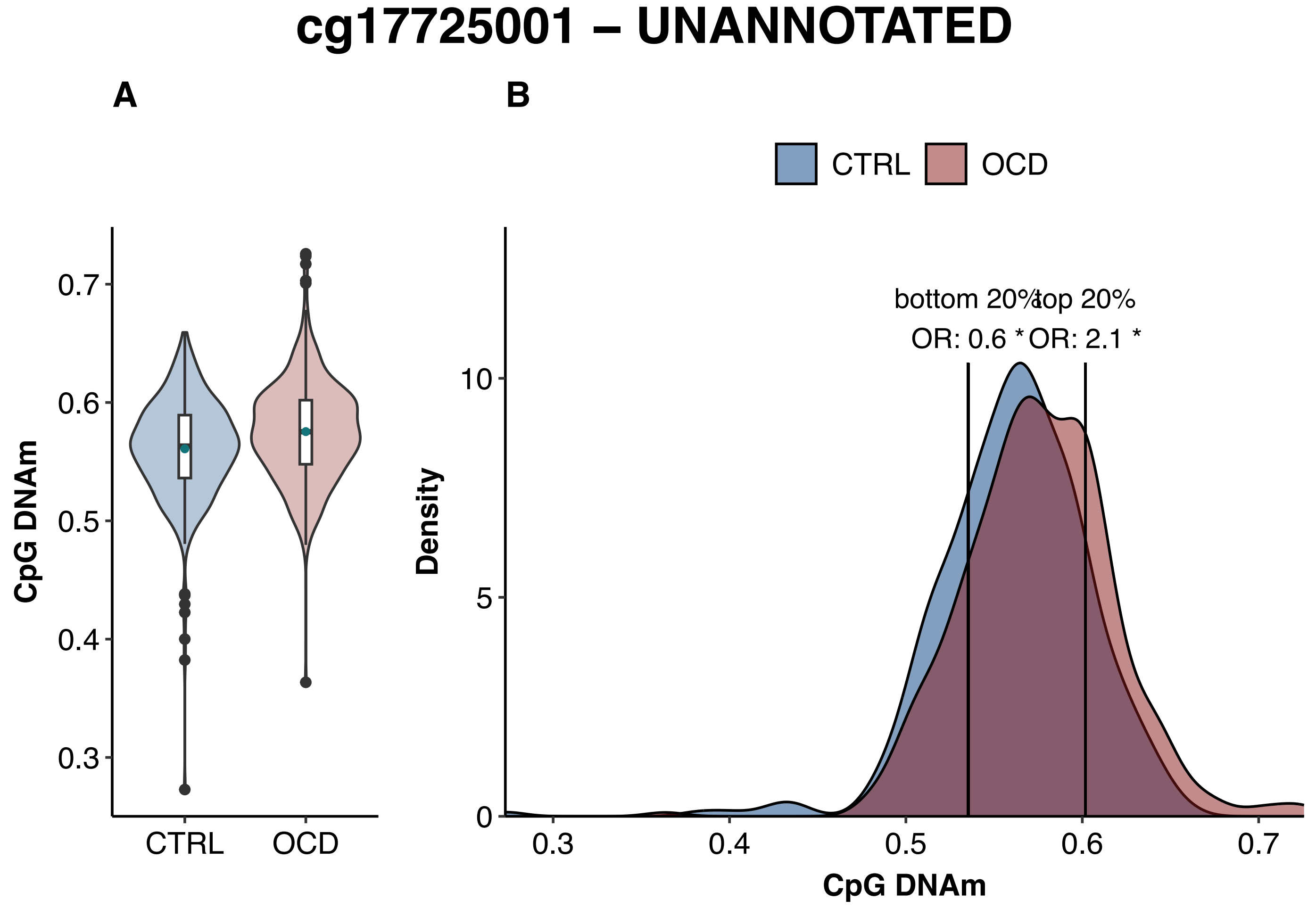
cg17725001 – UNANNOTATED
A
B
CTRL
OCD
0.7
0.6
0.5
0.4
0.3
CpG DNAm
CTRL
OCD
bottom 20%
top 20%
OR: 0.6 *
OR: 2.1 *
10
5
0
Density
0.3
0.4
0.5
0.6
0.7
CpG DNAm

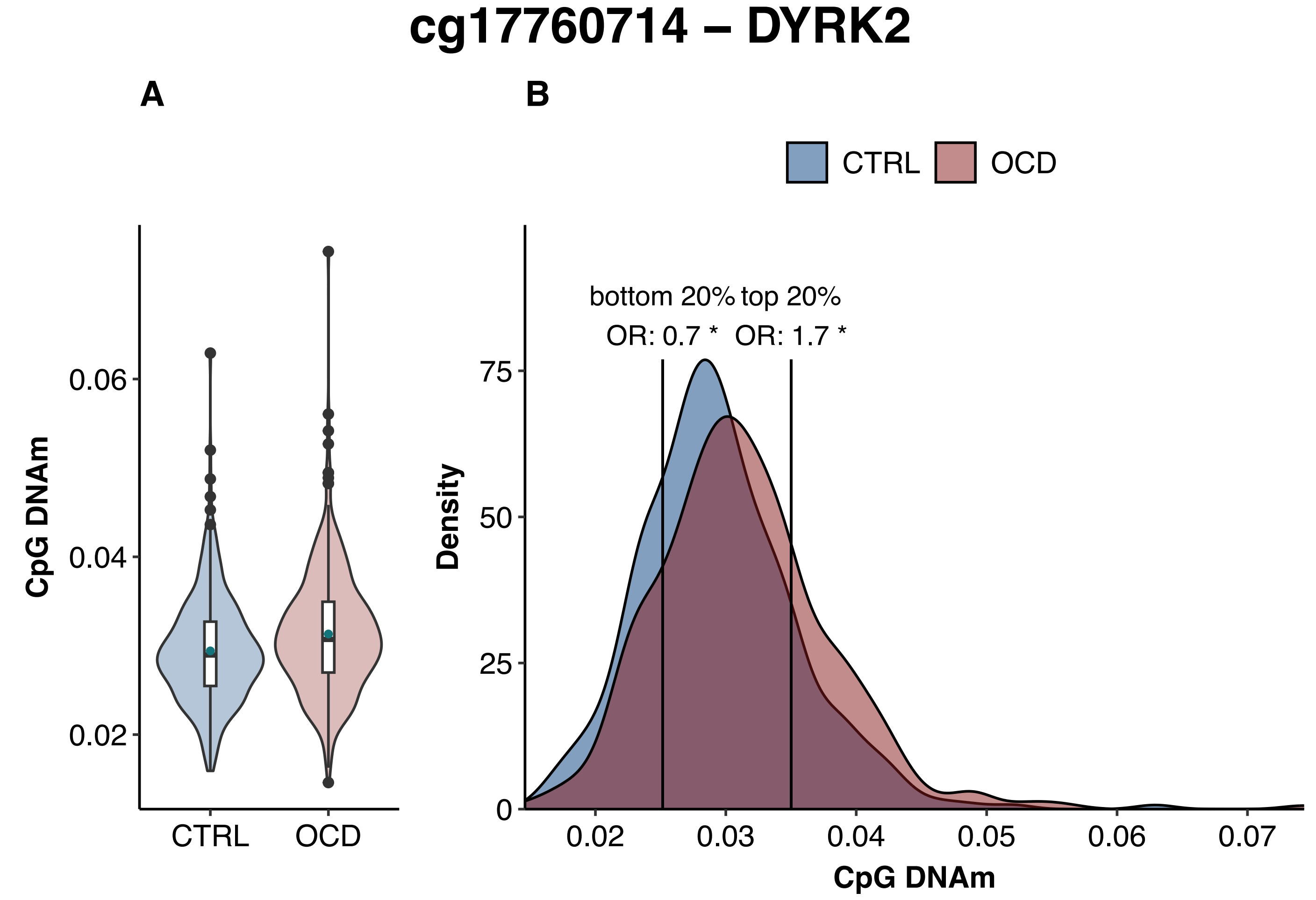

cg17760714 – DYRK2
A
B
CTRL
OCD
CpG DNAm
0.02
0.04
0.06
CTRL
OCD
bottom 20%
OR: 0.7 *
top 20%
OR: 1.7 *
Density
0
25
50
75
0.02
0.03
0.04
0.05
0.06
0.07
CpG DNAm

### cg18693051 – SBNO2

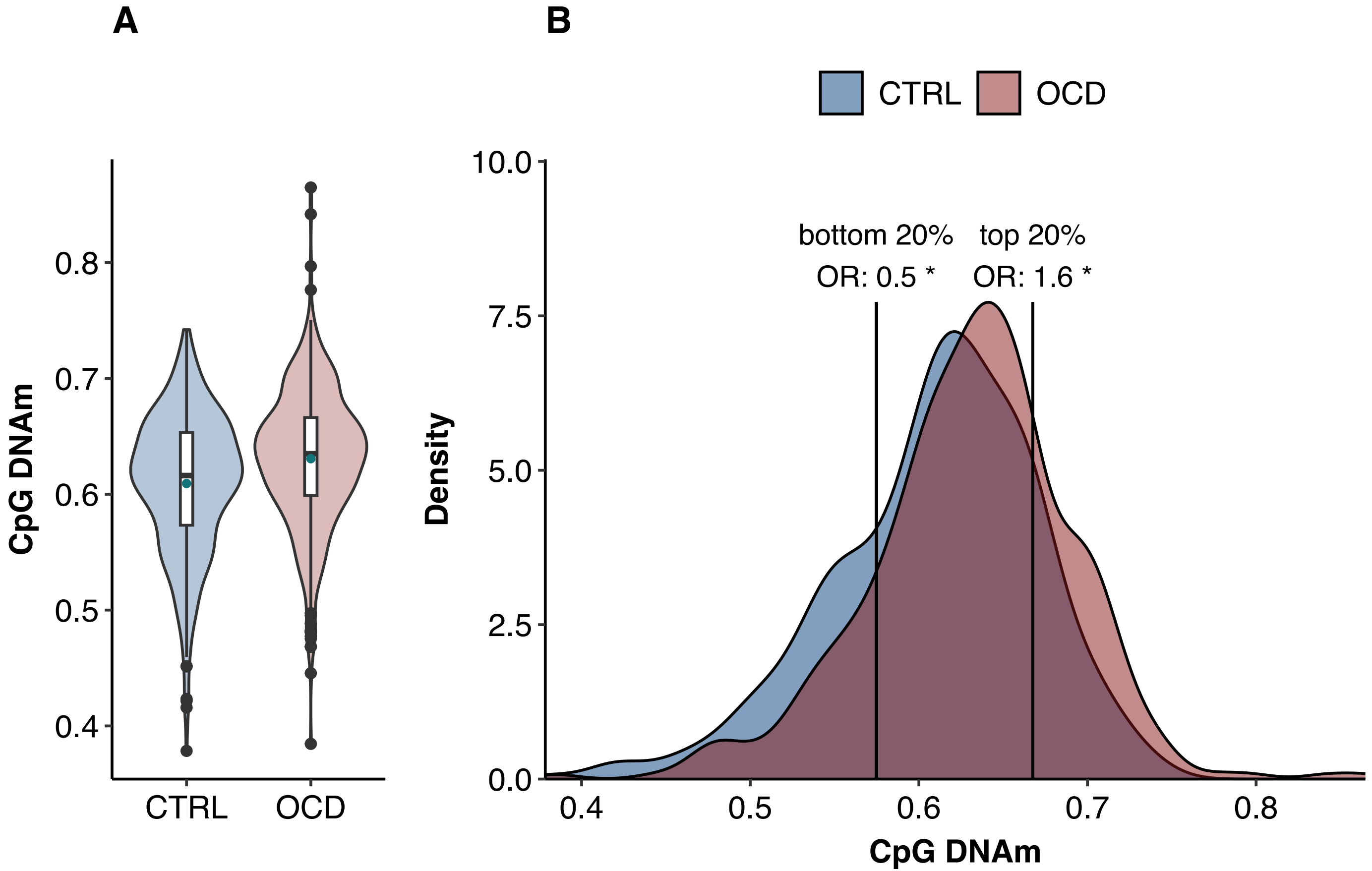

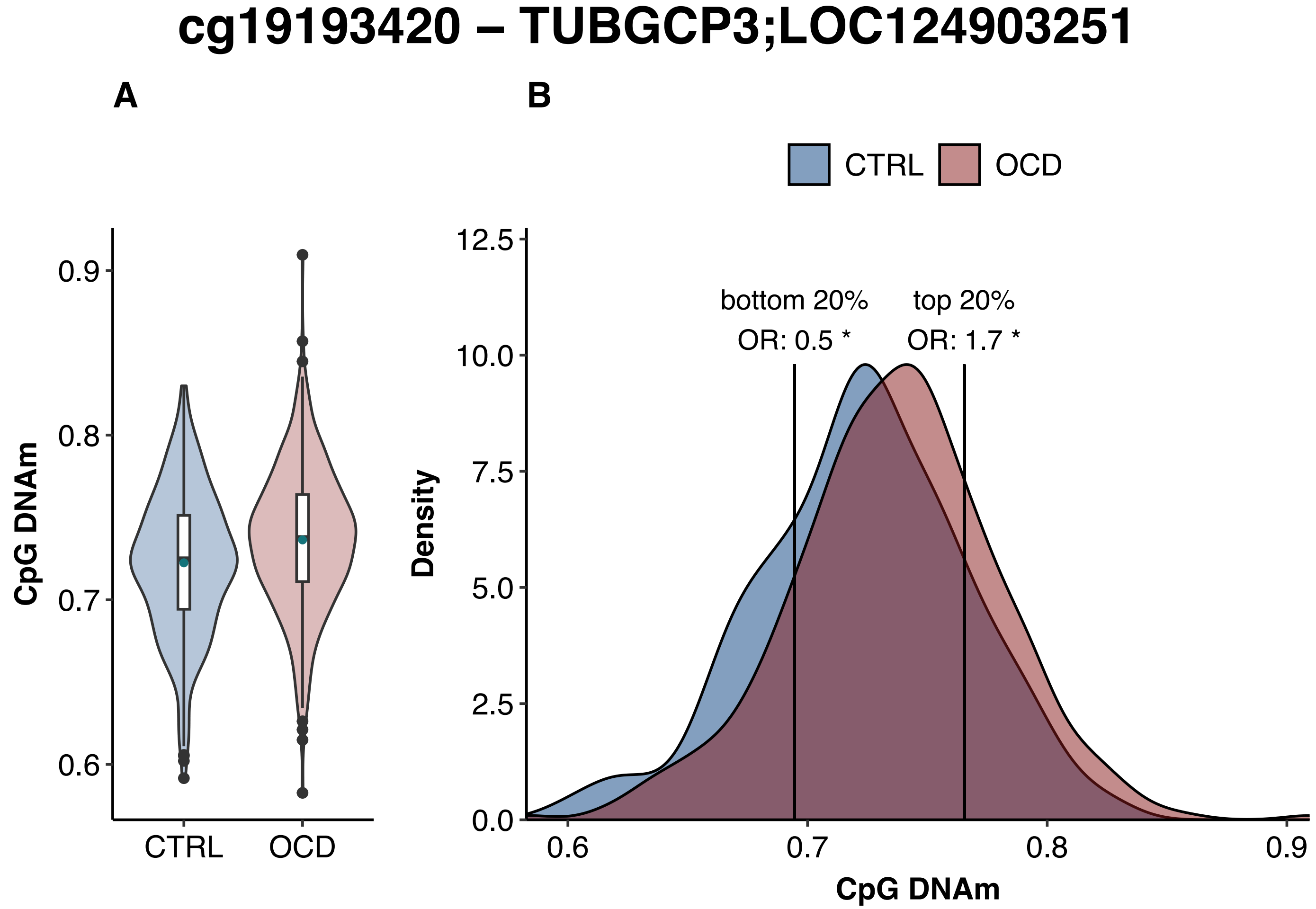
cg19193420 – TUBGCP3;LOC124903251
A
B
CTRL
OCD
CpG DNAm
0.9
0.8
0.7
0.6
CTRL
OCD
Density
12.5
10.0
7.5
5.0
2.5
0.0
bottom 20%
OR: 0.5 *
top 20%
OR: 1.7 *
0.6
0.7
0.8
0.9
CpG DNAm

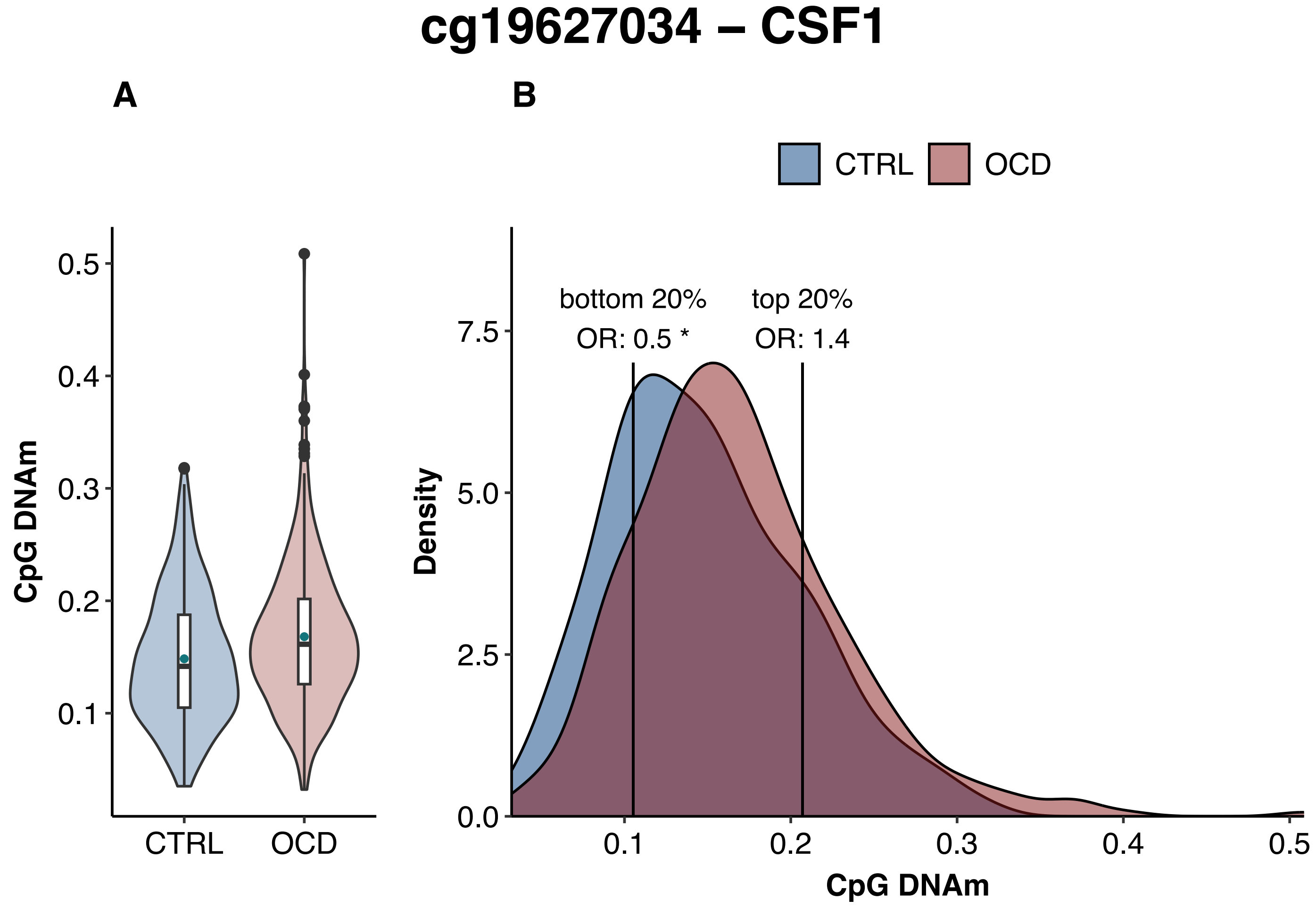
cg19627034 – CSF1
A
B
CTRL
OCD
0.5
0.4
0.3
0.2
0.1
CpG DNAm
CTRL
OCD
bottom 20%
OR: 0.5 *
top 20%
OR: 1.4
7.5
5.0
2.5
0.0
Density
0.1
0.2
0.3
0.4
0.5
CpG DNAm

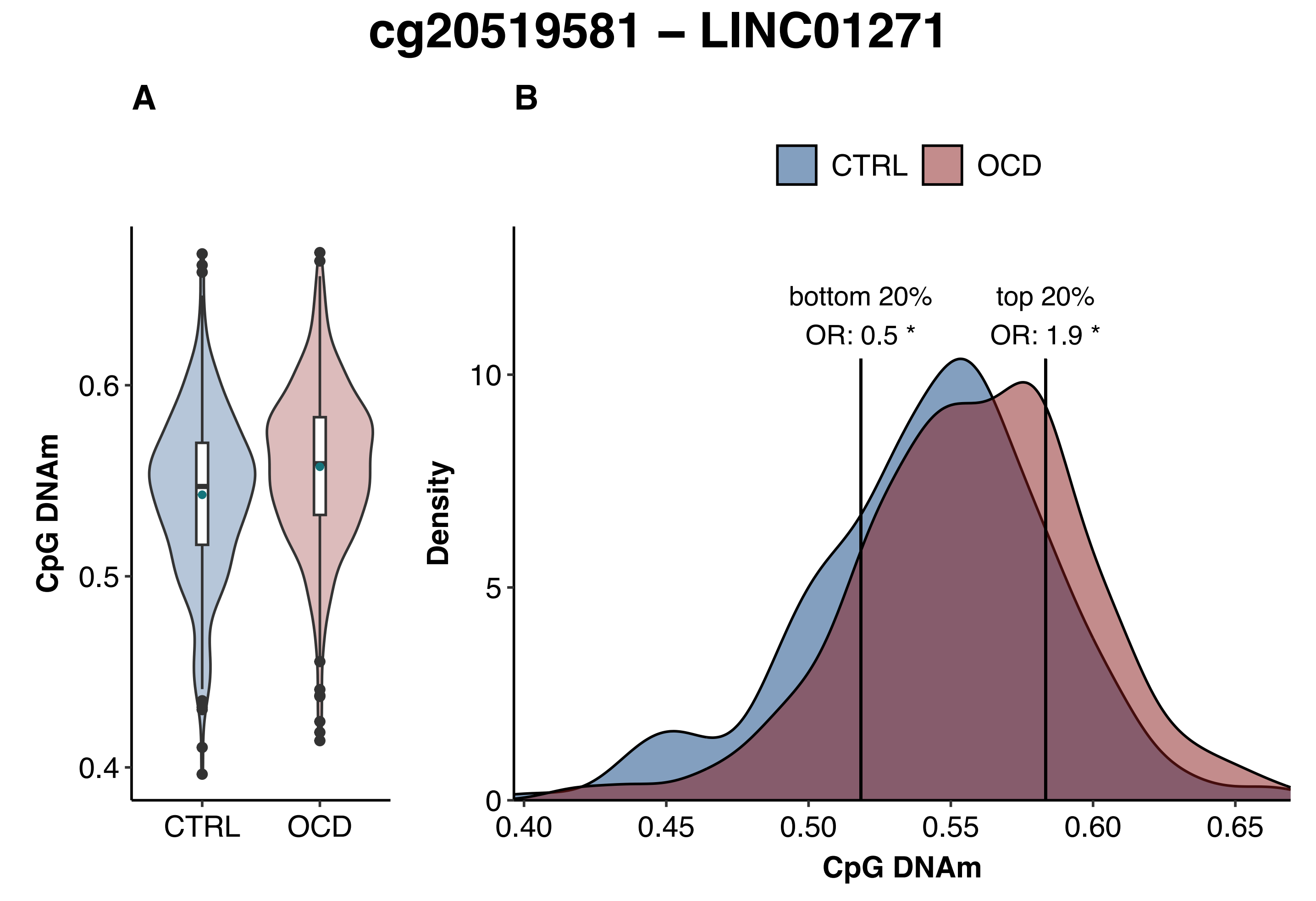

cg20519581 – LINC01271
A
B
CTRL
OCD
CpG DNAm
0.4
0.5
0.6
CTRL
OCD
bottom 20%
OR: 0.5 *
top 20%
OR: 1.9 *
Density
0
5
10
0.40
0.45
0.50
0.55
0.60
0.65
CpG DNAm

### cg20864214 – ARHGEF17

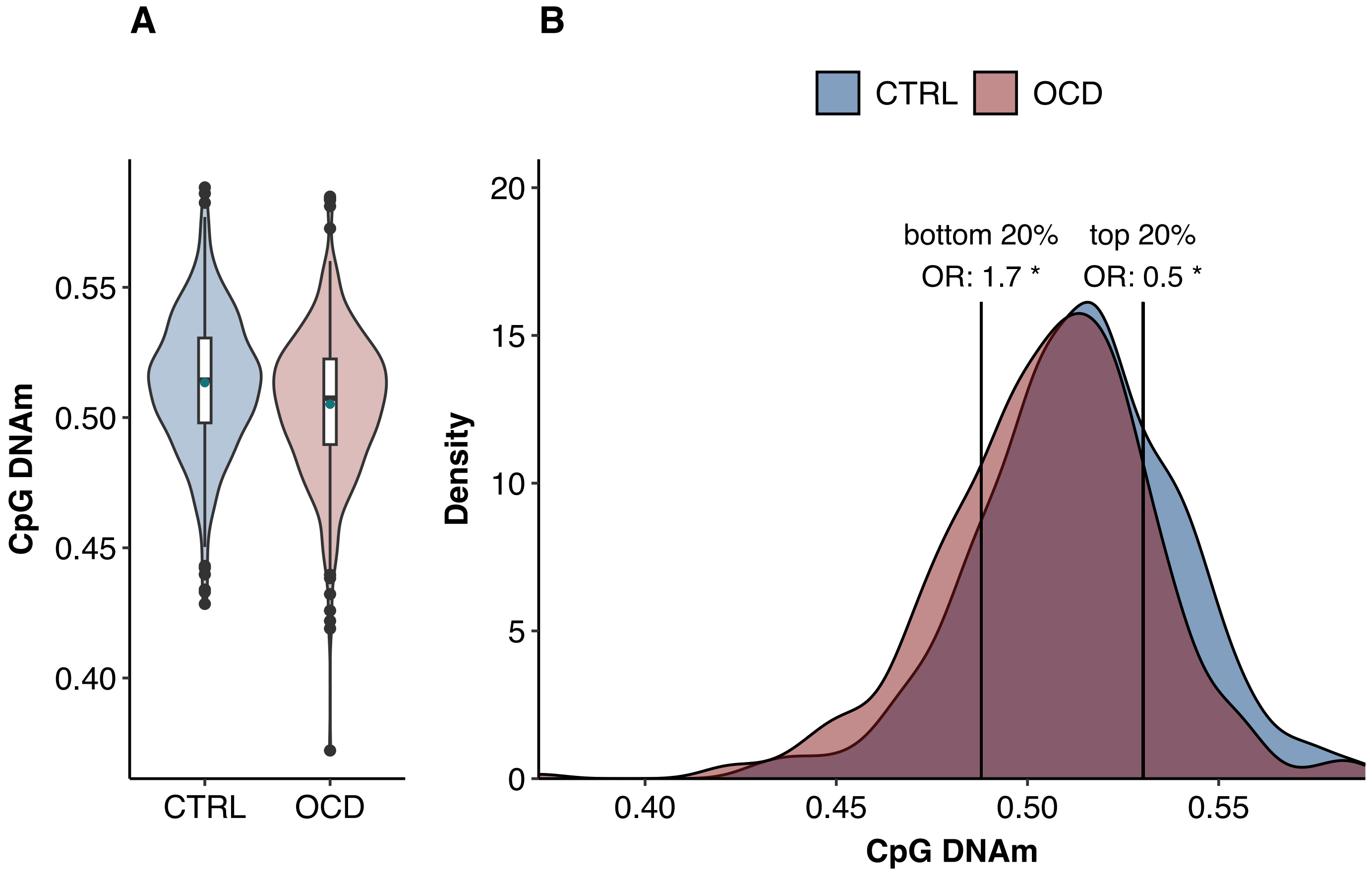

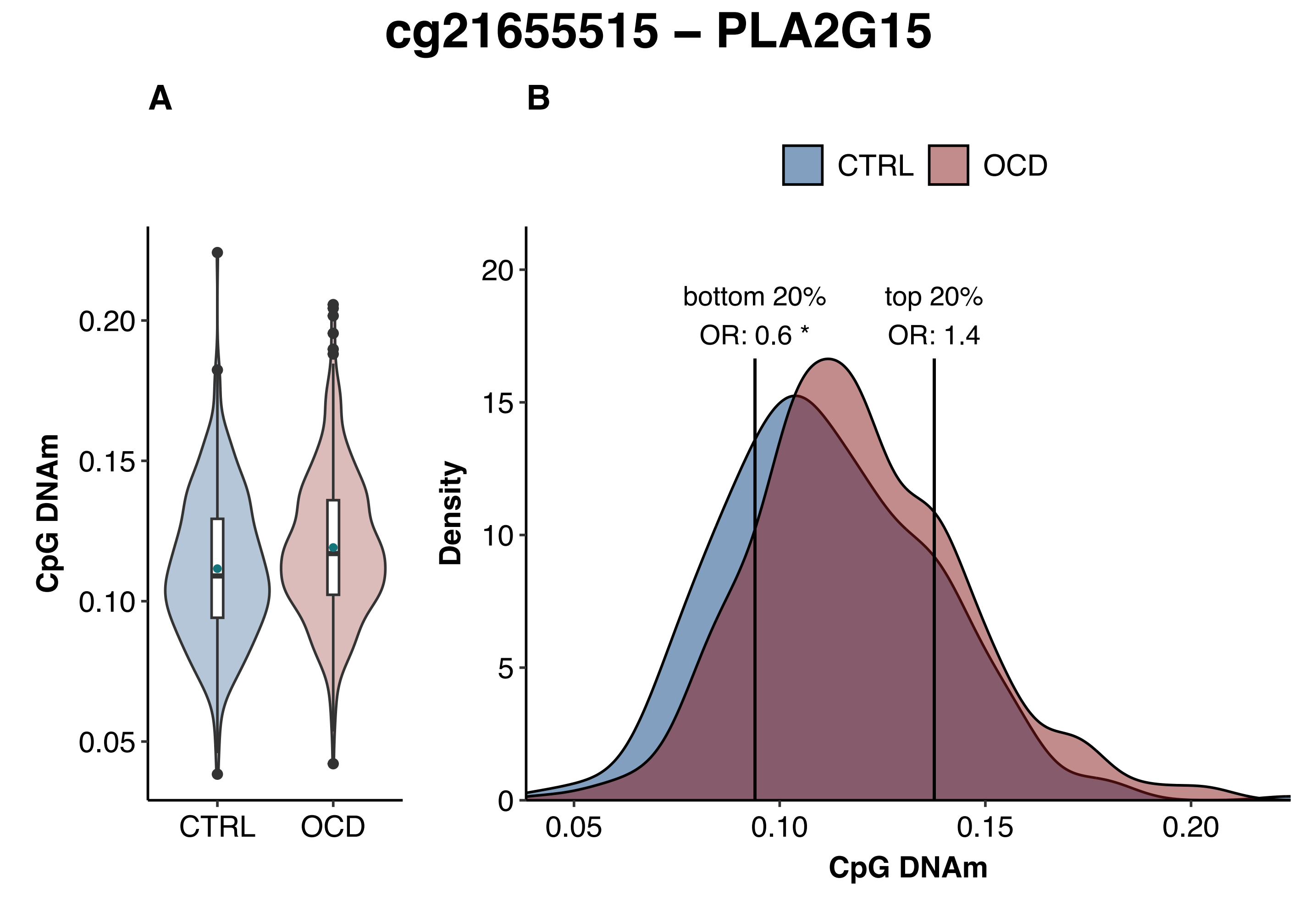
cg21655515 – PLA2G15
A
B
CTRL
OCD
CpG DNAm
0.20
0.15
0.10
0.05
CTRL
OCD
Density
20
15
10
5
0
bottom 20%
OR: 0.6 *
top 20%
OR: 1.4
0.05
0.10
0.15
0.20
CpG DNAm

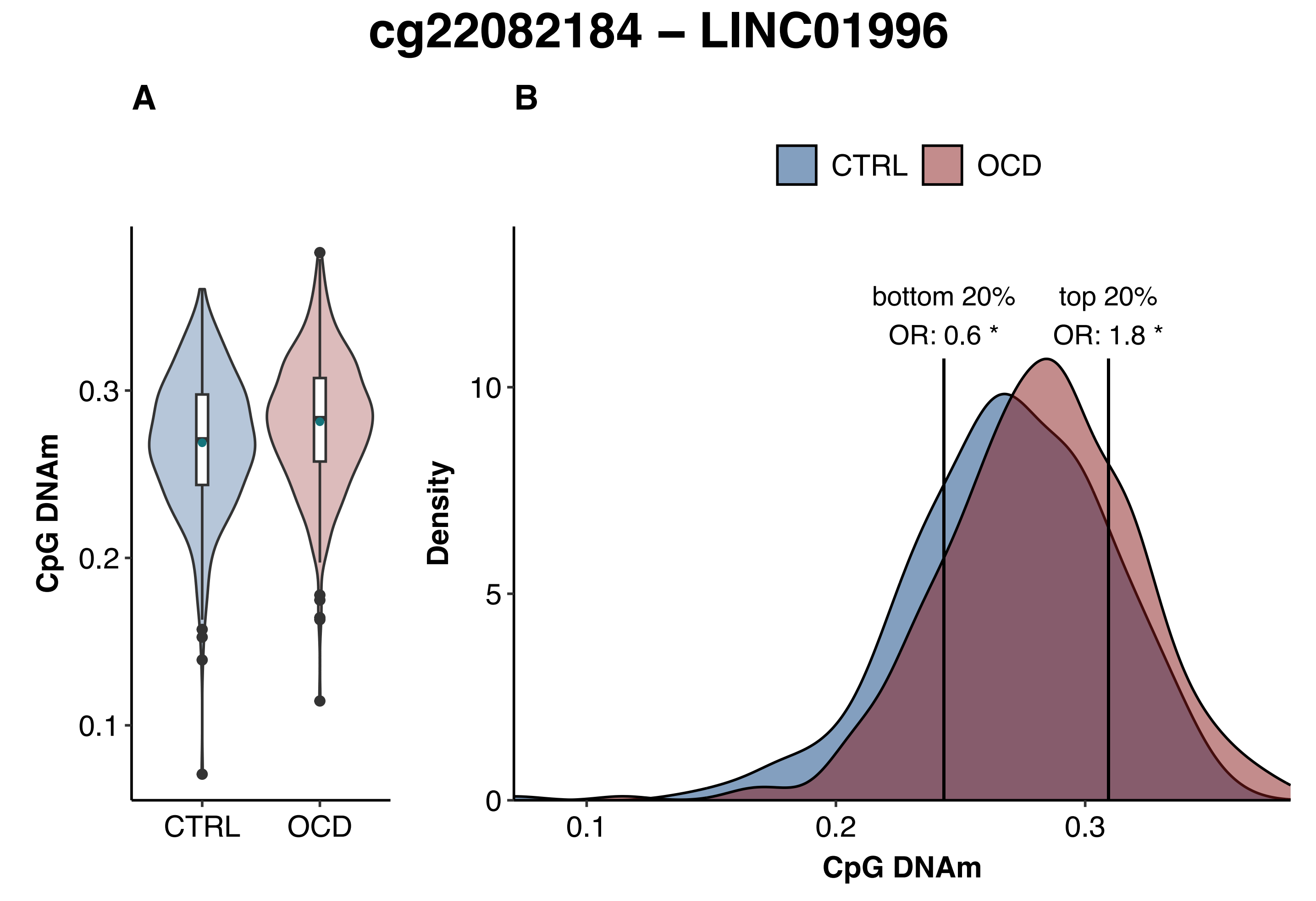
cg22082184 – LINC01996
A
B
CTRL
OCD
CpG DNAm
0.3
0.2
0.1
CTRL
OCD
bottom 20%
OR: 0.6 *
top 20%
OR: 1.8 *
Density
10
5
0
0.1
0.2
0.3
CpG DNAm

### cg22325408 – ABCA7

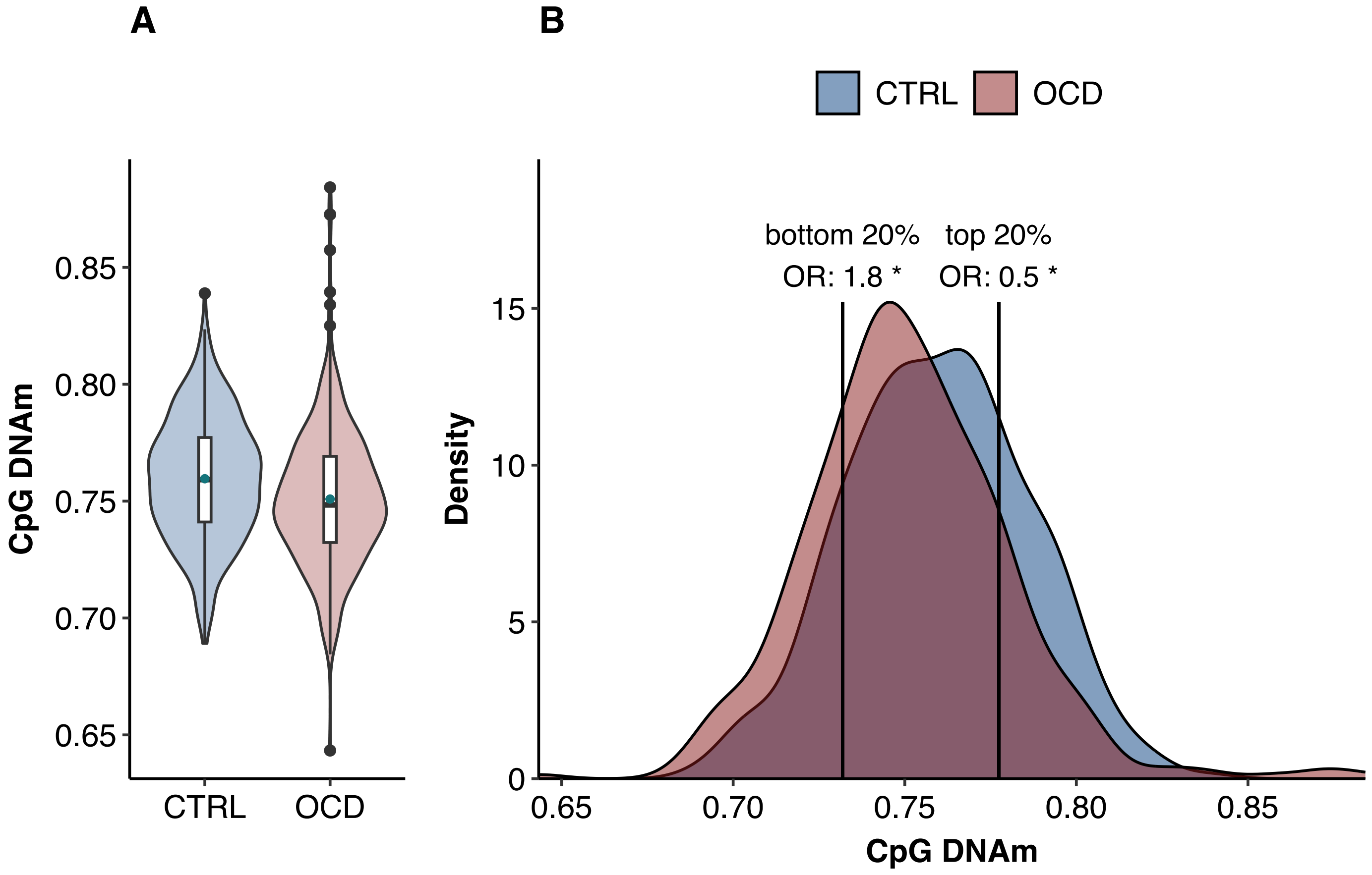

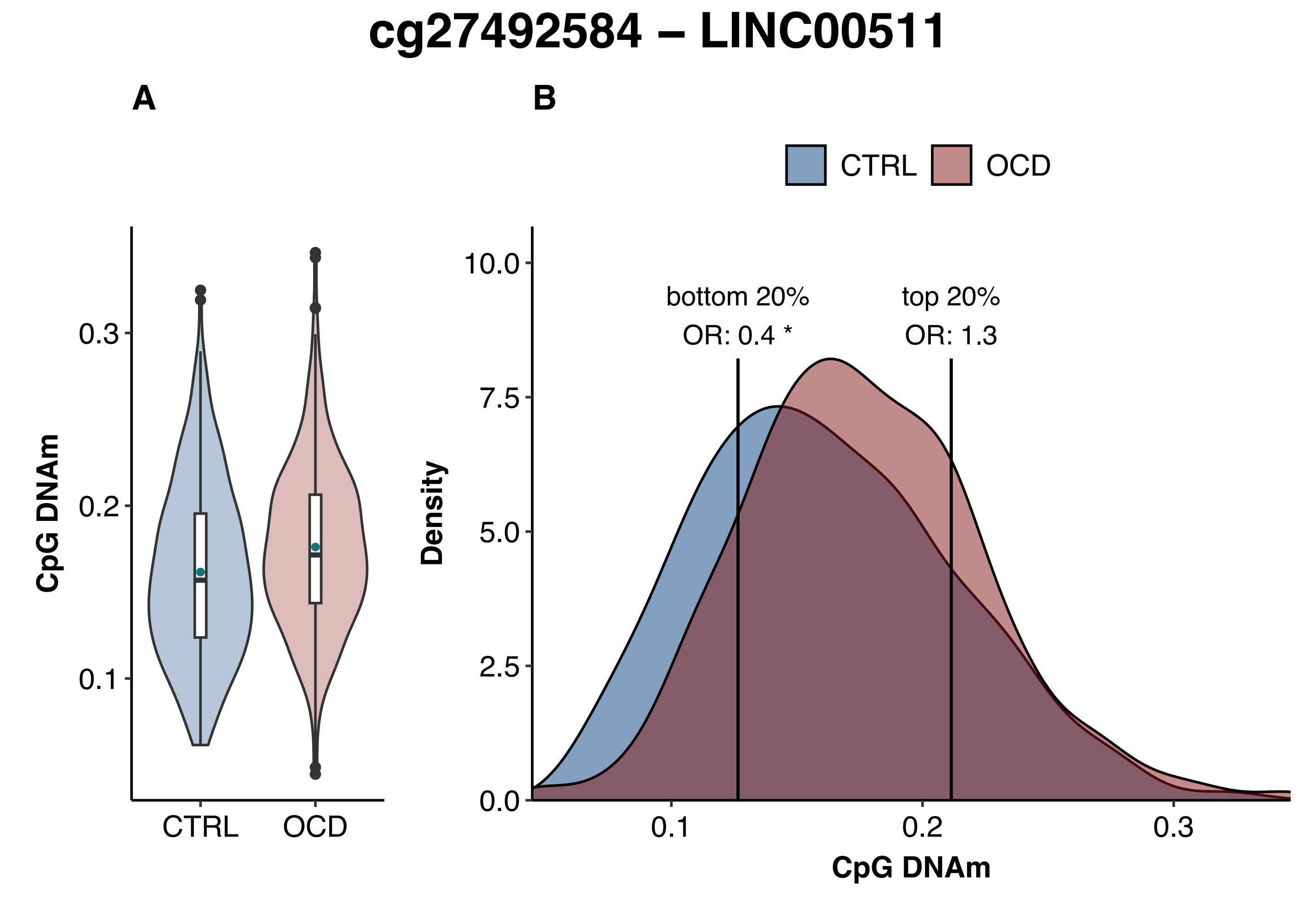
cg27492584 – LINC00511
A
B
CTRL
OCD
CpG DNAm
0.3
0.2
0.1
Density
10.0
7.5
5.0
2.5
0.0
bottom 20%
OR: 0.4 *
top 20%
OR: 1.3

### sex-specific analysis in females

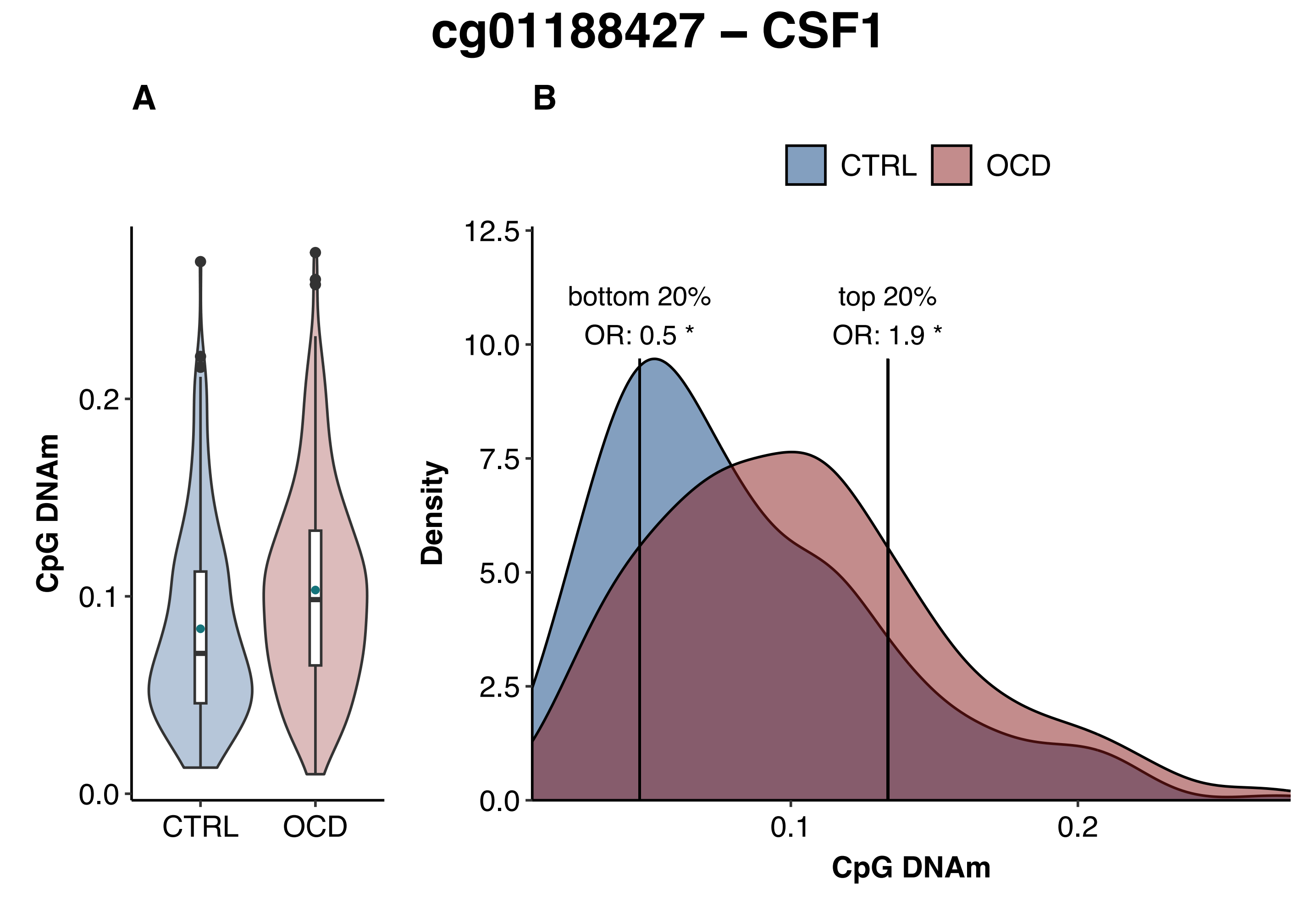
cg01188427 – CSF1
A
B
CTRL
OCD
CpG DNAm
0.0
0.1
0.2
CTRL
OCD
bottom 20%
OR: 0.5 *
top 20%
OR: 1.9 *
Density
0.0
2.5
5.0
7.5
10.0
12.5
0.1
0.2
CpG DNAm
