## Supplementary File 3 for "Methylome-Wide Association Study of Obsessive-Compulsive Disorder"

### Differentially methylated regions (DMRs)

A) Box-violin plots showing DNA methylation differences between individuals with OCD and controls.

B) Density plots of methylation levels, with OCD odds ratios (ORs) calculated for the highest and lowest 20% of methylation values (indicated by vertical lines). Significant ORs are marked with an asterisk (*).

C) Difference in median DNA methylation between individuals with OCD and healthy controls for each CpG site (red dots) within the DMR.

chr: chromosome, CTRL: healthy control, DMR: differentially methylated region. DNAm: DNA methylation, OCD: obsessive-compulsive disorder.

**sex-stratified meta-analysis**

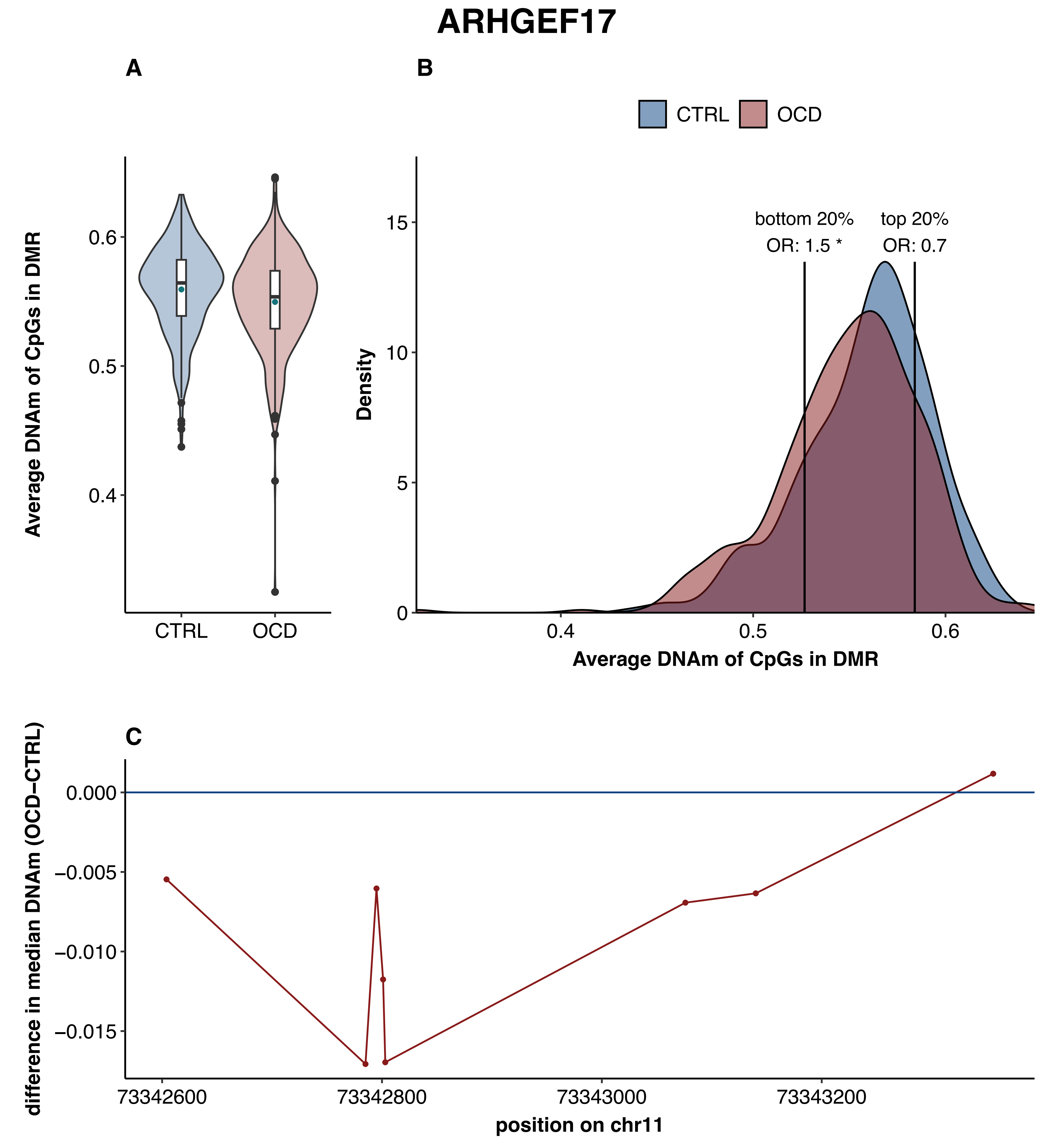
ARHGEF17
A
B
C
CTRL
OCD
Average DNAm of CpGs in DMR
0.6
0.5
0.4
Density
15
10
5
0
bottom 20%
OR: 1.5 *
top 20%
OR: 0.7
difference in median DNAm (OCD−CTRL)
0.000
−0.005
−0.010
−0.015
73342600
73342800
73343000
73343200
position on chr11

### B3GALT4

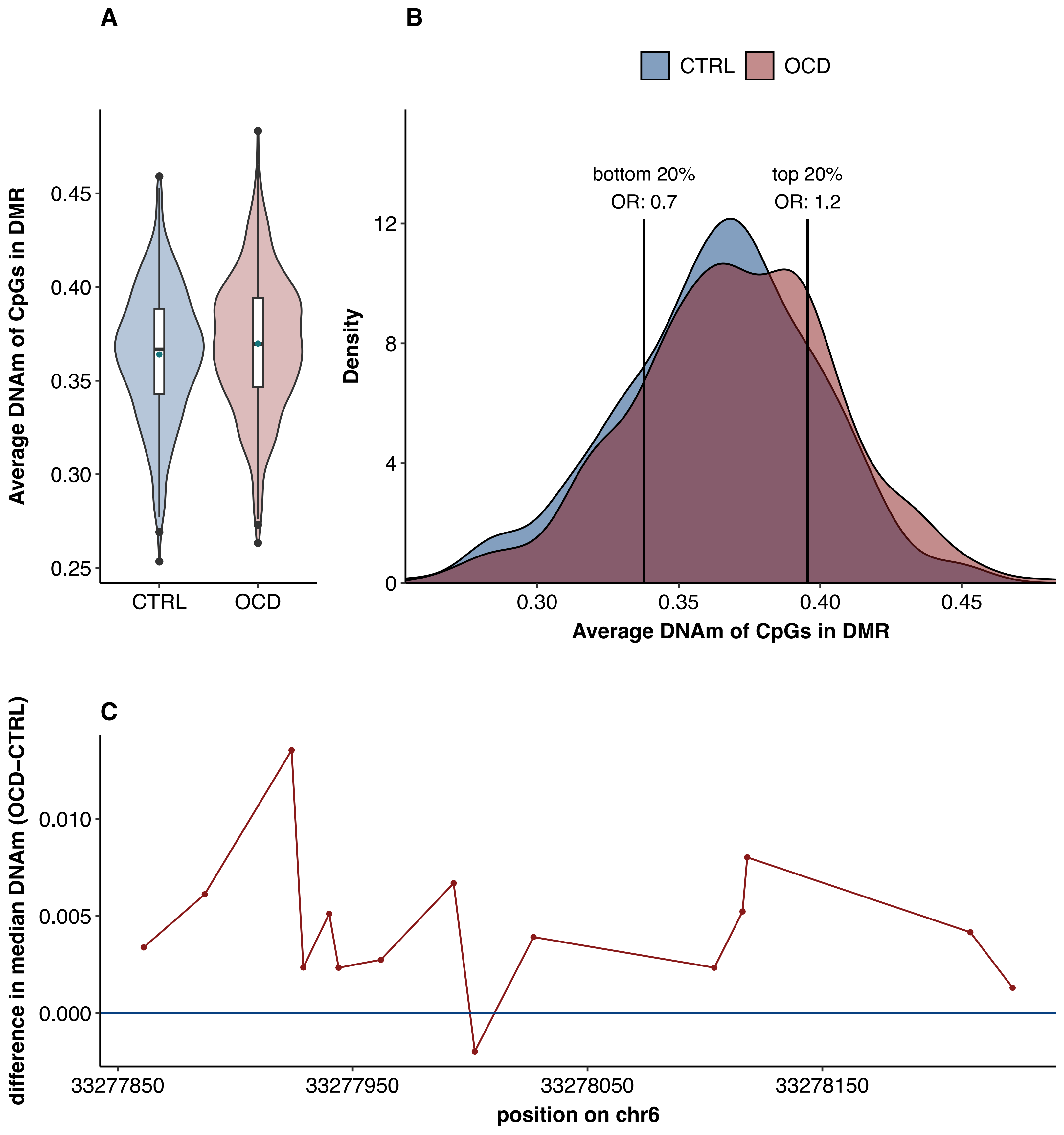
A
B
CTRL
OCD
Average DNAm of CpGs in DMR
0.25
0.30
0.35
0.40
0.45
bottom 20%
OR: 0.7
top 20%
OR: 1.2
Density
0
4
8
12
C
difference in median DNAm (OCD−CTRL)
0.000
0.005
0.010
33277850
33277950
33278050
33278150
position on chr6

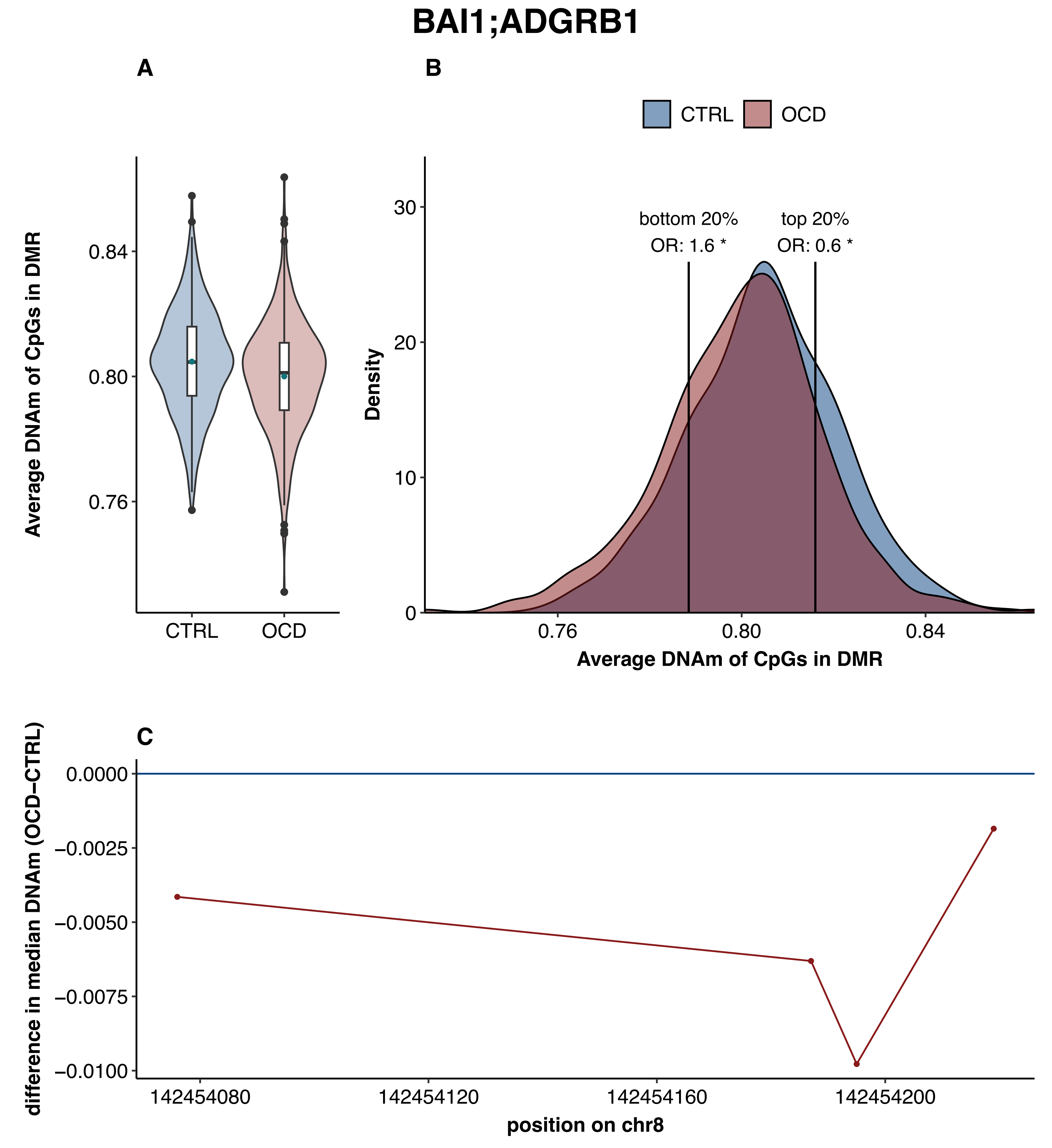
BAI1;ADGRB1
A
B
C
CTRL
OCD
Average DNAm of CpGs in DMR
0.84
0.80
0.76
bottom 20%
OR: 1.6 *
top 20%
OR: 0.6 *
Density
30
20
10
0
difference in median DNAm (OCD−CTRL)
0.0000
−0.0025
−0.0050
−0.0075
−0.0100
142454080
142454120
142454160
142454200
position on chr8

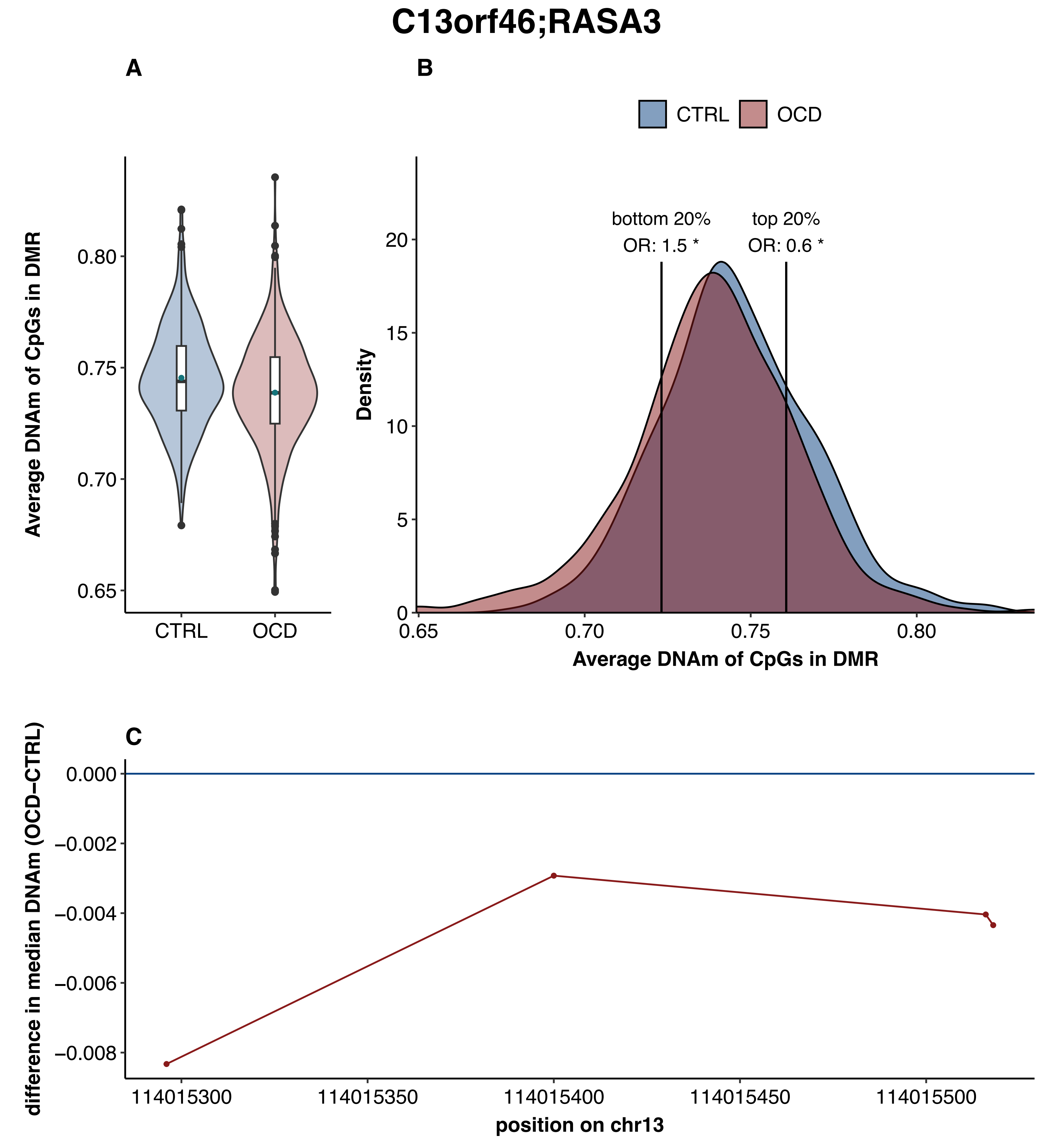
C13orf46;RASA3
A
B
C
CTRL
OCD
Average DNAm of CpGs in DMR
0.65
0.70
0.75
0.80
bottom 20%
OR: 1.5 *
top 20%
OR: 0.6 *
Density
0
5
10
15
20
difference in median DNAm (OCD−CTRL)
0.000
−0.002
−0.004
−0.006
−0.008
114015300
114015350
114015400
114015450
114015500
position on chr13

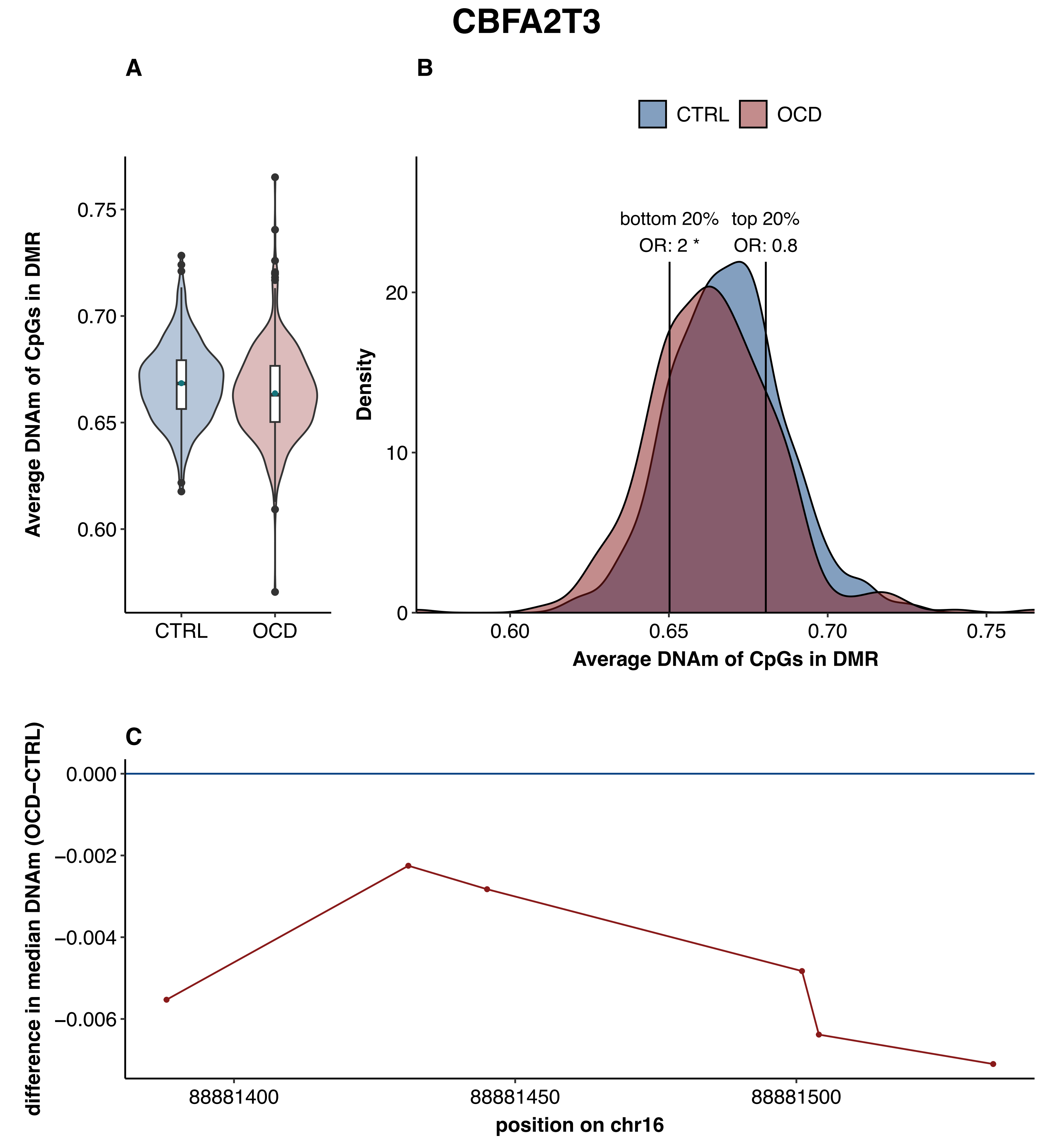
CBFA2T3
A
B
C
CTRL
OCD
bottom 20%
OR: 2 *
top 20%
OR: 0.8
Average DNAm of CpGs in DMR
Density
difference in median DNAm (OCD−CTRL)
position on chr16
0.75
0.70
0.65
0.60
0.000
−0.002
−0.004
−0.006
88881400
88881450
88881500
20
10
0

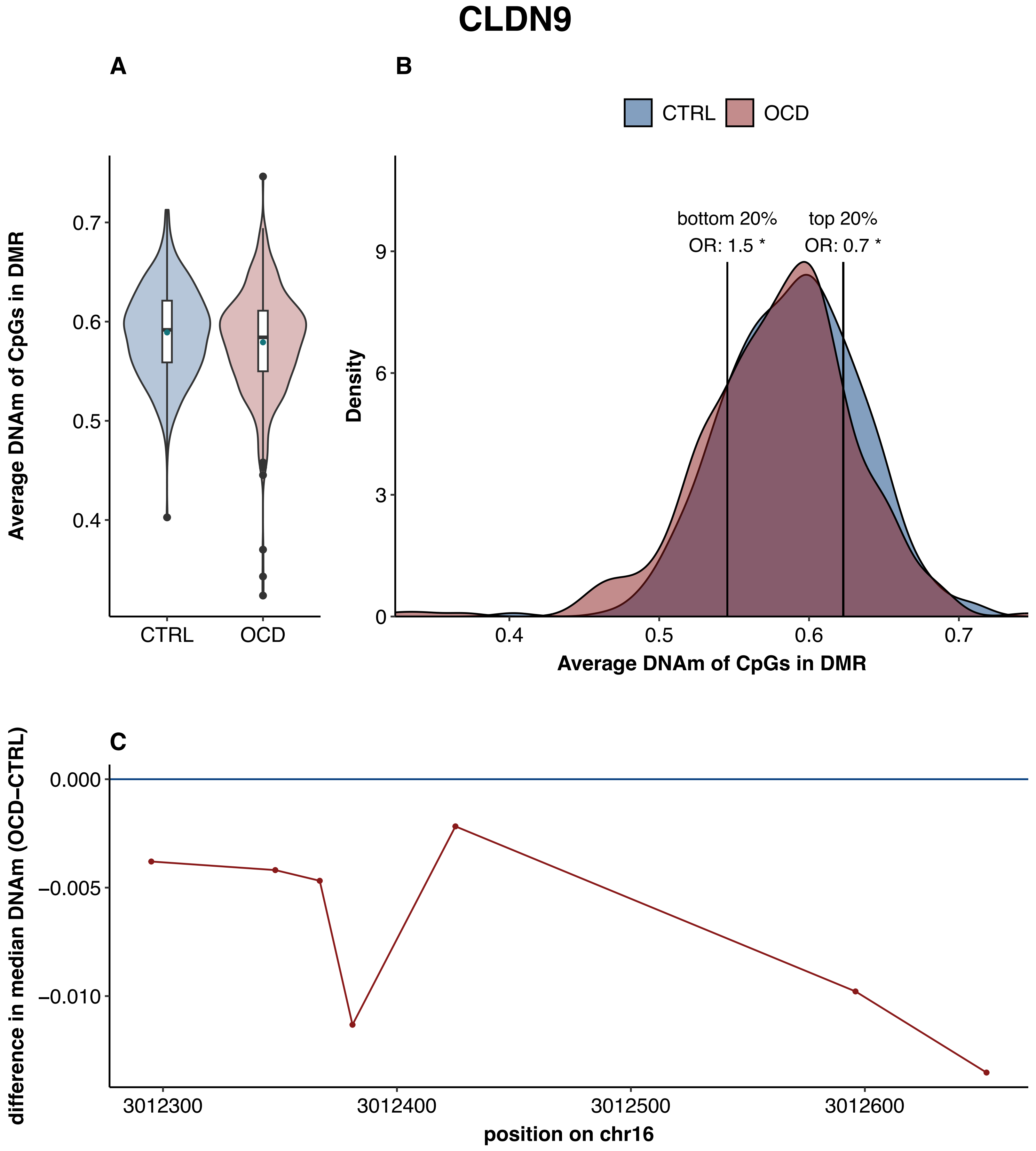

CLDN9
A
B
C
CTRL
OCD
Average DNAm of CpGs in DMR
0.4
0.5
0.6
0.7
bottom 20%
OR: 1.5 *
top 20%
OR: 0.7 *
Density
0
3
6
9
difference in median DNAm (OCD−CTRL)
0.000
−0.005
−0.010
3012300
3012400
3012500
3012600
position on chr16

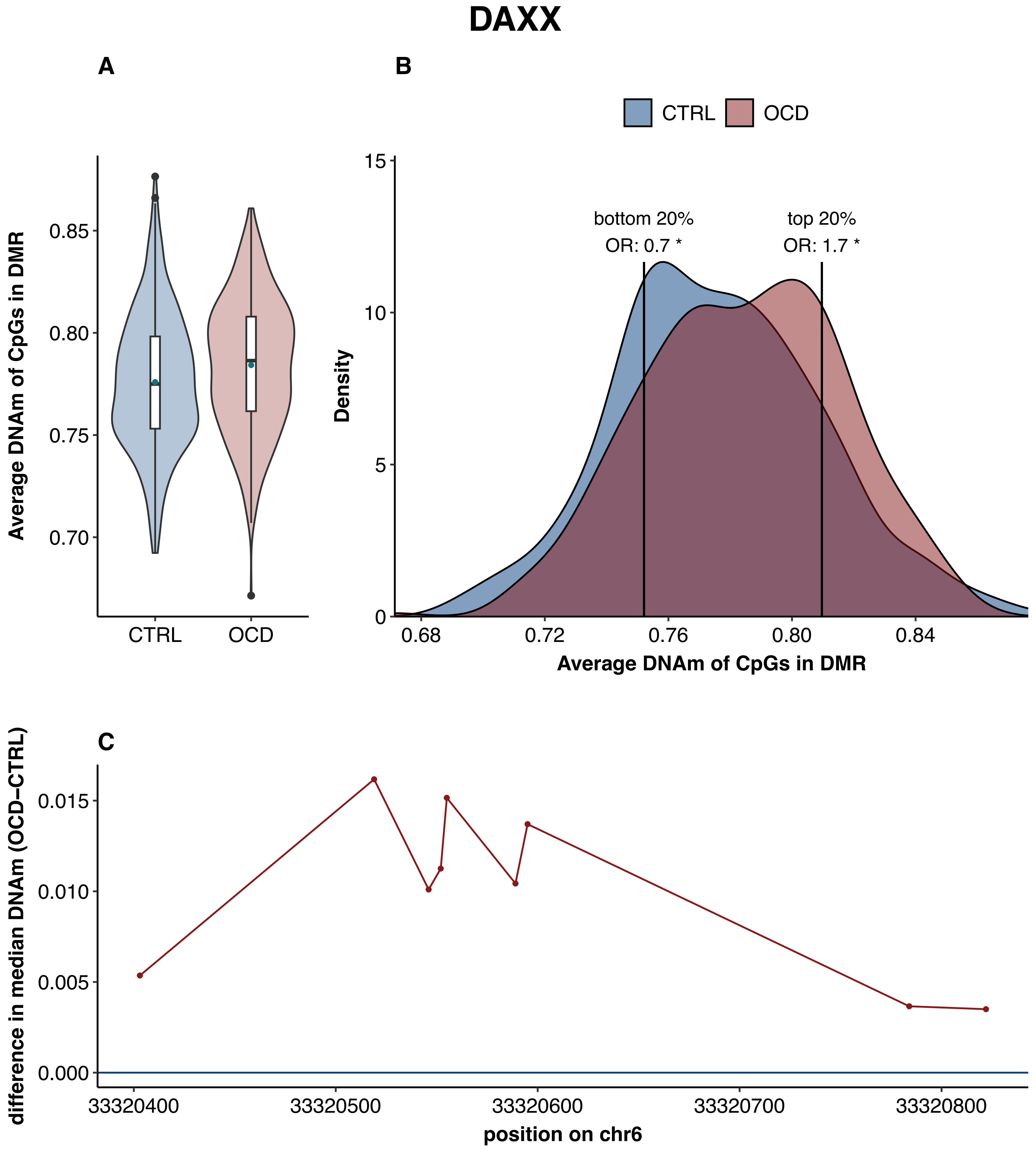

DAXX
A
B
CTRL
OCD
Average DNAm of CpGs in DMR
0.85
0.80
0.75
0.70
CTRL
OCD
Density
15
10
5
0
bottom 20%
OR: 0.7 *
top 20%
OR: 1.7 *
0.68
0.72
0.76
0.80
0.84
Average DNAm of CpGs in DMR
C
difference in median DNAm (OCD−CTRL)
0.015
0.010
0.005
0.000
33320400
33320500
33320600
33320700
33320800
position on chr6

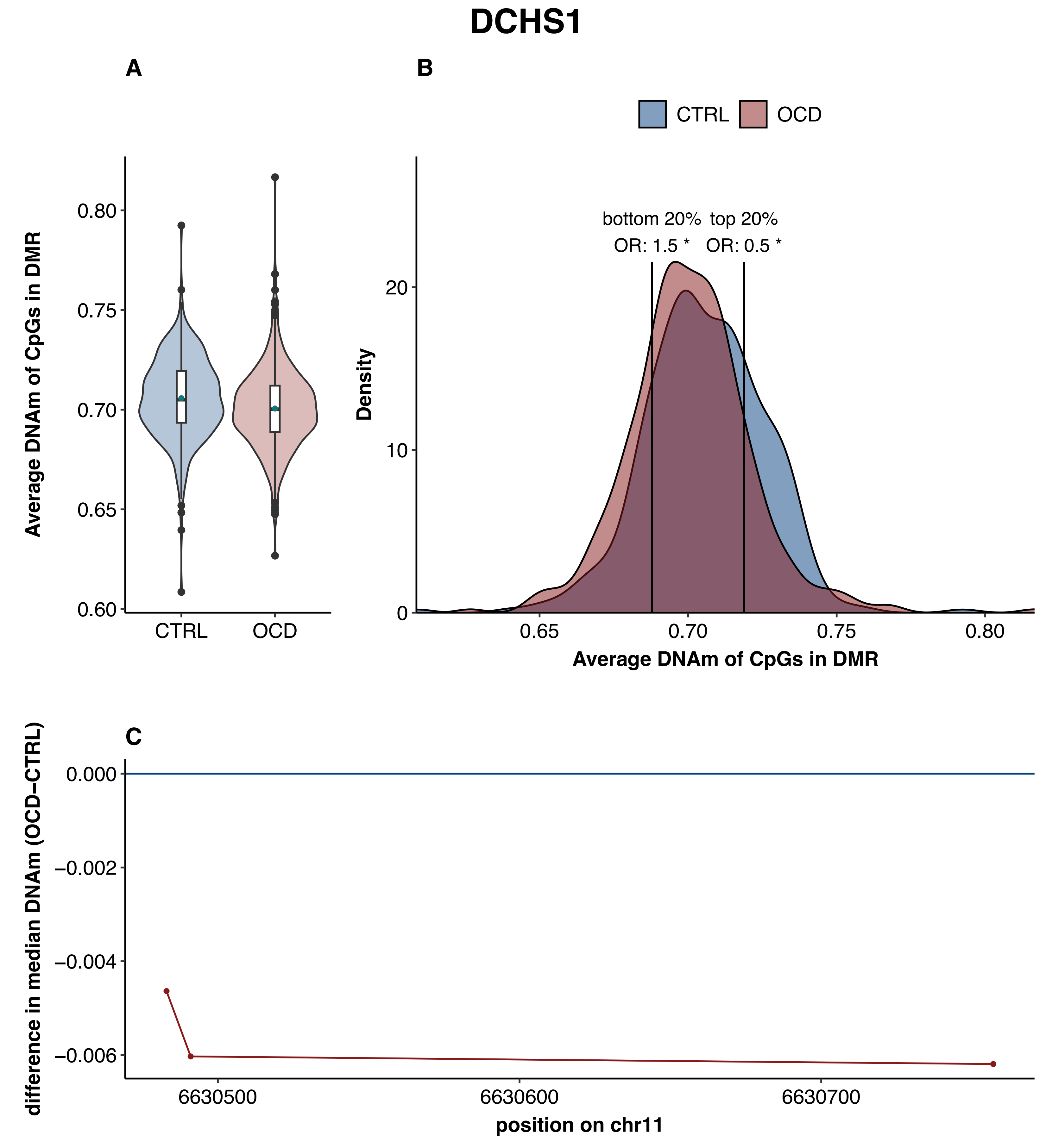
DCHS1
A
B
CTRL
OCD
Average DNAm of CpGs in DMR
0.80
0.75
0.70
0.65
0.60
CTRL
OCD
bottom 20%
OR: 1.5 *
top 20%
OR: 0.5 *
Density
20
10
0
0.65
0.70
0.75
0.80
Average DNAm of CpGs in DMR
C
difference in median DNAm (OCD−CTRL)
0.000
−0.002
−0.004
−0.006
6630500
6630600
6630700
position on chr11

### FAM120B;DLL1

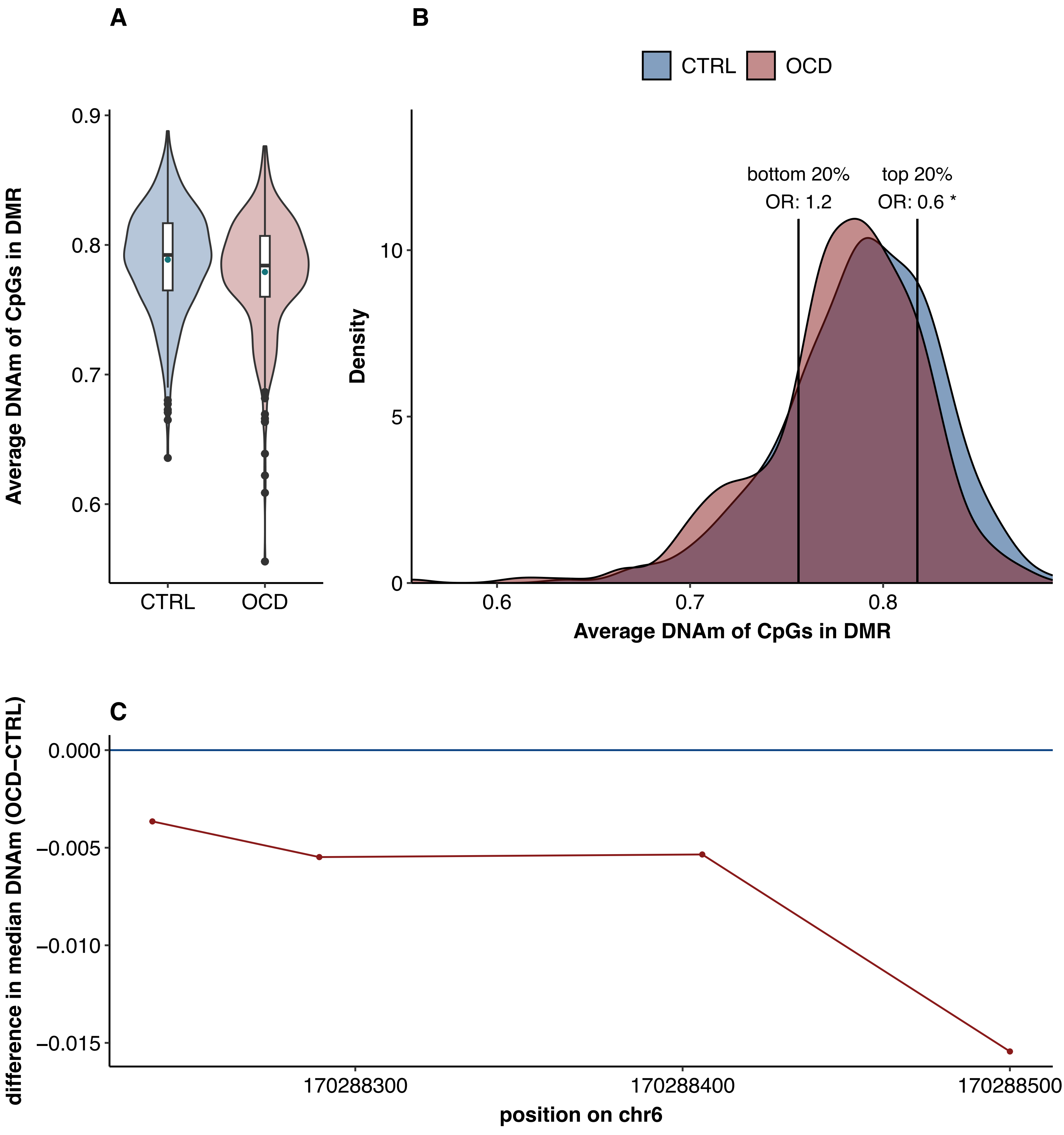

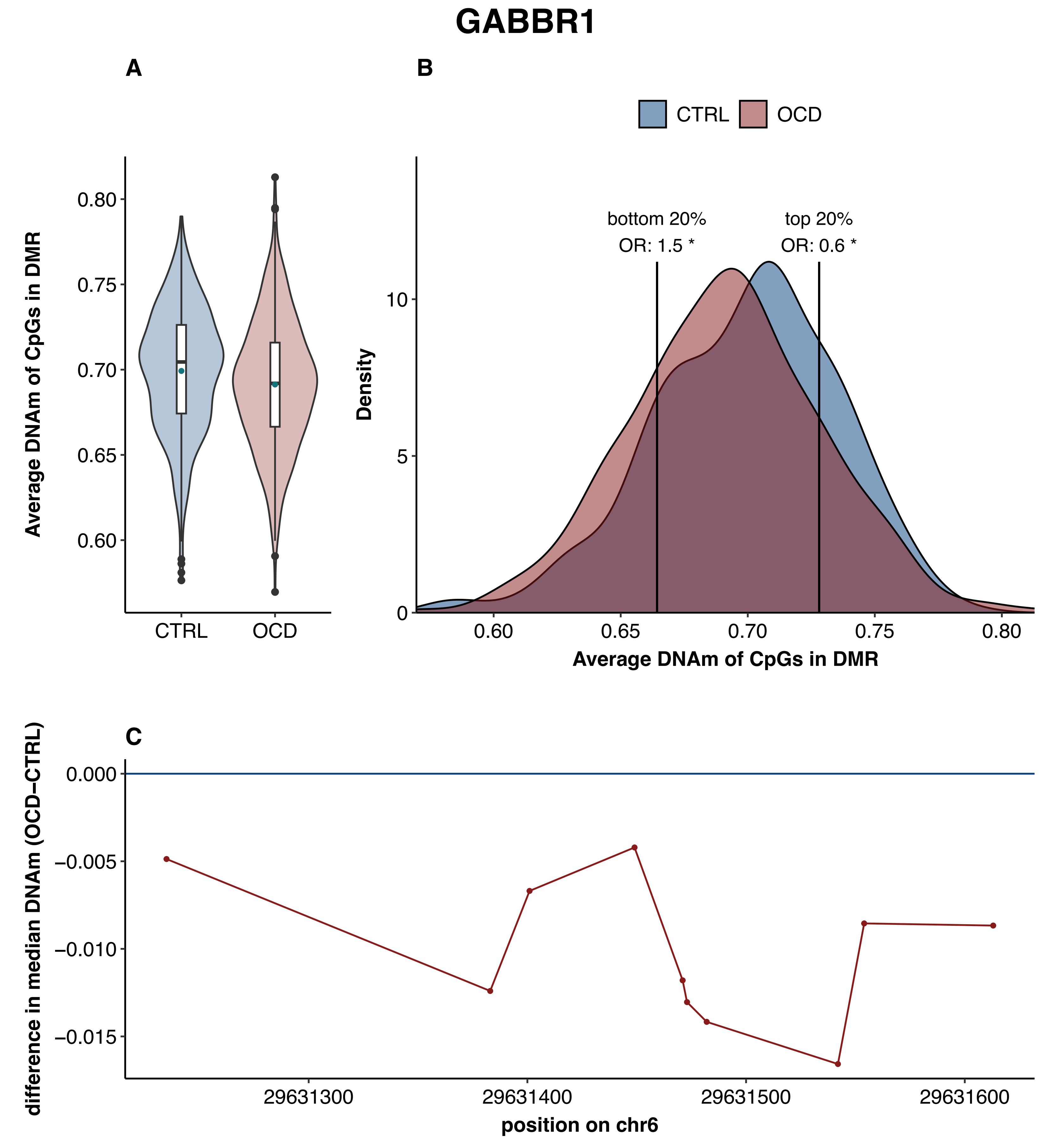
GABBR1
A
B
C
CTRL
OCD
bottom 20%
OR: 1.5 *
top 20%
OR: 0.6 *
Average DNAm of CpGs in DMR
Density
difference in median DNAm (OCD−CTRL)
position on chr6
0.80
0.75
0.70
0.65
0.60
10
5
0
0.000
−0.005
−0.010
−0.015
29631300
29631400
29631500
29631600

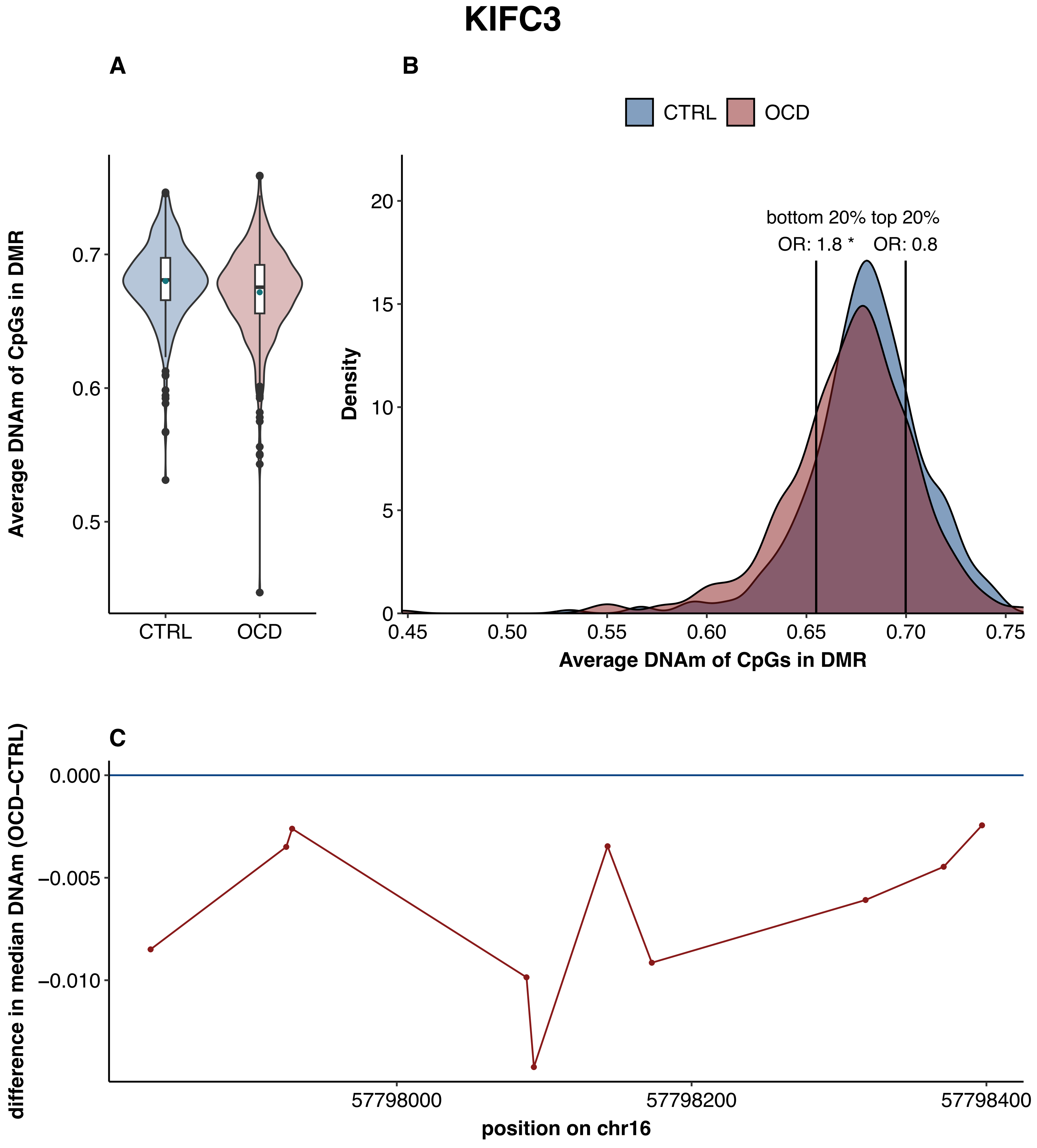

KIFC3
A
B
CTRL
OCD
Average DNAm of CpGs in DMR
0.7
0.6
0.5
CTRL
OCD
bottom 20% top 20%
OR: 1.8 *
OR: 0.8
Density
20
15
10
5
0
0.45
0.50
0.55
0.60
0.65
0.70
0.75
Average DNAm of CpGs in DMR
C
difference in median DNAm (OCD−CTRL)
0.000
−0.005
−0.010
57798000
57798200
57798400
position on chr16

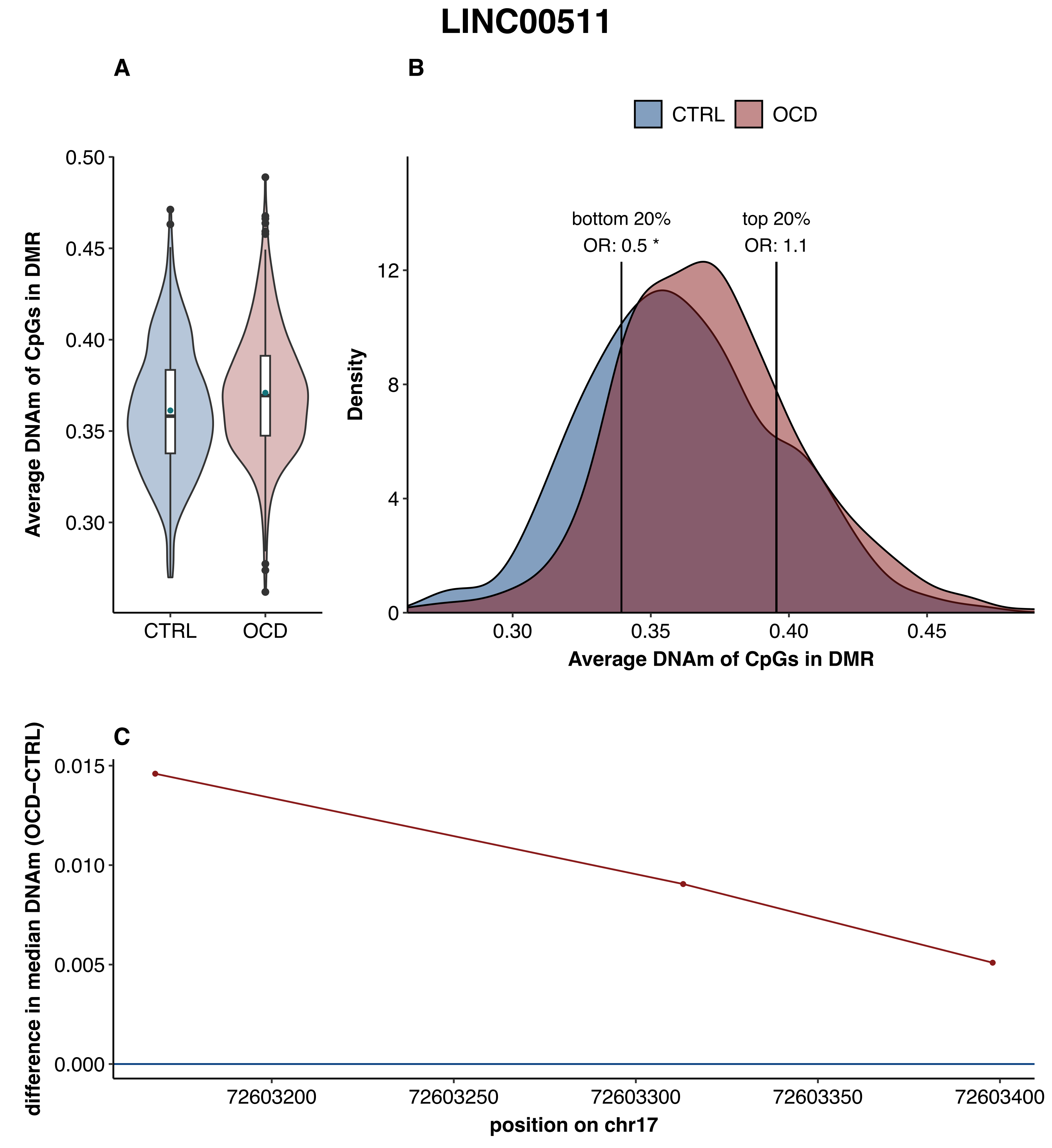

LINC00511
A
B
CTRL
OCD
Average DNAm of CpGs in DMR
0.50
0.45
0.40
0.35
0.30
CTRL
OCD
bottom 20%
OR: 0.5 *
top 20%
OR: 1.1
Density
12
8
4
0
0.30
0.35
0.40
0.45
Average DNAm of CpGs in DMR
C
difference in median DNAm (OCD−CTRL)
0.015
0.010
0.005
0.000
72603200
72603250
72603300
72603350
72603400
position on chr17

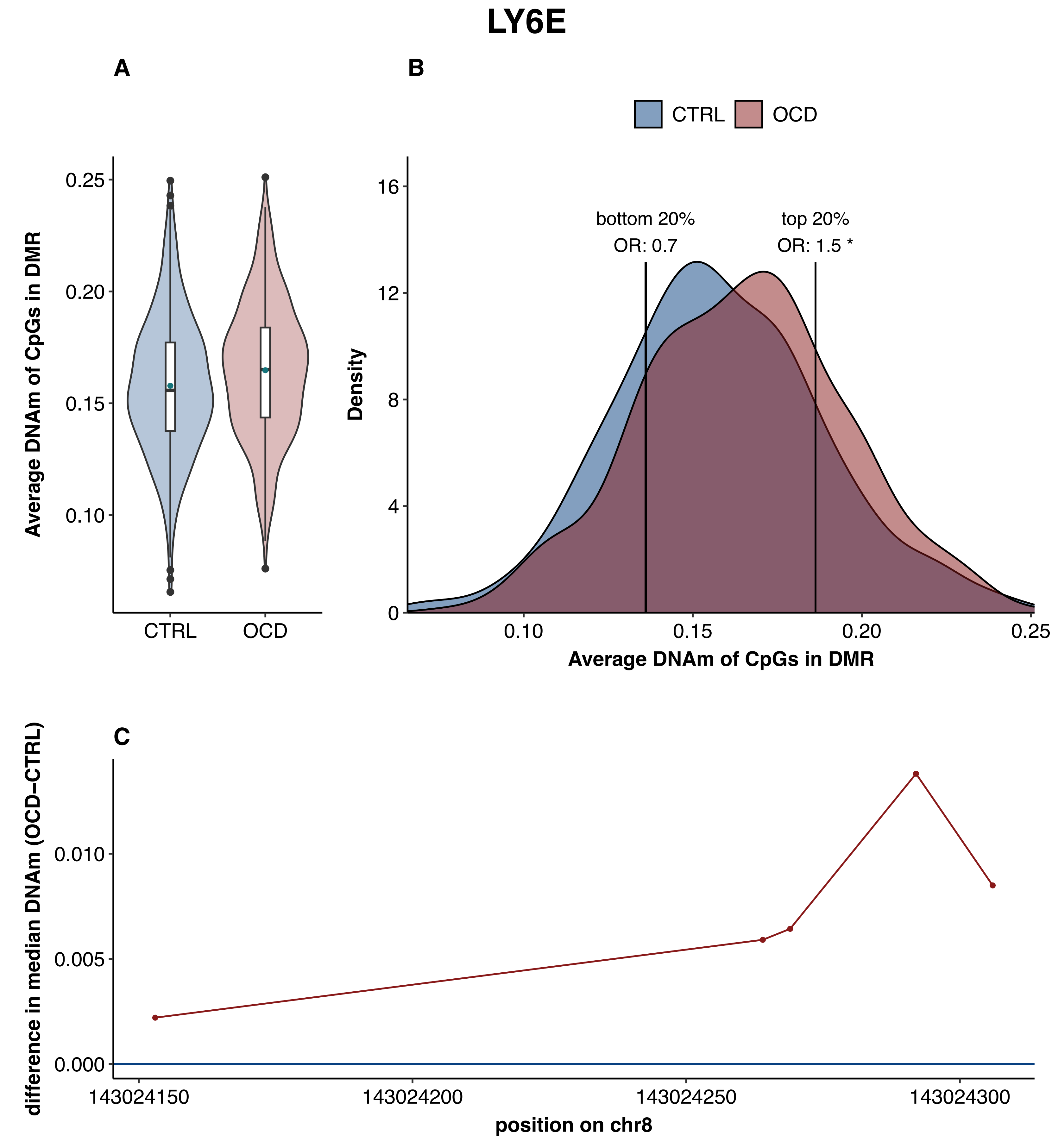
LY6E
A
B
CTRL
OCD
Average DNAm of CpGs in DMR
0.25
0.20
0.15
0.10
Density
16
12
8
4
0
bottom 20%
OR: 0.7
top 20%
OR: 1.5 *
0.10
0.15
0.20
0.25
Average DNAm of CpGs in DMR
C
difference in median DNAm (OCD−CTRL)
0.010
0.005
0.000
143024150
143024200
143024250
143024300
position on chr8

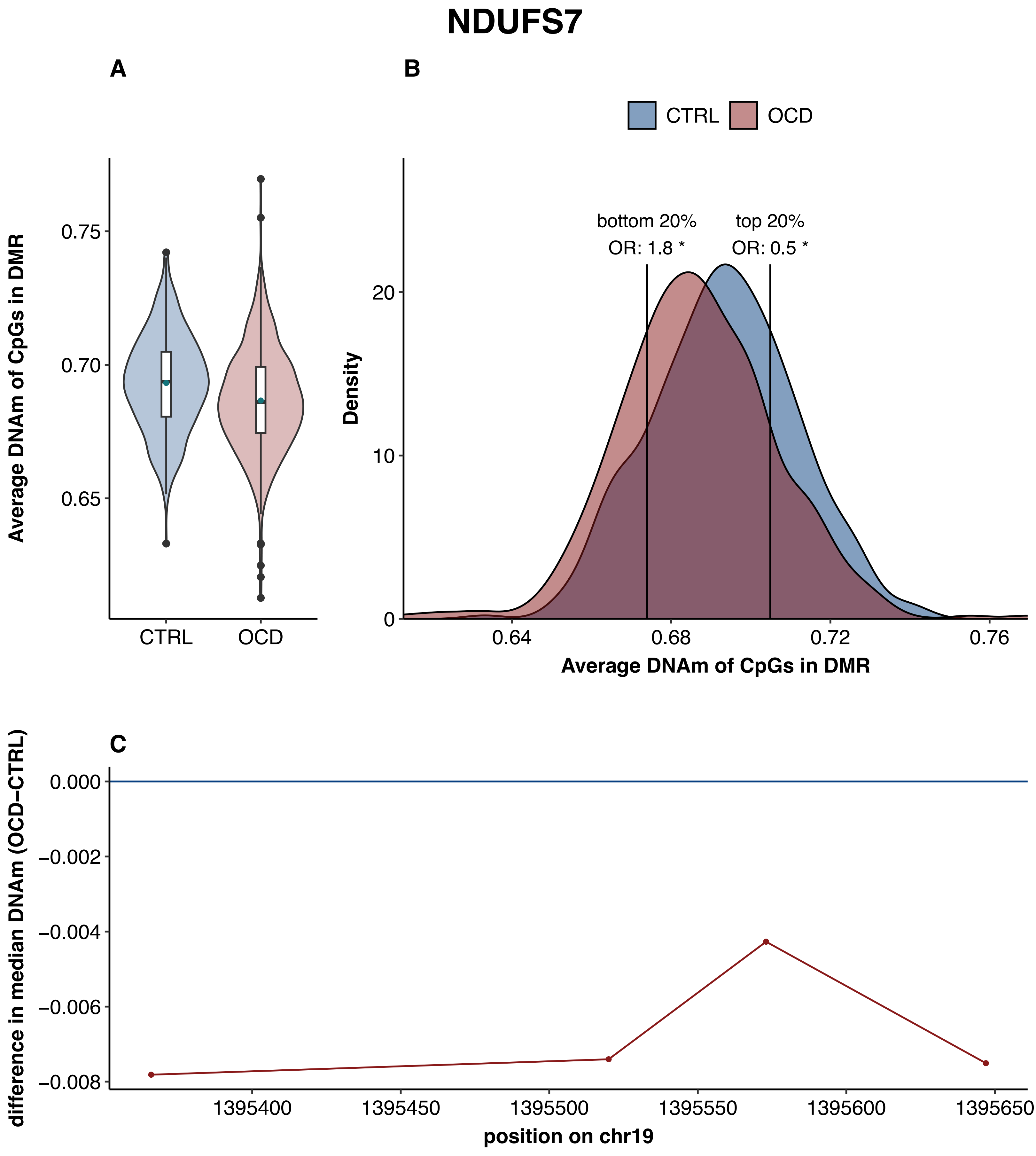
NDUFS7
A
B
C
CTRL
OCD
Average DNAm of CpGs in DMR
0.75
0.70
0.65
bottom 20%
OR: 1.8 *
top 20%
OR: 0.5 *
Density
20
10
0
0.64
0.68
0.72
0.76
difference in median DNAm (OCD−CTRL)
0.000
−0.002
−0.004
−0.006
−0.008
1395400
1395450
1395500
1395550
1395600
1395650
position on chr19

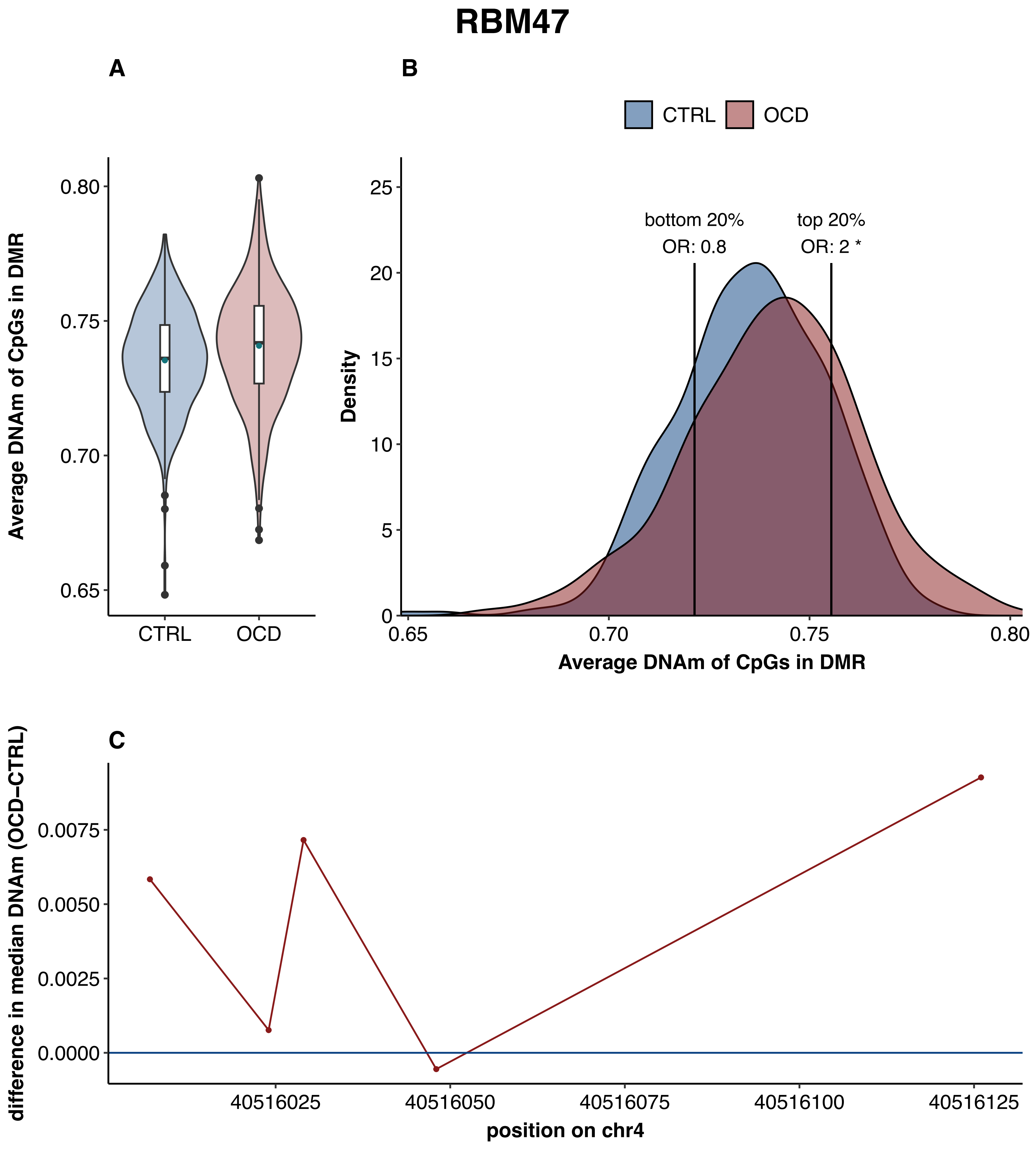
RBM47
A
B
CTRL
OCD
0.80
0.75
0.70
0.65
Average DNAm of CpGs in DMR
CTRL
OCD
25
20
15
10
5
0
Density
bottom 20%
OR: 0.8
top 20%
OR: 2 *
0.65
0.70
0.75
0.80
Average DNAm of CpGs in DMR
C
0.0075
0.0050
0.0025
0.0000
difference in median DNAm (OCD−CTRL)
40516025
40516050
40516075
40516100
40516125
position on chr4

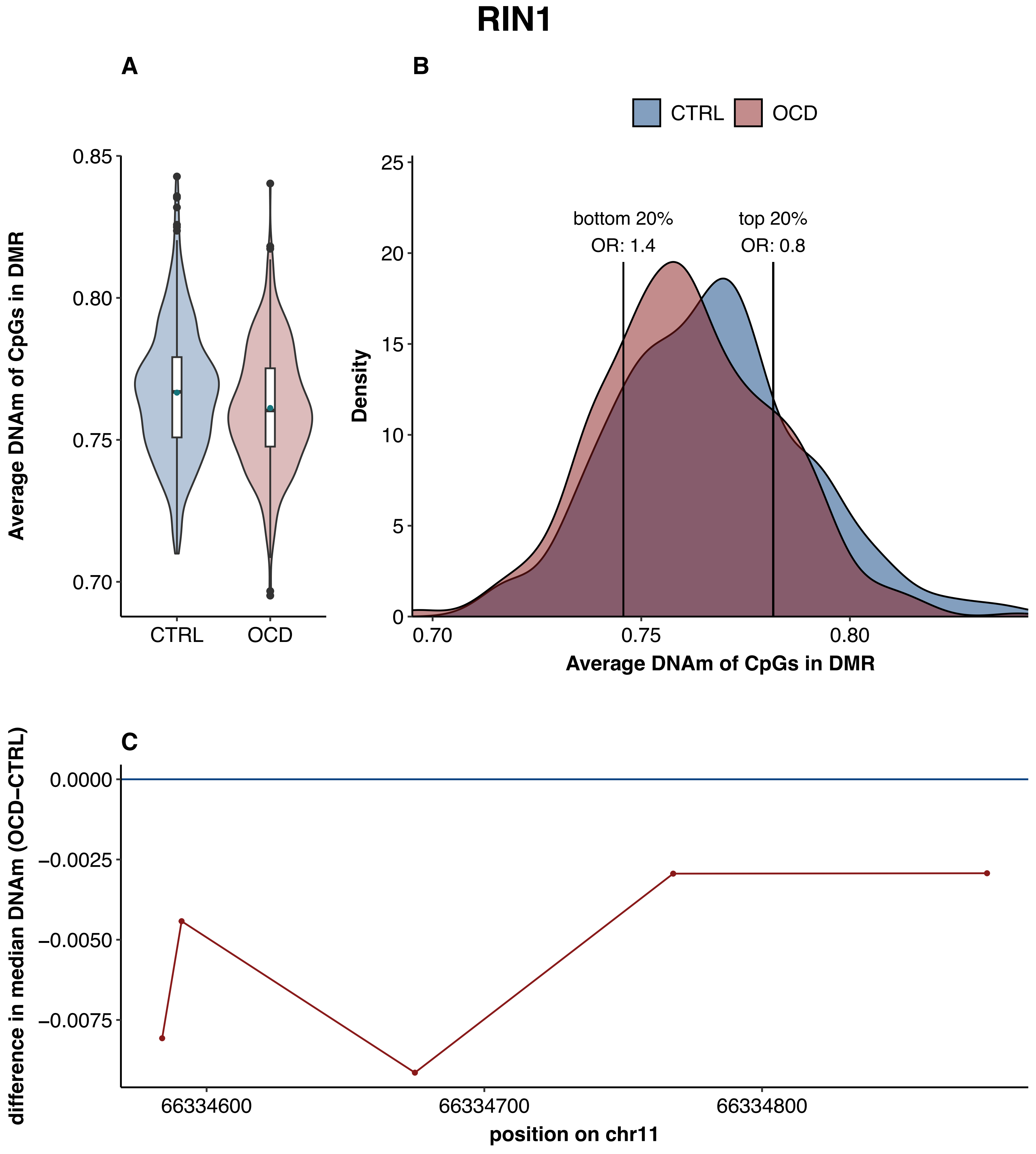
RIN1
A
B
CTRL
OCD
0.85
0.80
0.75
0.70
Average DNAm of CpGs in DMR
bottom 20%
OR: 1.4
top 20%
OR: 0.8
25
20
15
10
5
0
Density
0.70
0.75
0.80
Average DNAm of CpGs in DMR
C
0.0000
−0.0025
−0.0050
−0.0075
difference in median DNAm (OCD−CTRL)
66334600
66334700
66334800
position on chr11

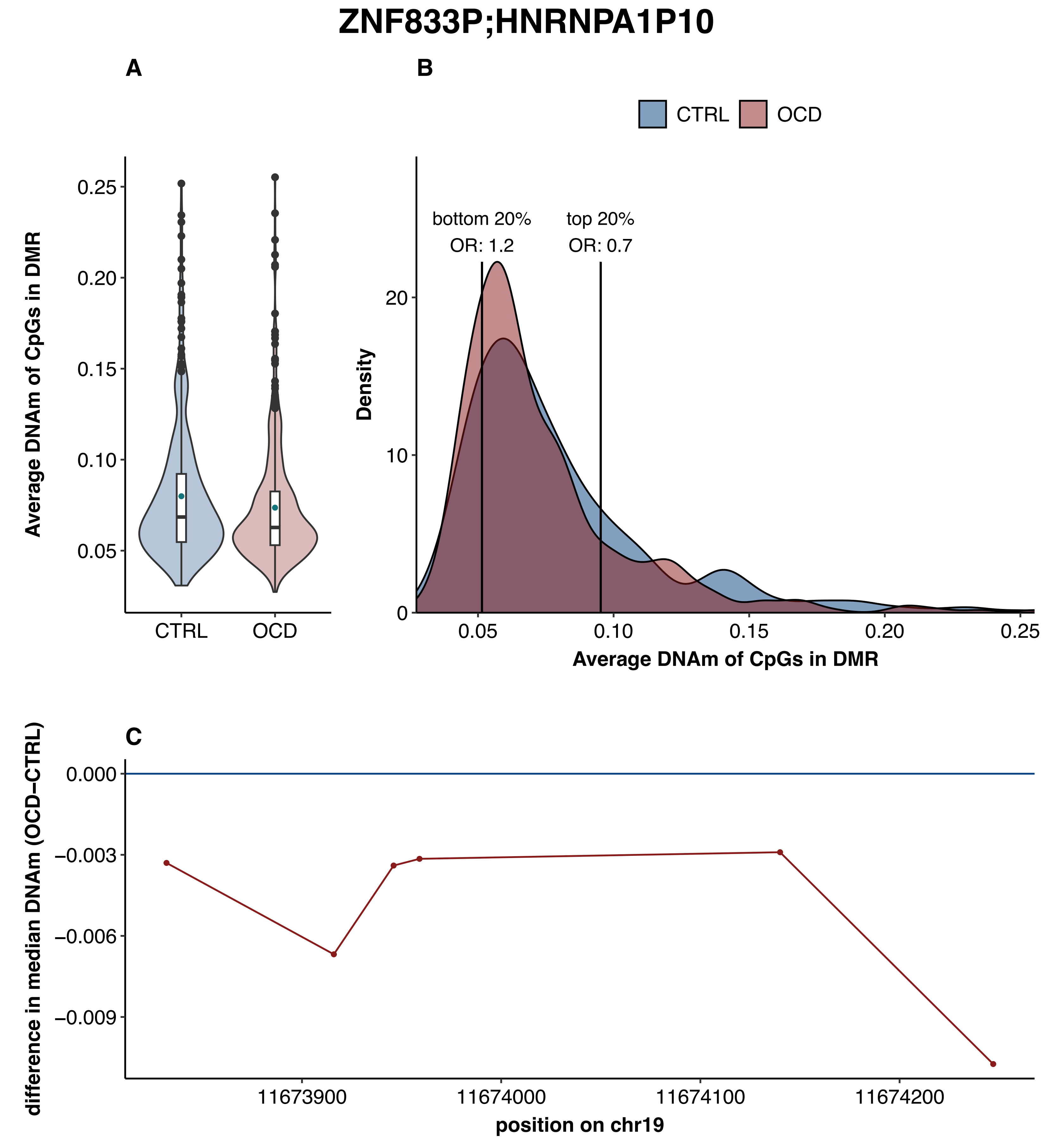
ZNF833P;HNRNPA1P10
A
B
CTRL
OCD
Average DNAm of CpGs in DMR
0.25
0.20
0.15
0.10
0.05
CTRL
OCD
bottom 20%
OR: 1.2
top 20%
OR: 0.7
Density
20
10
0
0.05
0.10
0.15
0.20
0.25
Average DNAm of CpGs in DMR
C
difference in median DNAm (OCD−CTRL)
0.000
−0.003
−0.006
−0.009
11673900
11674000
11674100
11674200
position on chr19

### sex-specific analysis in females

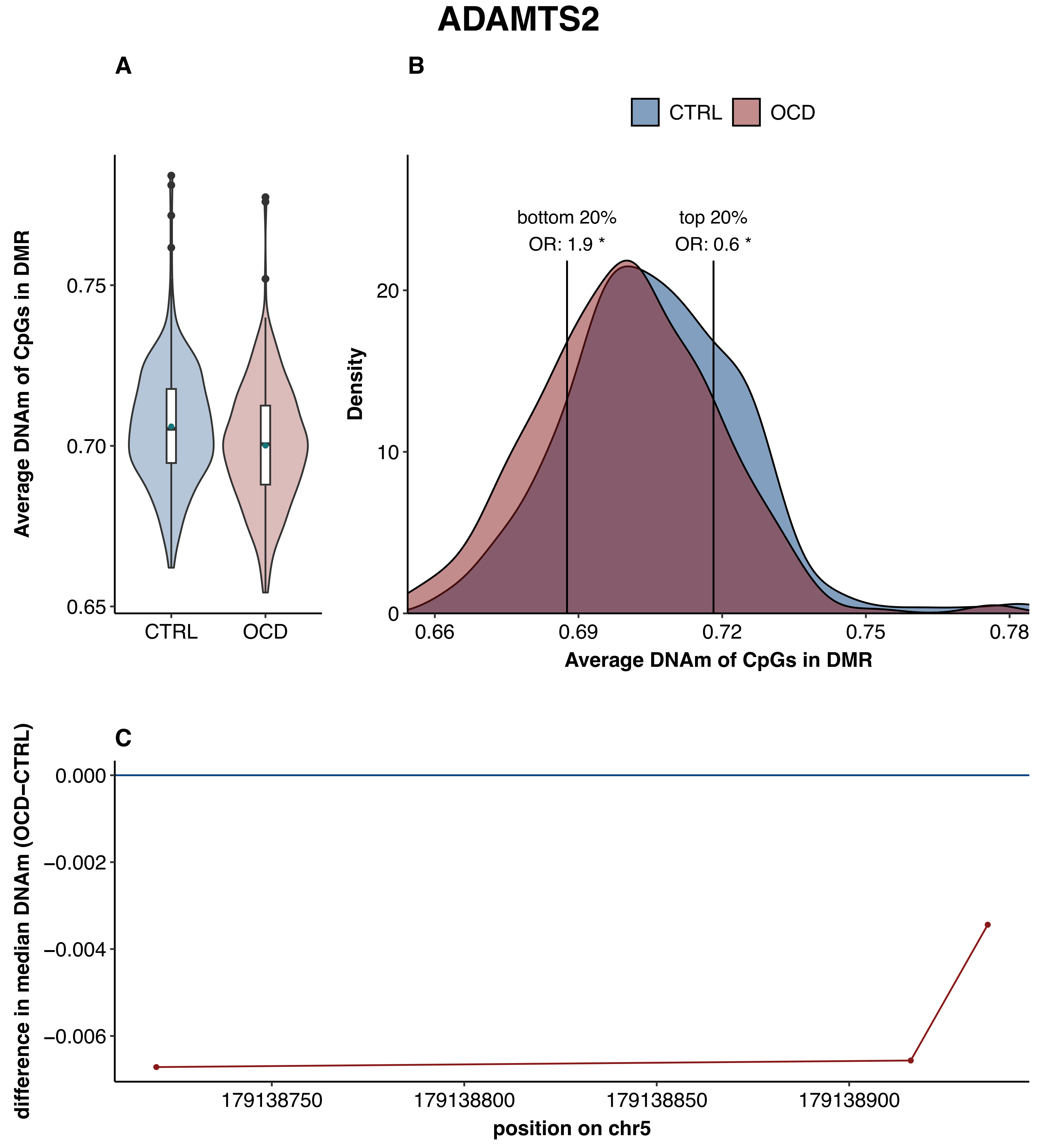

ADAMTS2
A
B
CTRL
OCD
Average DNAm of CpGs in DMR
0.75
0.70
0.65
CTRL
OCD
bottom 20%
OR: 1.9 *
top 20%
OR: 0.6 *
Density
20
10
0
0.66
0.69
0.72
0.75
0.78
Average DNAm of CpGs in DMR
C
difference in median DNAm (OCD−CTRL)
0.000
−0.002
−0.004
−0.006
179138750
179138800
179138850
179138900
position on chr5

### ARHGEF17

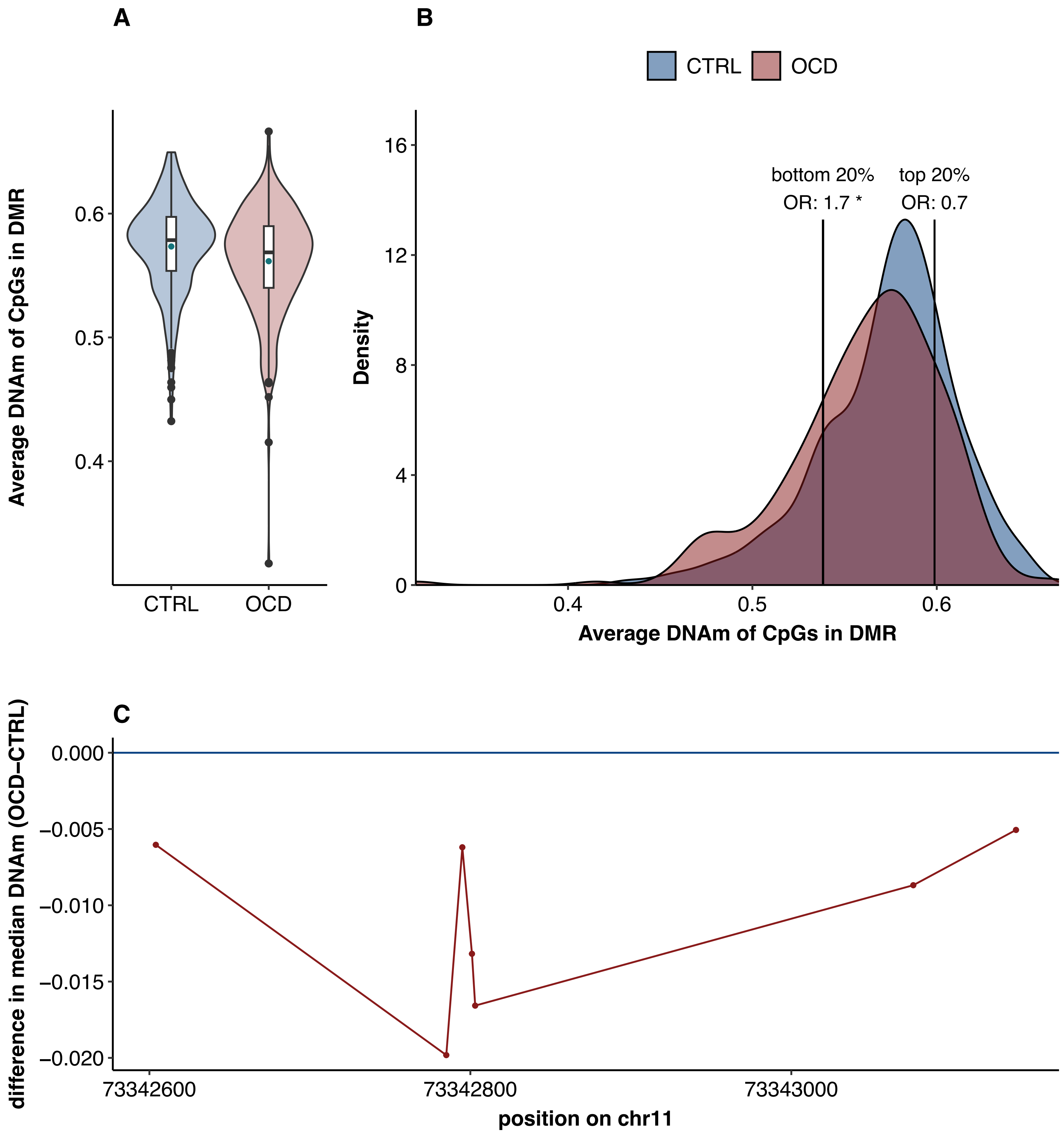

A
B
CTRL
OCD
Average DNAm of CpGs in DMR
0.6
0.5
0.4
Density
16
12
8
4
0
bottom 20%
OR: 1.7 *
top 20%
OR: 0.7
Average DNAm of CpGs in DMR
C
difference in median DNAm (OCD−CTRL)
0.000
−0.005
−0.010
−0.015
−0.020
73342600
73342800
73343000
position on chr11

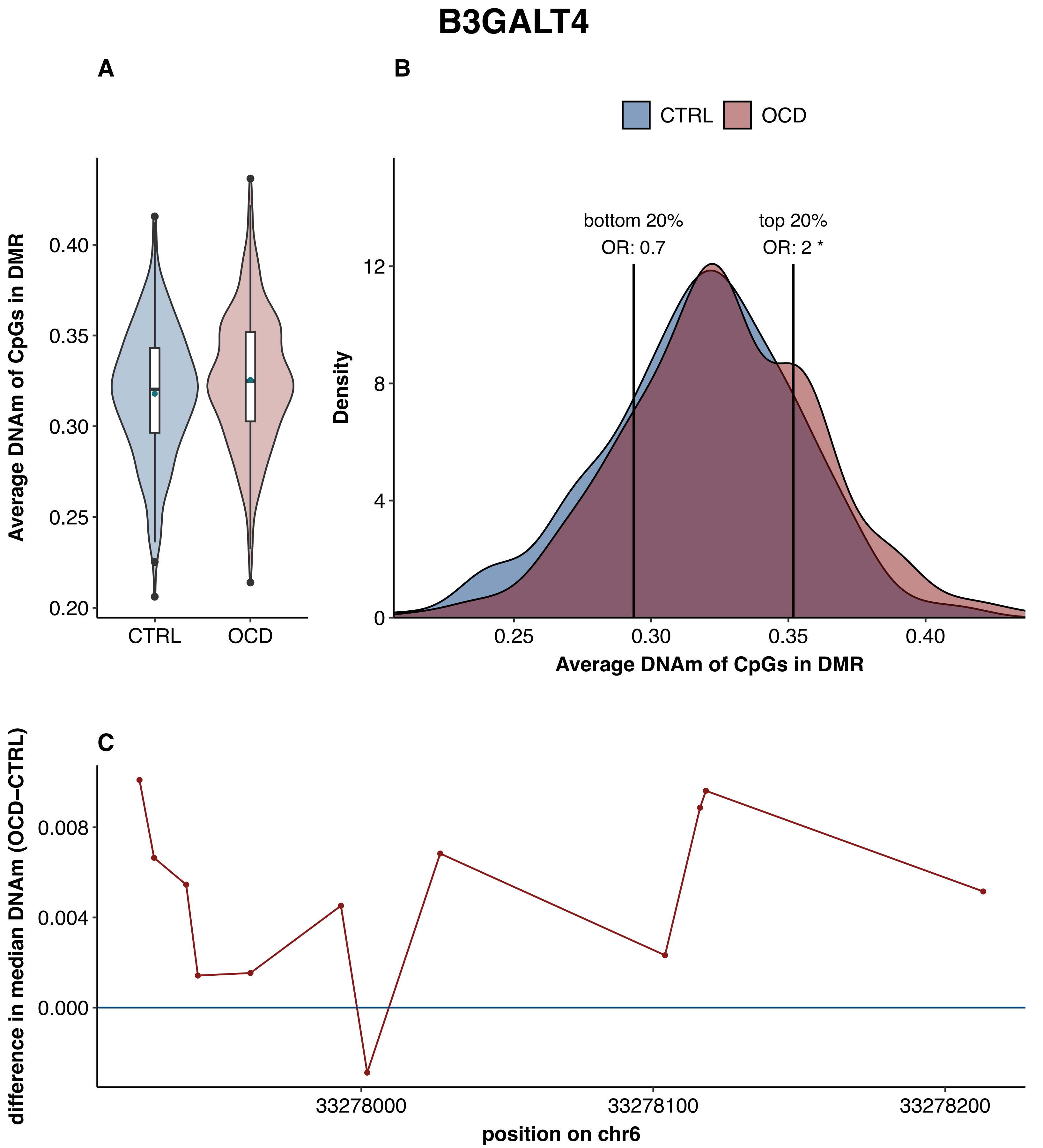

B3GALT4
A
B
CTRL
OCD
Average DNAm of CpGs in DMR
0.40
0.35
0.30
0.25
0.20
CTRL
OCD
bottom 20%
OR: 0.7
top 20%
OR: 2 *
Density
12
8
4
0
0.25
0.30
0.35
0.40
Average DNAm of CpGs in DMR
C
difference in median DNAm (OCD–CTRL)
0.008
0.004
0.000
33278000
33278100
33278200
position on chr6

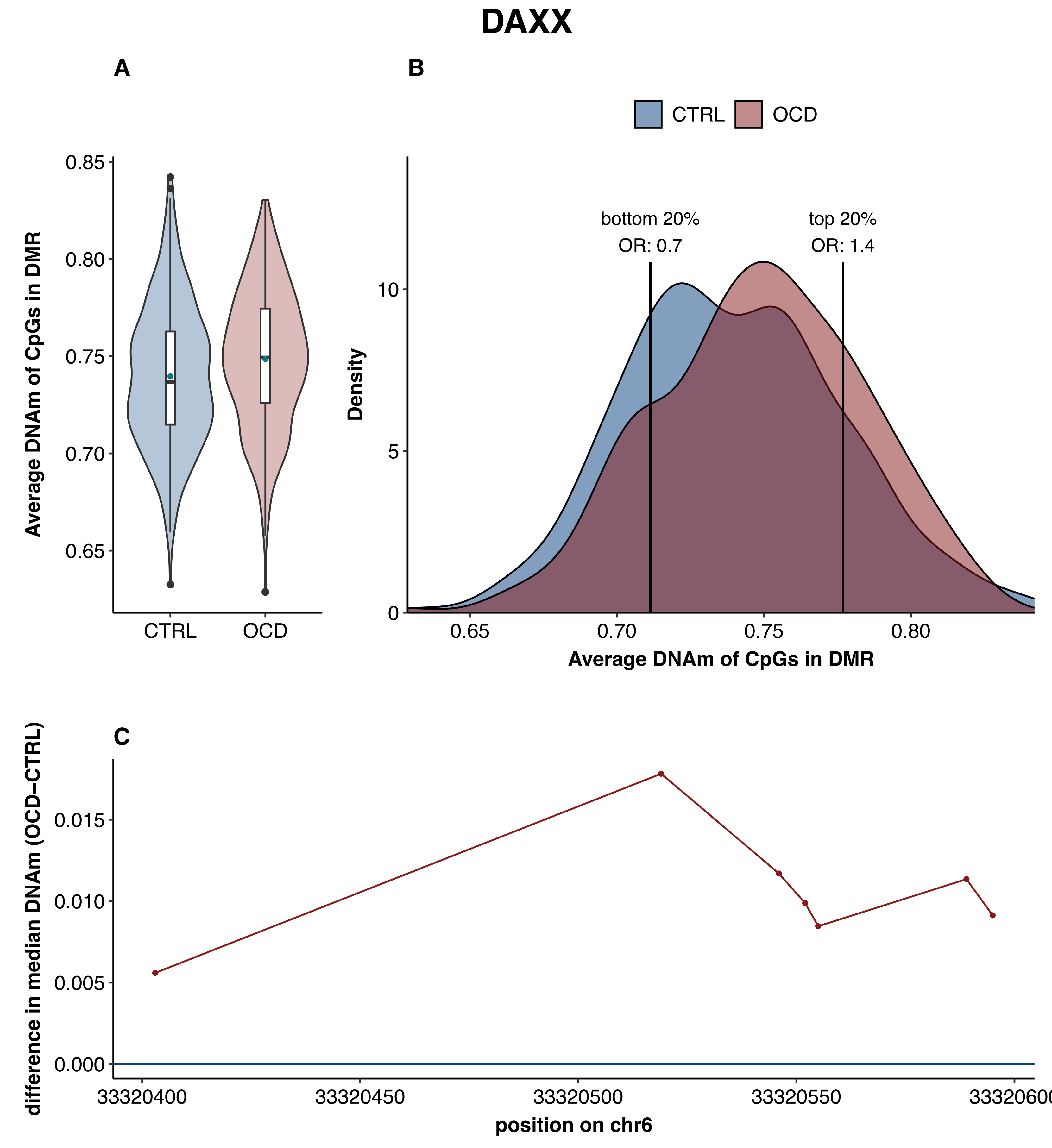
DAXX
A
B
C
CTRL
OCD
Average DNAm of CpGs in DMR
0.85
0.80
0.75
0.70
0.65
CTRL
OCD
bottom 20%
OR: 0.7
top 20%
OR: 1.4
Density
10
5
0
0.65
0.70
0.75
0.80
Average DNAm of CpGs in DMR
difference in median DNAm (OCD–CTRL)
0.015
0.010
0.005
0.000
33320400
33320450
33320500
33320550
33320600
position on chr6

### DYNLT4;BTBD19

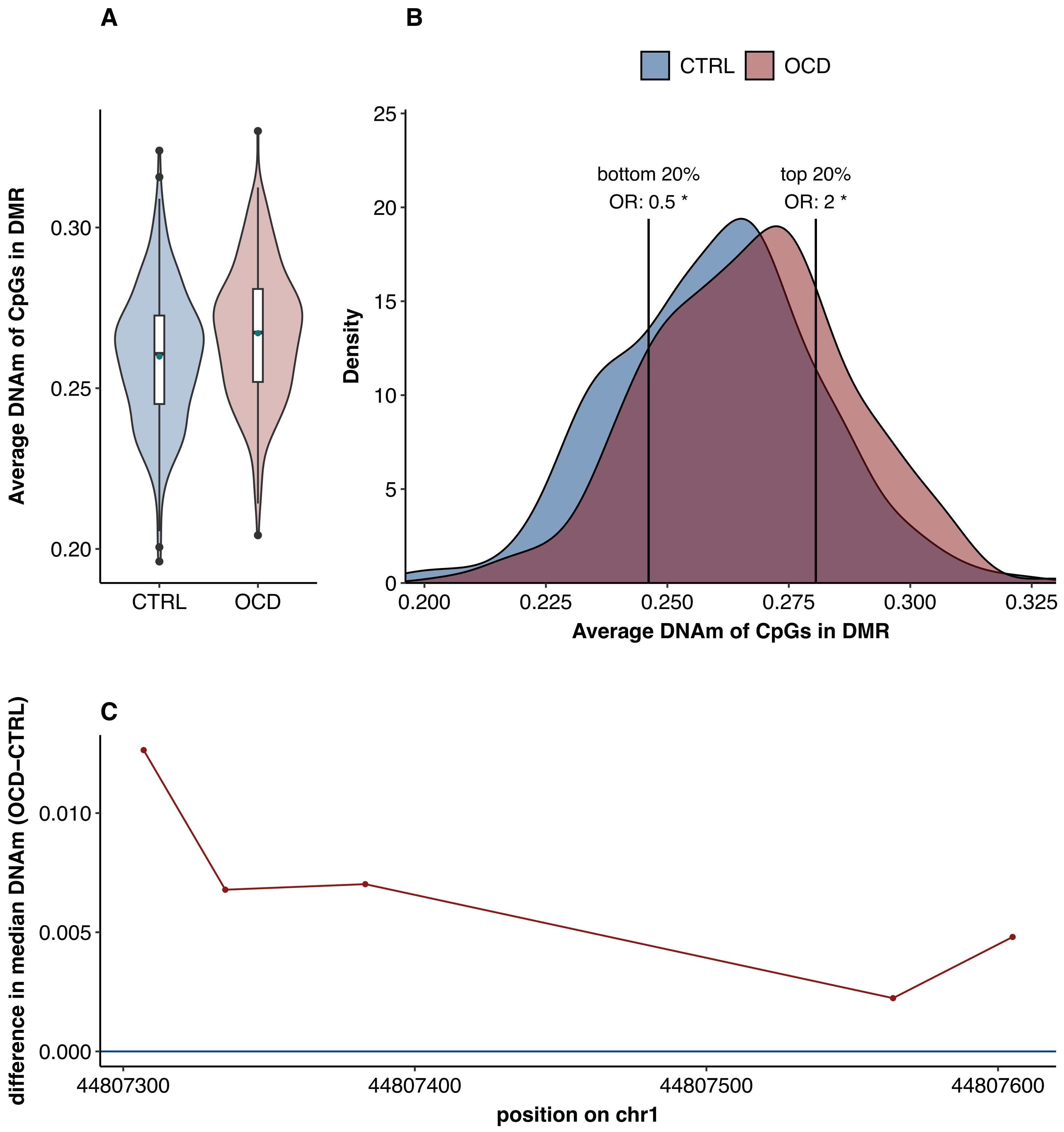

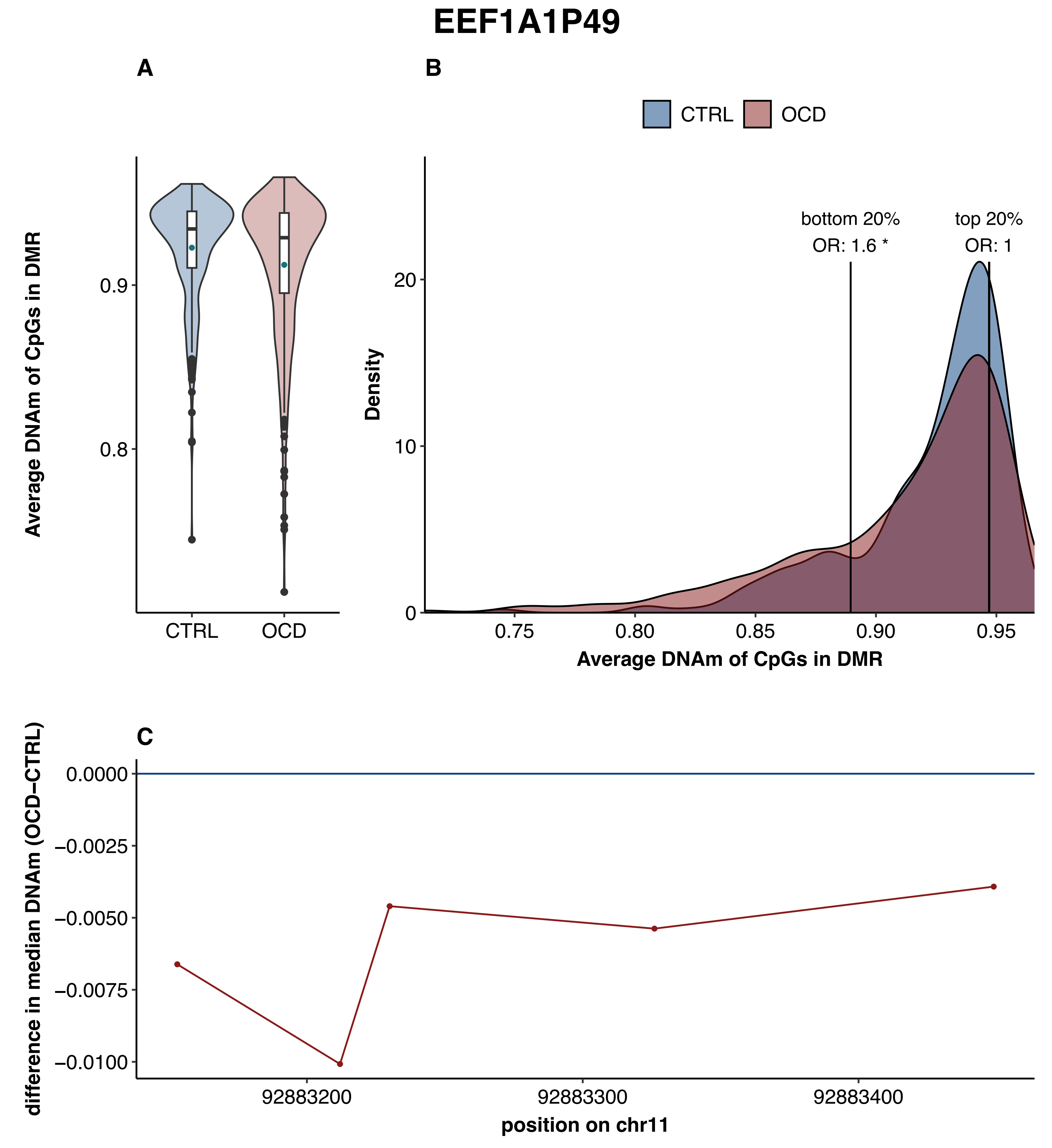
EEF1A1P49
A
B
CTRL
OCD
Average DNAm of CpGs in DMR
0.9
0.8
CTRL
OCD
bottom 20%
OR: 1.6 *
top 20%
OR: 1
Density
20
10
0
0.75
0.80
0.85
0.90
0.95
Average DNAm of CpGs in DMR
C
difference in median DNAm (OCD−CTRL)
0.0000
−0.0025
−0.0050
−0.0075
−0.0100
92883200
92883300
92883400
position on chr11

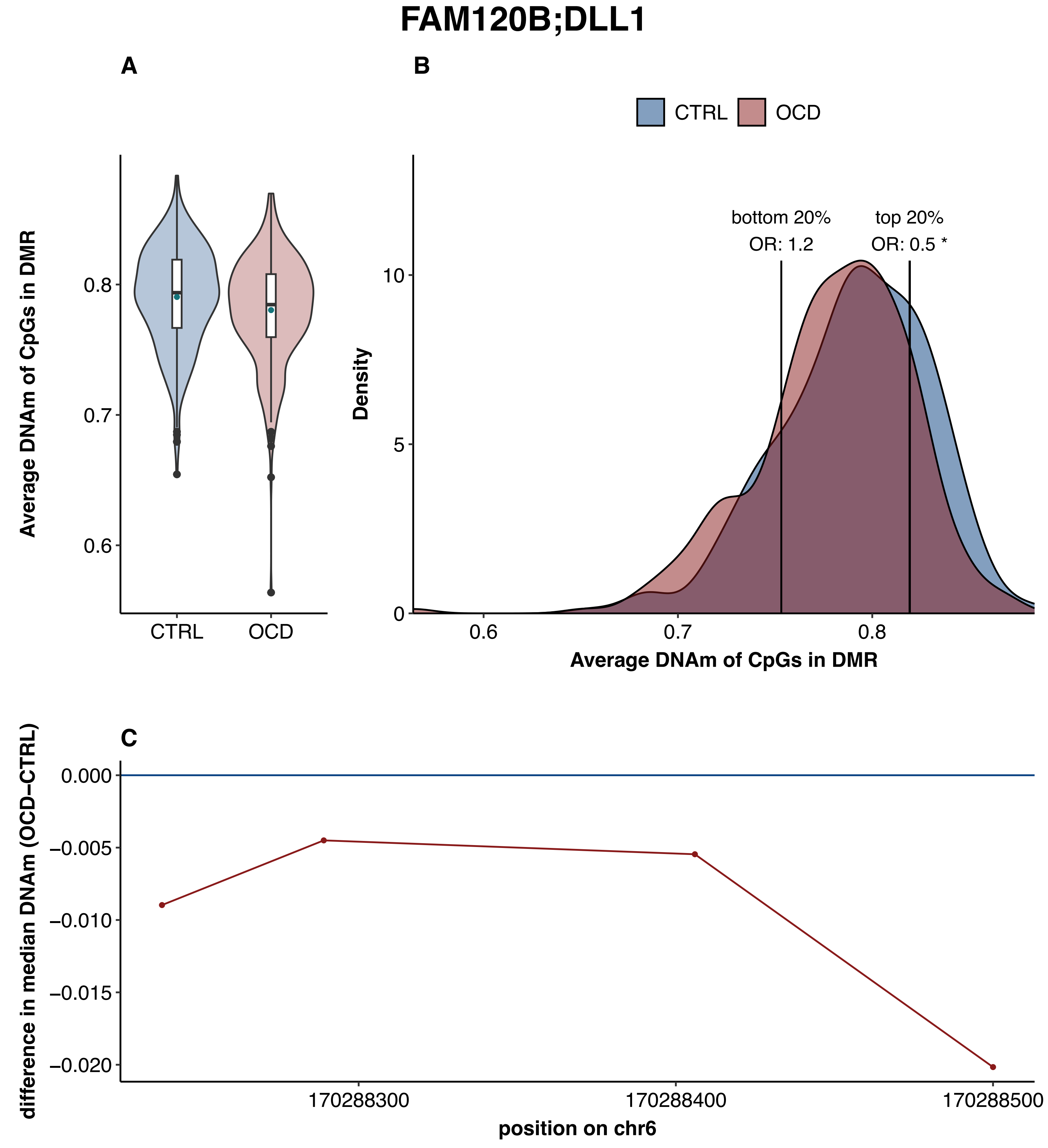
FAM120B;DLL1
A
B
C
CTRL
OCD
Average DNAm of CpGs in DMR
0.8
0.7
0.6
bottom 20%
OR: 1.2
top 20%
OR: 0.5 *
Density
10
5
0
0.6
0.7
0.8
difference in median DNAm (OCD−CTRL)
0.000
−0.005
−0.010
−0.015
−0.020
170288300
170288400
170288500
position on chr6

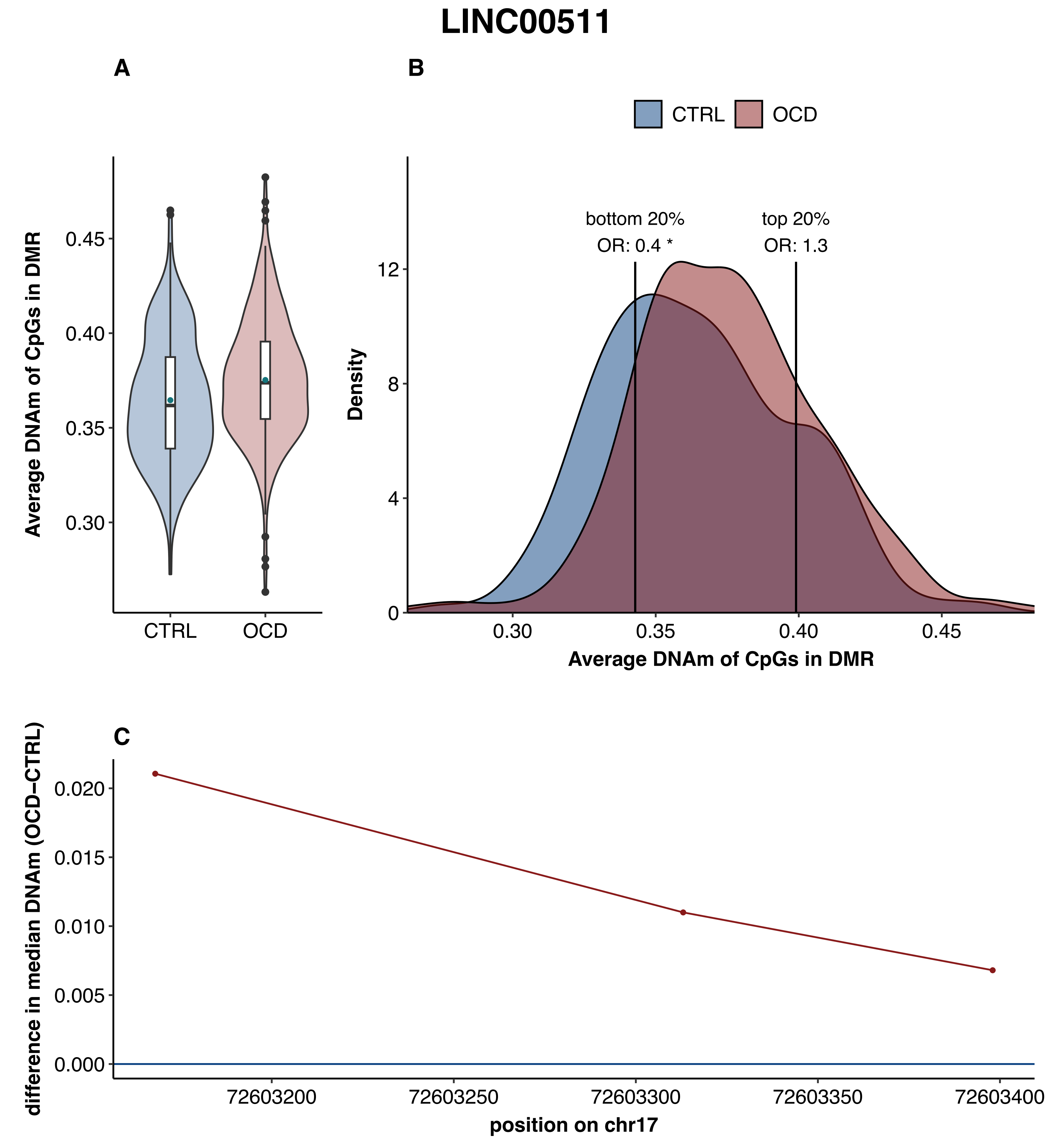

LINC00511
A
B
CTRL
OCD
Average DNAm of CpGs in DMR
0.45
0.40
0.35
0.30
CTRL
OCD
bottom 20%
OR: 0.4 *
top 20%
OR: 1.3
Density
12
8
4
0
0.30
0.35
0.40
0.45
Average DNAm of CpGs in DMR
C
difference in median DNAm (OCD−CTRL)
0.020
0.015
0.010
0.005
0.000
72603200
72603250
72603300
72603350
72603400
position on chr17

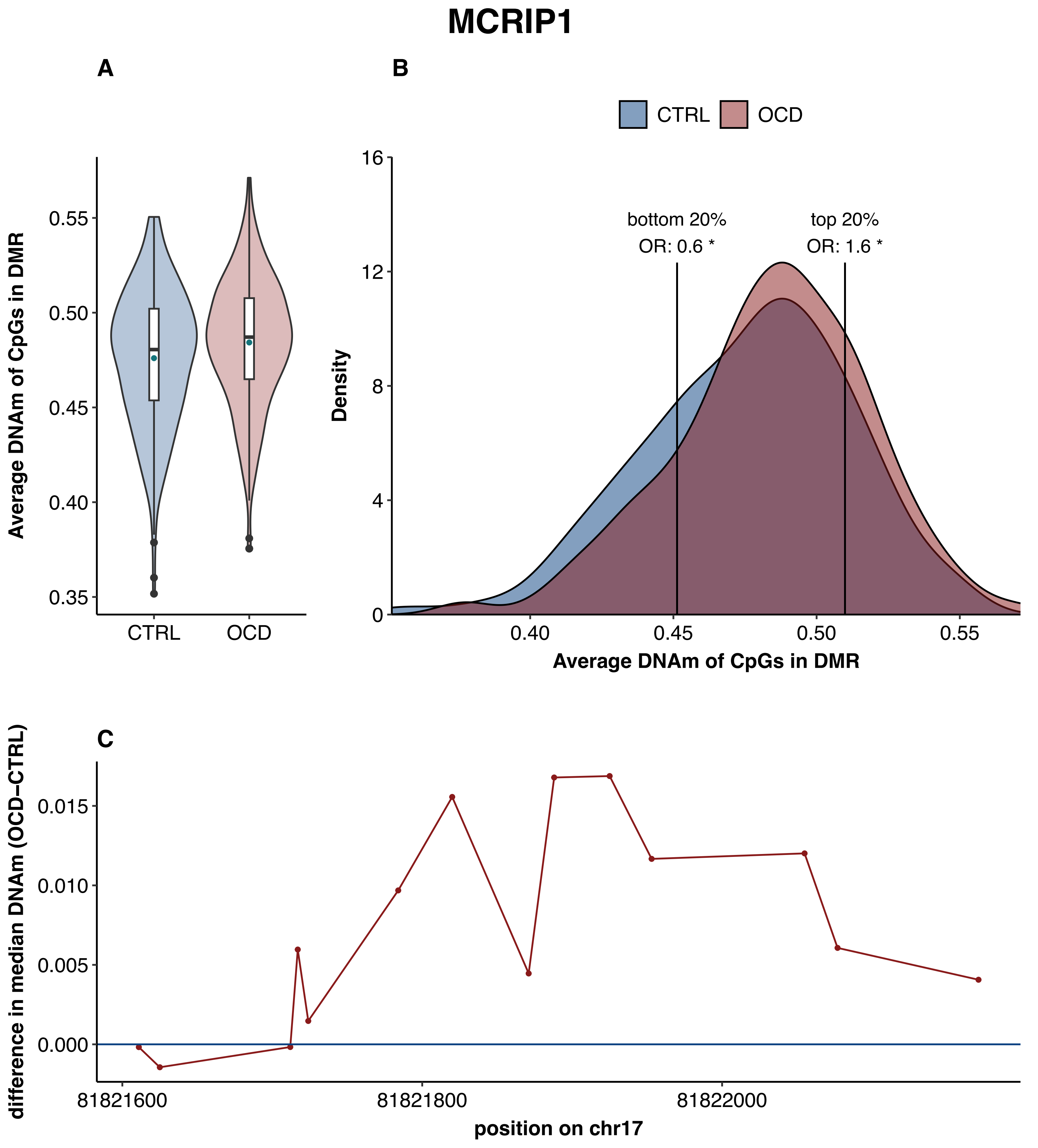

MCRIP1
A
B
C
CTRL
OCD
Average DNAm of CpGs in DMR
0.55
0.50
0.45
0.40
0.35
bottom 20%
OR: 0.6 *
top 20%
OR: 1.6 *
Density
16
12
8
4
0
difference in median DNAm (OCD−CTRL)
0.015
0.010
0.005
0.000
81821600
81821800
81822000
position on chr17

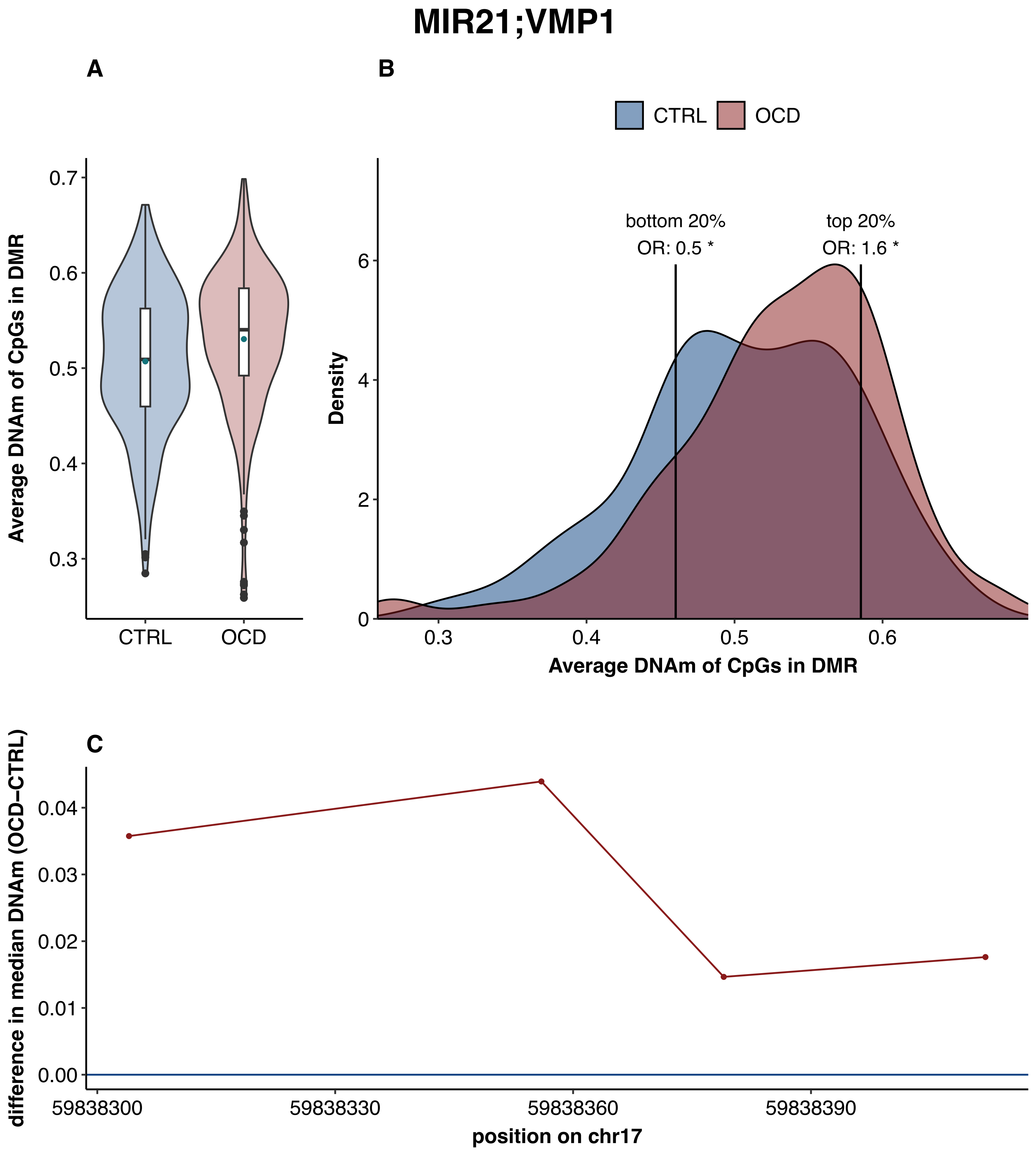

MIR21;VMP1
A
B
C
CTRL
OCD
Average DNAm of CpGs in DMR
0.7
0.6
0.5
0.4
0.3
bottom 20%
OR: 0.5 *
top 20%
OR: 1.6 *
Density
Average DNAm of CpGs in DMR
difference in median DNAm (OCD−CTRL)
0.04
0.03
0.02
0.01
0.00
59838300
59838330
59838360
59838390
position on chr17

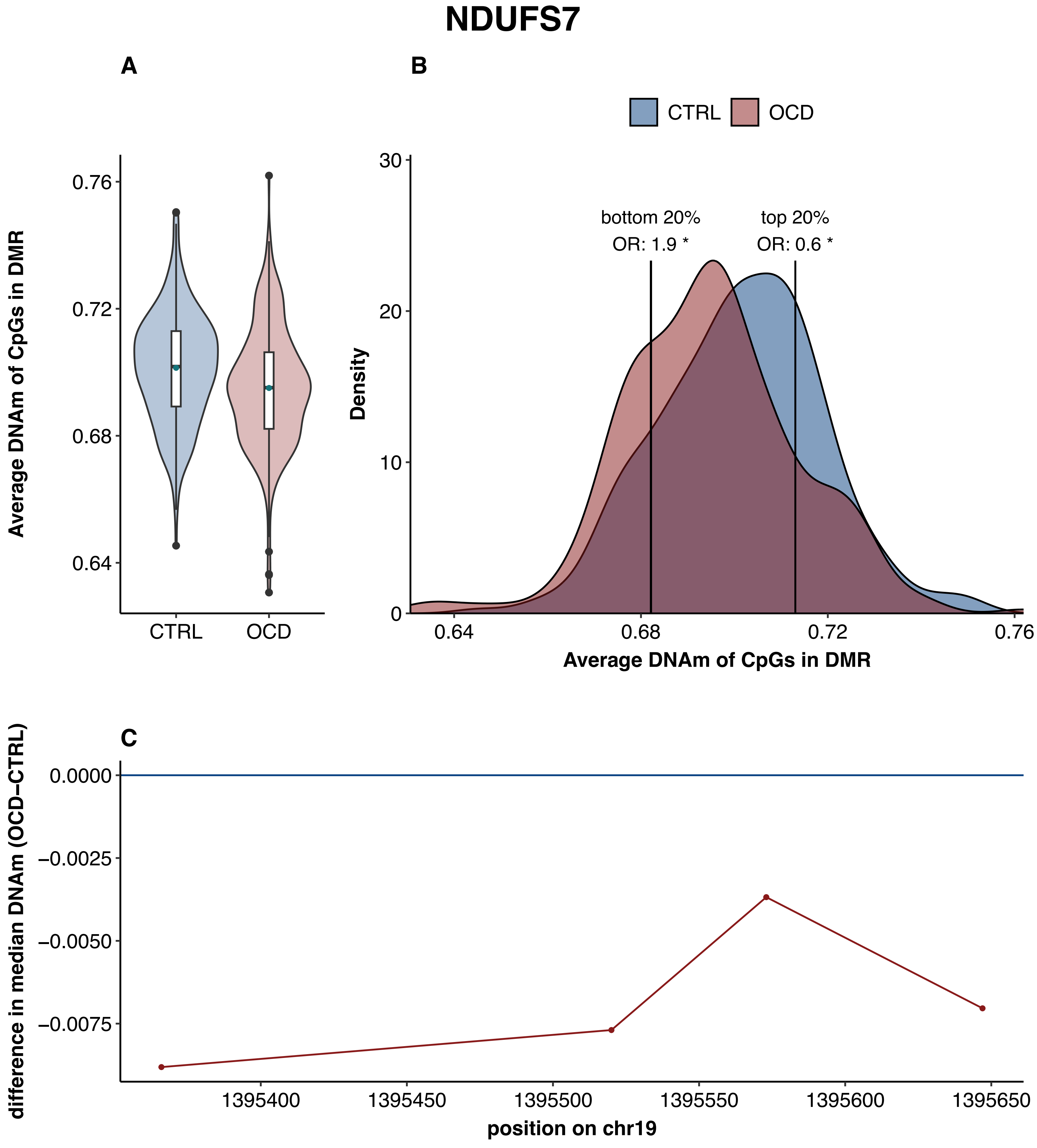
NDUFS7
A
B
C
CTRL
OCD
Average DNAm of CpGs in DMR
0.76
0.72
0.68
0.64
Density
30
20
10
0
bottom 20%
OR: 1.9 *
top 20%
OR: 0.6 *
0.64
0.68
0.72
0.76
difference in median DNAm (OCD−CTRL)
0.0000
−0.0025
−0.0050
−0.0075
1395400
1395450
1395500
1395550
1395600
1395650
position on chr19

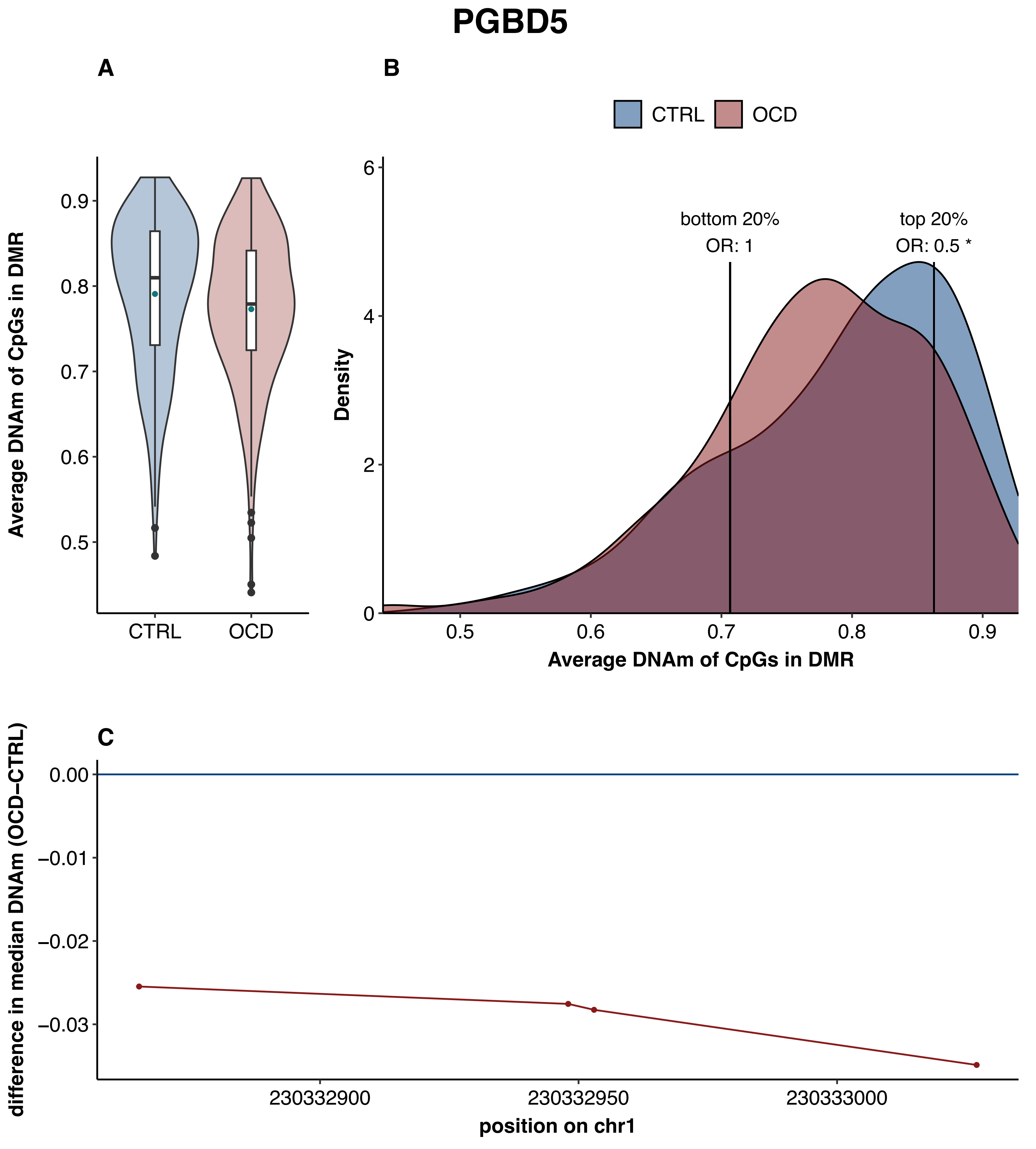
PGBD5
A
B
C
CTRL
OCD
Average DNAm of CpGs in DMR
0.9
0.8
0.7
0.6
0.5
Density
6
4
2
0
bottom 20%
OR: 1
top 20%
OR: 0.5 *
0.5
0.6
0.7
0.8
0.9
difference in median DNAm (OCD−CTRL)
0.00
−0.01
−0.02
−0.03
230332900
230332950
230333000
position on chr1

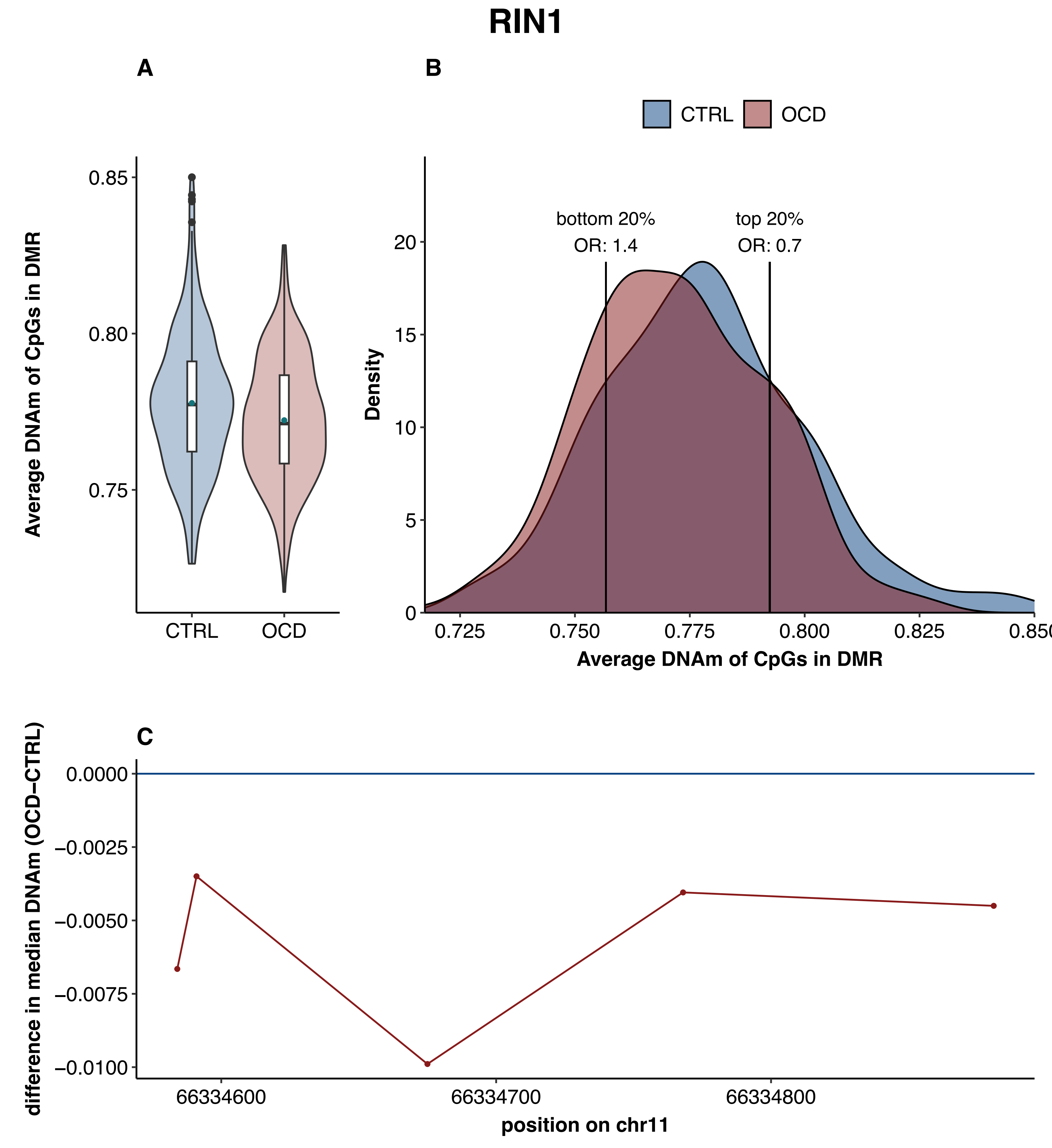
RIN1
A
B
C
CTRL
OCD
Average DNAm of CpGs in DMR
0.85
0.80
0.75
Density
bottom 20%
OR: 1.4
top 20%
OR: 0.7
20
15
10
5
0
0.725
0.750
0.775
0.800
0.825
0.850
difference in median DNAm (OCD−CTRL)
0.0000
−0.0025
−0.0050
−0.0075
−0.0100
66334600
66334700
66334800
position on chr11

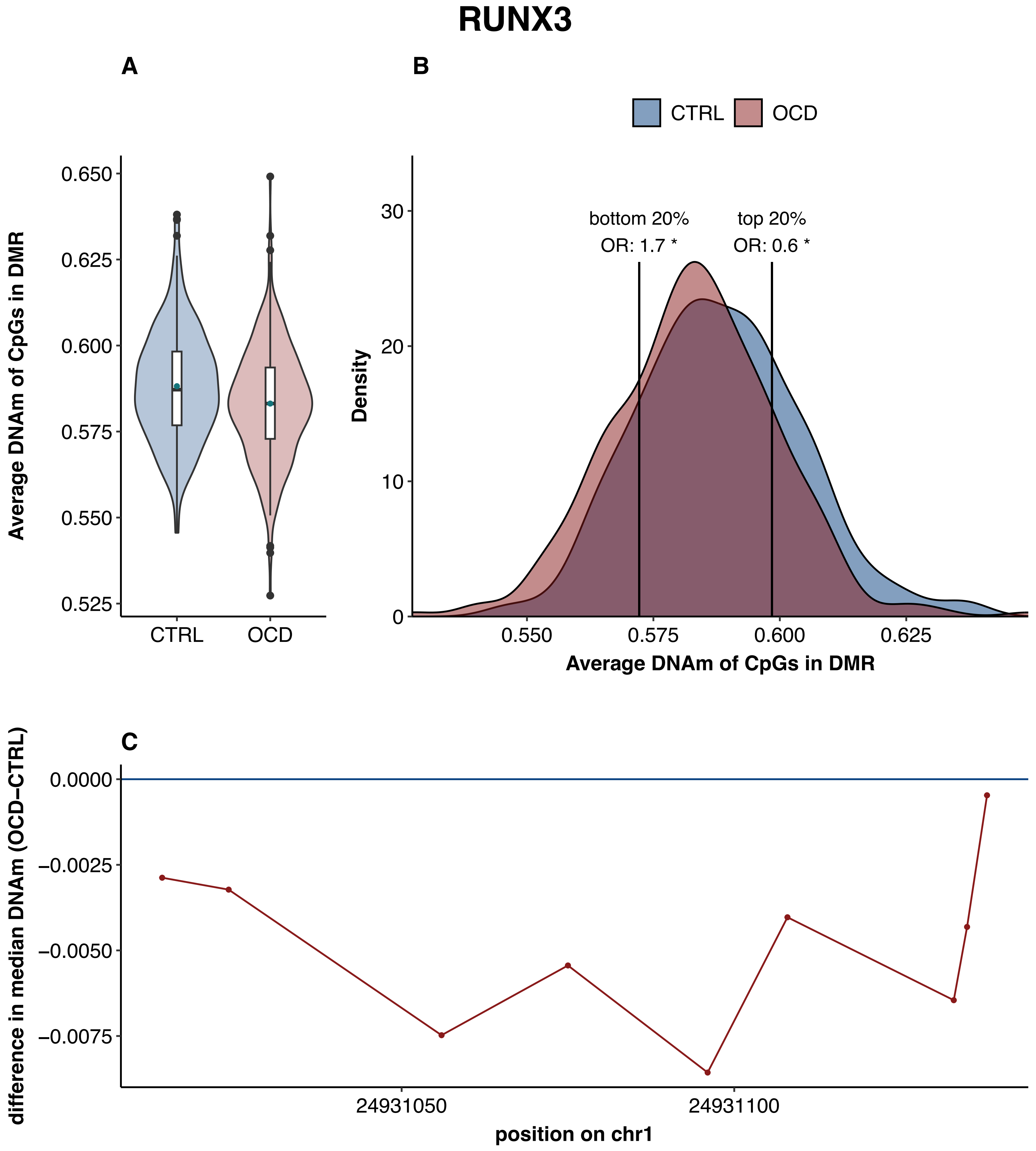
RUNX3
A
B
CTRL
OCD
Average DNAm of CpGs in DMR
0.650
0.625
0.600
0.575
0.550
0.525
bottom 20%
OR: 1.7 *
top 20%
OR: 0.6 *
Density
30
20
10
0
0.550
0.575
0.600
0.625
C
difference in median DNAm (OCD−CTRL)
0.0000
−0.0025
−0.0050
−0.0075
24931050
24931100
position on chr1

### sex-specific analysis in males

### AKAP12

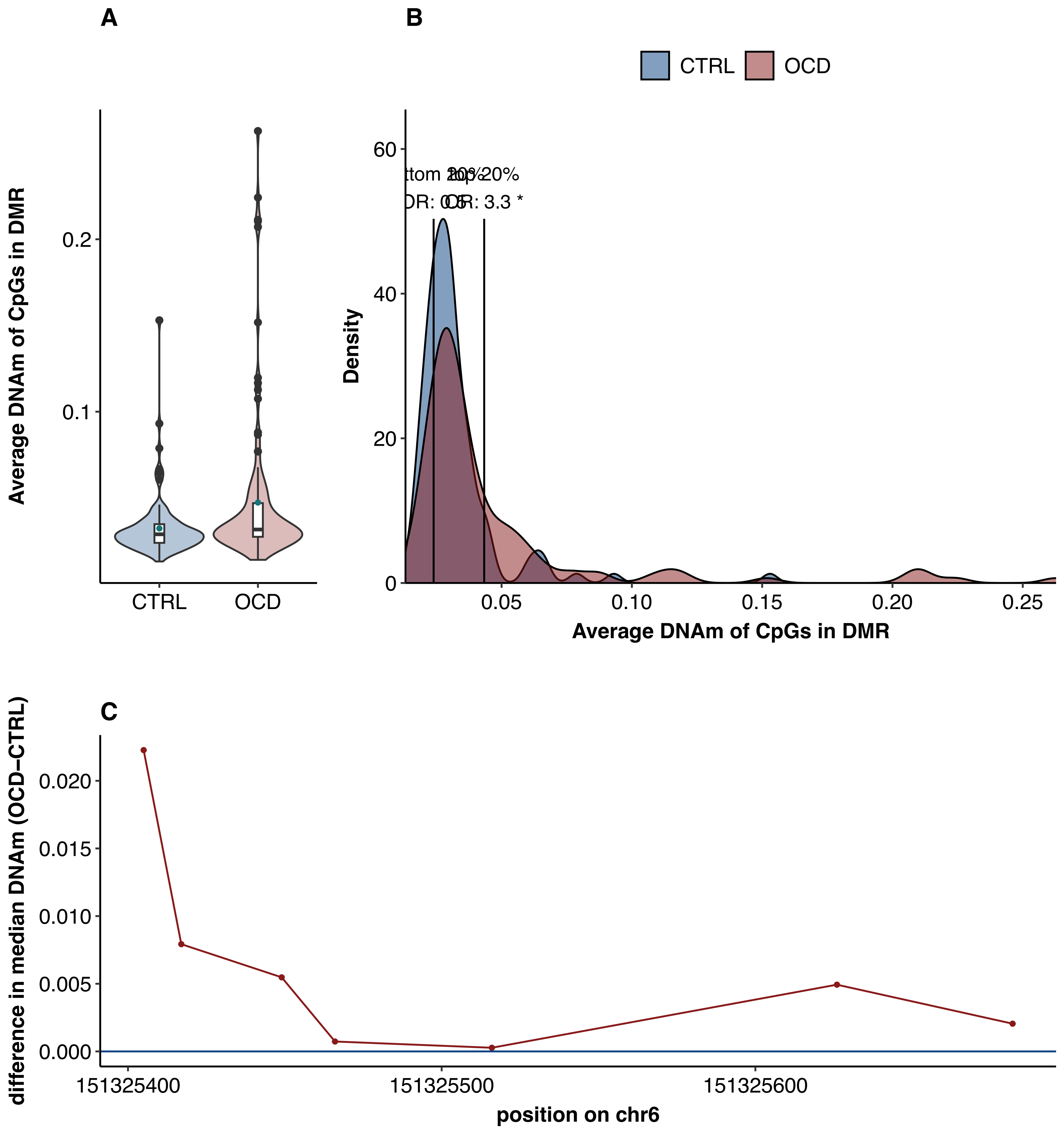

A
B
CTRL
OCD
Average DNAm of CpGs in DMR
0.2
0.1
CTRL
OCD
Density
60
40
20
0
ttom 20%
top 20%
OR: 0.5
OR: 3.3 *
0.05
0.10
0.15
0.20
0.25
Average DNAm of CpGs in DMR
C
difference in median DNAm (OCD−CTRL)
0.020
0.015
0.010
0.005
0.000
151325400
151325500
151325600
position on chr6

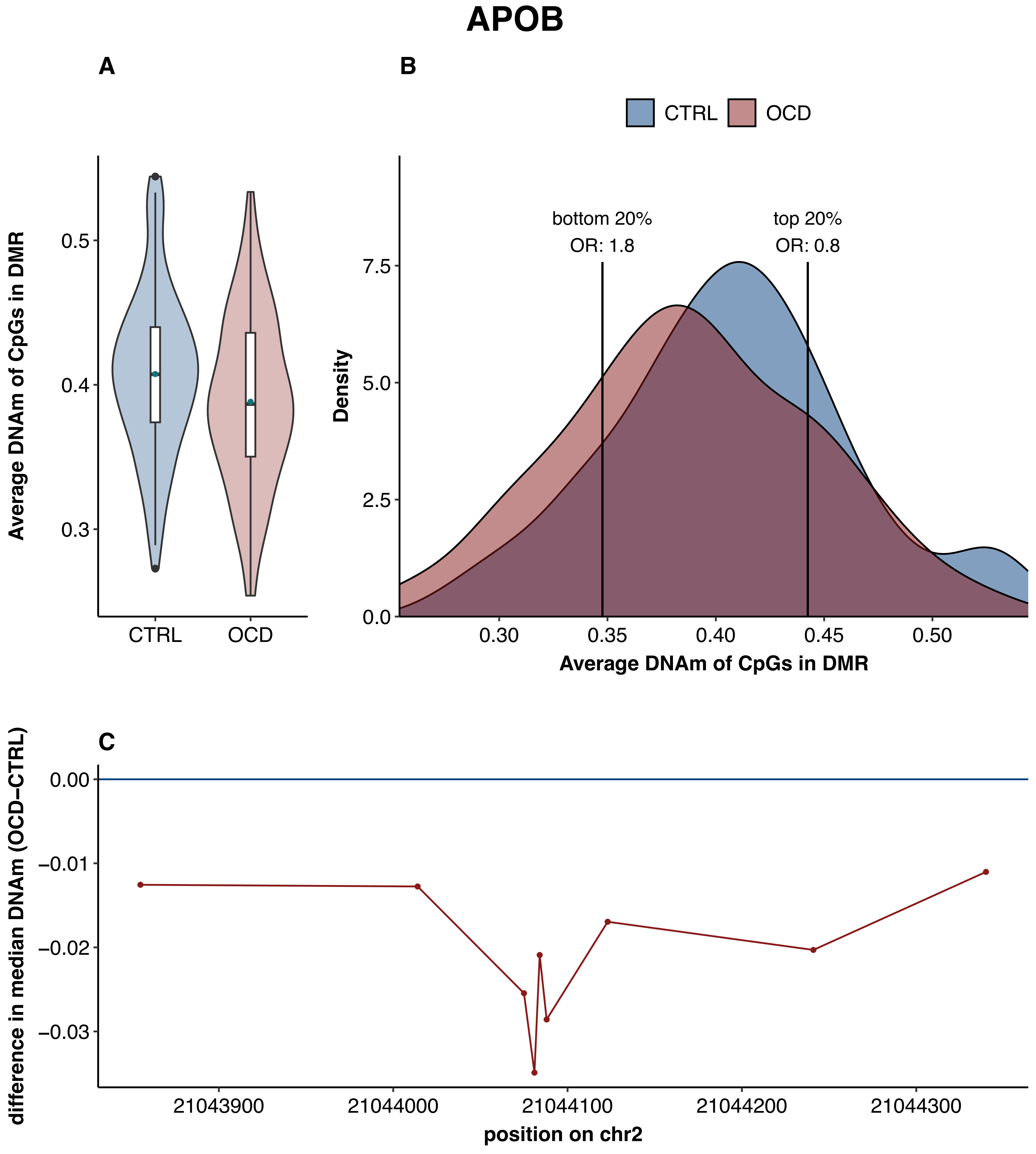
APOB
A
B
C
CTRL
OCD
Average DNAm of CpGs in DMR
0.5
0.4
0.3
bottom 20%
OR: 1.8
top 20%
OR: 0.8
Density
7.5
5.0
2.5
0.0
0.30
0.35
0.40
0.45
0.50
difference in median DNAm (OCD−CTRL)
0.00
−0.01
−0.02
−0.03
21043900
21044000
21044100
21044200
21044300
position on chr2

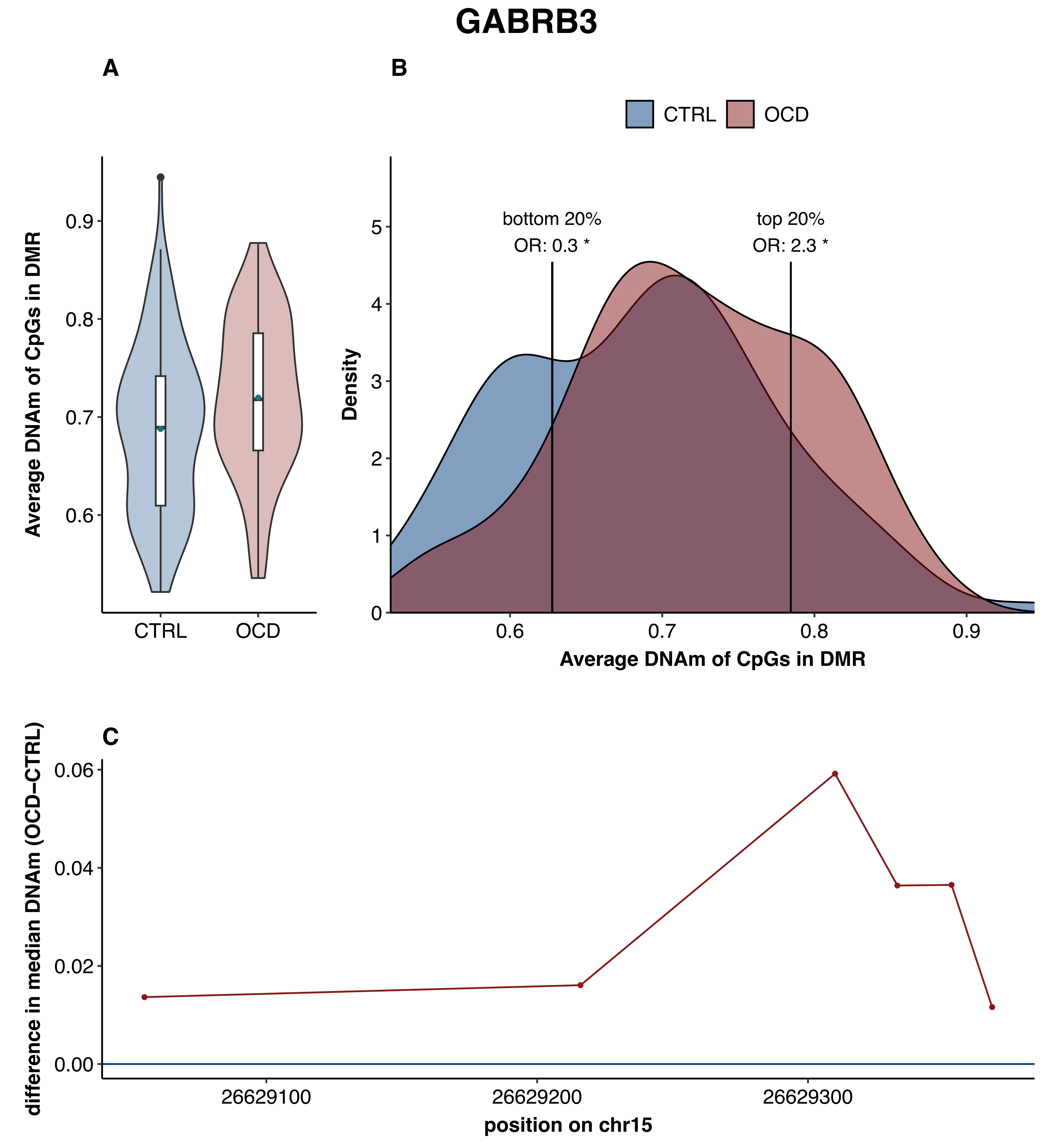
GABRB3
A
B
C
CTRL
OCD
Average DNAm of CpGs in DMR
0.9
0.8
0.7
0.6
bottom 20%
OR: 0.3 *
top 20%
OR: 2.3 *
Density
5
4
3
2
1
0
difference in median DNAm (OCD−CTRL)
0.06
0.04
0.02
0.00
26629100
26629200
26629300
position on chr15

### KIFC3

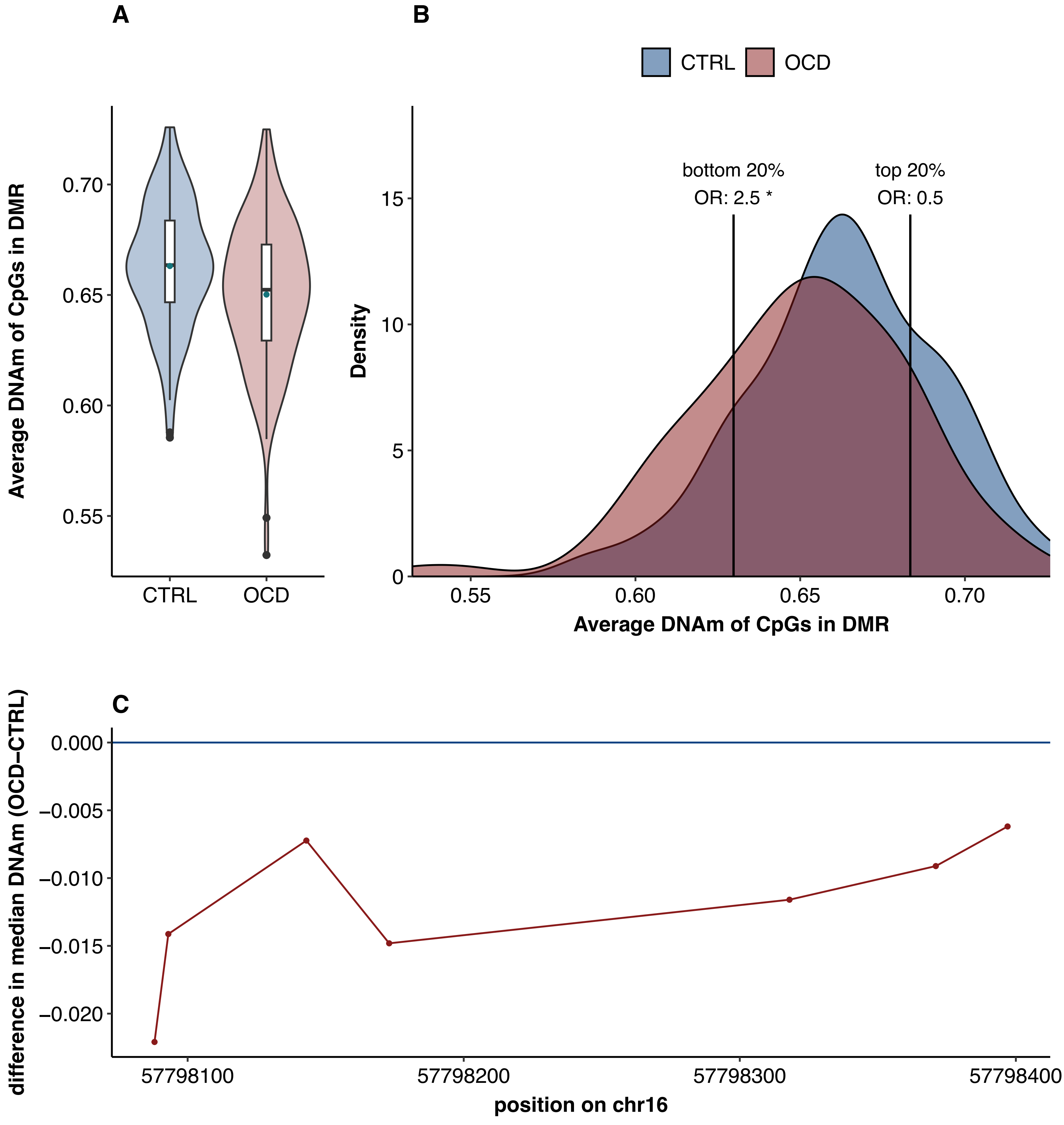
A
B
C
Average DNAm of CpGs in DMR
CTRL
OCD
0.70
0.65
0.60
0.55
bottom 20%
OR: 2.5 *
top 20%
OR: 0.5
Density
15
10
5
0
difference in median DNAm (OCD−CTRL)
0.000
−0.005
−0.010
−0.015
−0.020
57798100
57798200
57798300
57798400
position on chr16

### MOB3A

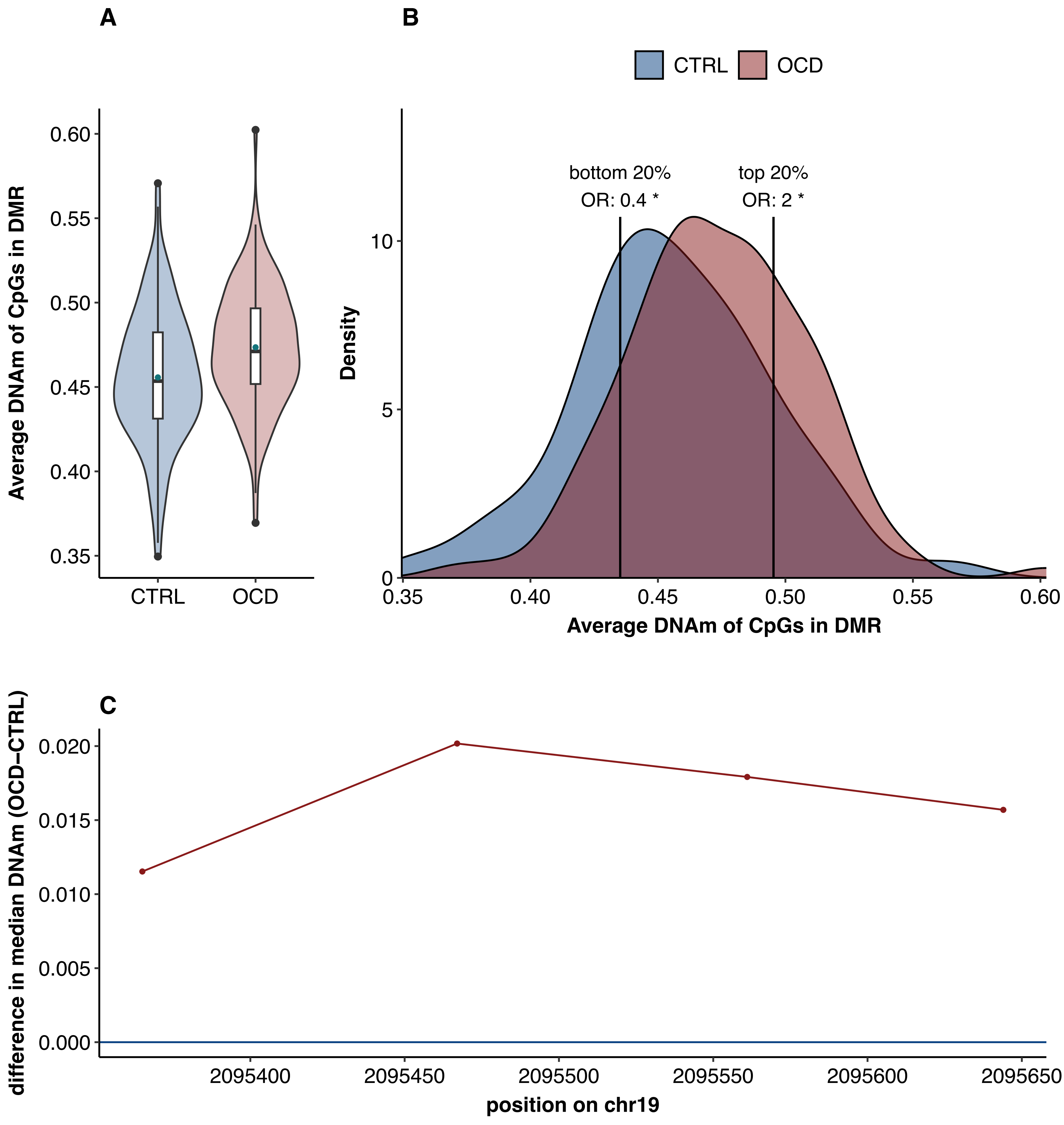

A
B
CTRL
OCD
0.60
0.55
0.50
0.45
0.40
0.35
Average DNAm of CpGs in DMR
CTRL
OCD
bottom 20%
OR: 0.4 *
top 20%
OR: 2 *
10
5
0
Density
0.35
0.40
0.45
0.50
0.55
0.60
Average DNAm of CpGs in DMR
C
0.020
0.015
0.010
0.005
0.000
difference in median DNAm (OCD−CTRL)
2095400
2095450
2095500
2095550
2095600
2095650
position on chr19

### PIWIL1;LOC101927786

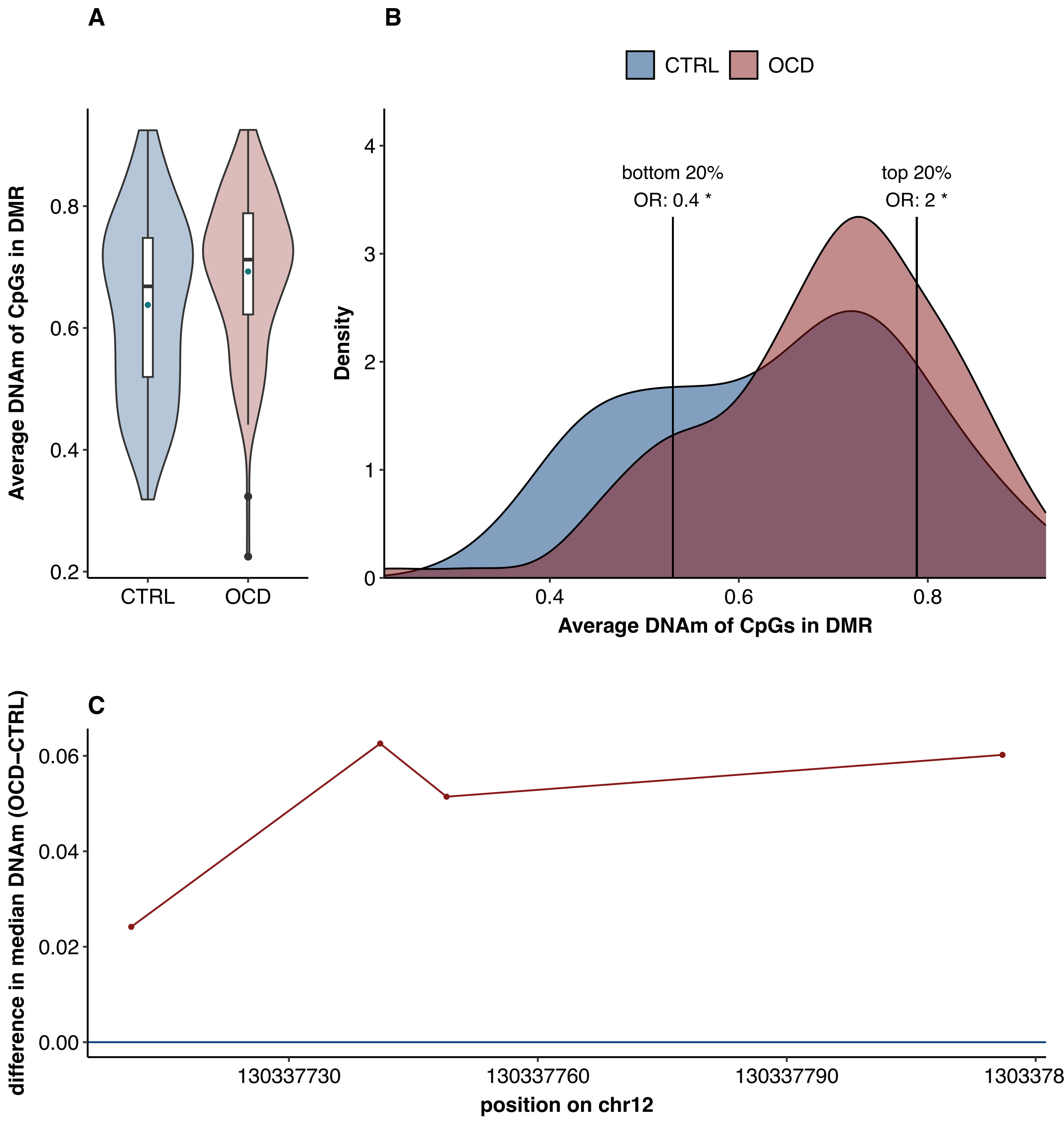

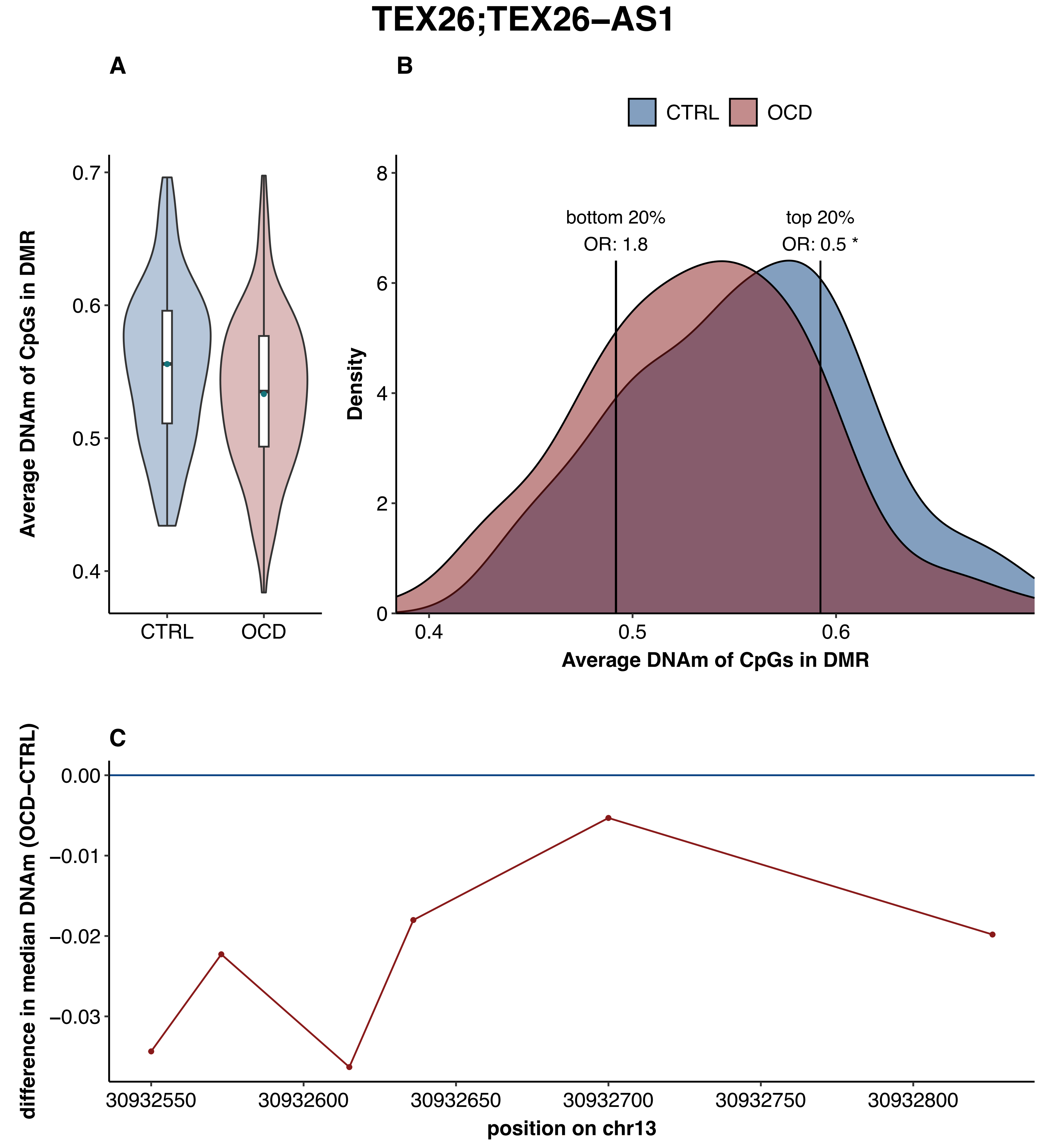

TEX26;TEX26−AS1
A
B
CTRL
OCD
0.7
0.6
0.5
0.4
Average DNAm of CpGs in DMR
CTRL
OCD
8
6
4
2
0
Density
bottom 20%
OR: 1.8
top 20%
OR: 0.5 *
0.4
0.5
0.6
Average DNAm of CpGs in DMR
C
0.00
−0.01
−0.02
−0.03
difference in median DNAm (OCD−CTRL)
30932550
30932600
30932650
30932700
30932750
30932800
position on chr13
